# The role of mitochondrial DNA copy number in neuropsychiatric disorders: A bidirectional two-sample Mendelian randomization study

**DOI:** 10.1101/2024.04.25.24306401

**Authors:** Mengqi Niu, Chen Chen, Michael Maes

## Abstract

**Background:** Inconsistent findings characterize studies on mitochondrial DNA copy number (mtDNA-CN) and its relation to neuropsychiatric disorders. This bidirectional two-sample Mendelian Randomization (MR) study explores potential causal links between mtDNA-CN and neuropsychiatric disorders, including Alzheimer’s disease, Attention-deficit/hyperactivity disorder, Anorexia nervosa, Autism spectrum disorder (ASD), Bipolar disorder, Major depressive disorder, Obsessive-compulsive disorder, Schizophrenia, Anxiety disorders, and Post-traumatic stress disorder.

**Methods:** Genetic associations with mtDNA-CN were drawn from the UK Biobank’s GWAS data (n = 395,718), while neuropsychiatric disorder data came from the Psychiatric Genomics Consortium and FinnGen Consortium. Three MR methods—Inverse Variance Weighting (IVW), MR-Egger, and Weighted Median—were used to establish relationships. Cochran’s Q test, MR-PRESSO, and MR-Egger’s intercept test assessed heterogeneity and pleiotropy. A leave-one-out analysis evaluated the impact of individual SNPs on MR results, and a bidirectional analysis examined the relationship between mtDNA-CN and neuropsychiatric disorders.

**Results:** The MR analysis indicated a causal relationship between mtDNA-CN and ASD using the IVW method (OR = 0.735, 95%CI: 0.597 to 0.905; P = 0.004). Conversely, a causal relationship was identified between Anxiety disorders and mtDNA-CN (β= 0.029, 95%CI: 0.010 to 0.048; P = 0.003). No causal associations were found for other disorders. Sensitivity tests corroborated the robustness of these findings.

**Conclusion:** In this study, potential causal relationships between mtDNA-CN and both ASD and Anxiety disorders were established. These findings provide insights into the biological mechanisms of mtDNA-CN on ASD and underscore the significance of mtDNA copy number as a potential biomarker for Anxiety disorders.

## 1. Introduction

Mitochondria are critical organelles regulating cellular energy production, metabolism, proliferation, and apoptosis (1). They possess their own genome known as mitochondrial DNA (mtDNA), which is a circular, intron-free, double-stranded, haploid molecule approximately 16.6 kb in size. This genome encodes several proteins essential for cellular energy metabolism (2). The number of mtDNA copies, which represents the quantity of mtDNA molecules in a single cell, is an important indicator of mitochondrial function and the cellular metabolic state (3). Recent studies have linked variations in mtDNA copy number (mtDNA-CN) to a variety of human diseases, including metabolic syndrome, cardiovascular diseases, neurodegenerative diseases, and cancer (4–6).

Mental disorders rank second in terms of premature mortality and disability, constituting one of the most significant public health challenges on a global scale. They are frequently linked to cognitive, emotional, or behavioral alterations. Mental disorders are believed to arise from intricate interplays between genetic predisposition and environmental factors (7, 8). Existing studies have shown that mtDNA-CN alterations are associated with Alzheimer’s disease (AD) (9, 10) attention-deficit/hyperactivity disorder (ADHD) (11–13) autism spectrum disorder (ASD) (14–17) bipolar disorder (BD) (18–21) major depressive disorder (MDD) (18, 22–27) schizophrenia (SCZ) (28–32) anxiety disorders (33, 34) and post-traumatic stress disorder (PTSD) (35–37), therefore, may be related to the occurrence and development of neuropsychiatric disorders. However, the observational results of different studies are controversial. To date, it is not clear whether there is a causal relationship between mtDNA-CN and these neuropsychiatric disorders. In addition, whether there is reverse causality for neuropsychiatric disorders affecting mtDNA-CN remains to be investigated.

The relationship between exposure and outcomes in traditional observational epidemiology is influenced by various factors, such as confounding elements and reverse causation. Moreover, this approach has limitations due to its time-consuming nature and high demands on resources (38). The recently developed Mendelian randomization (MR) design has been employed to investigate whether specific exposures have a causal effect on particular outcomes (39). MR methods use genetic variants such as single nucleotide polymorphisms (SNPs) as instrumental variables for exposures (40). By leveraging the random assortment of alleles during meiosis and the fixed allocation at conception, MR studies are less susceptible to confounding factors and reverse causality biases. Moreover, using summary statistics from genome-wide association studies (GWAS) in a two-sample MR approach significantly enhances the statistical power to infer causality (41). To date, no studies have utilized the MR design to investigate the association between mtDNA-CN and neuropsychiatric disorders, nor have any studies explored the reverse relationship using this method.

In this study, based on data from large-sample GWAS, we conducted a bidirectional two-sample MR analysis to investigate the association between mtDNA-CN and ten common neuropsychiatric disorders: AD, ADHD, anorexia nervosa (AN), ASD, BD, MDD, obsessive-compulsive disorder (OCD), SCZ, anxiety disorders, and PTSD.

## 2. Methods

### 2.1. Overall Study Design

We conducted a bidirectional two-sample MR analysis using the latest summary statistics from GWAS to explore the relationship between mtDNA-CN and neuropsychiatric disorders and to assess whether these neuropsychiatric disorders could lead to changes in mtDNA-CN. Our MR analysis adhered to three critical assumptions: (1) The selected SNPs are associated with the exposure; (2) The SNPS are independent of confounders of the exposure-outcome relationship (independence assumption); (3) The SNPS affect the outcome only through exposure (42). The study design included outcomes for 10 neuropsychiatric disorders: AD, ADHD, AN, ASD, BD, MDD, OCD, SCZ, anxiety disorders, and PTSD. Figure 1 illustrates the study workflow.

**Figure 1.**
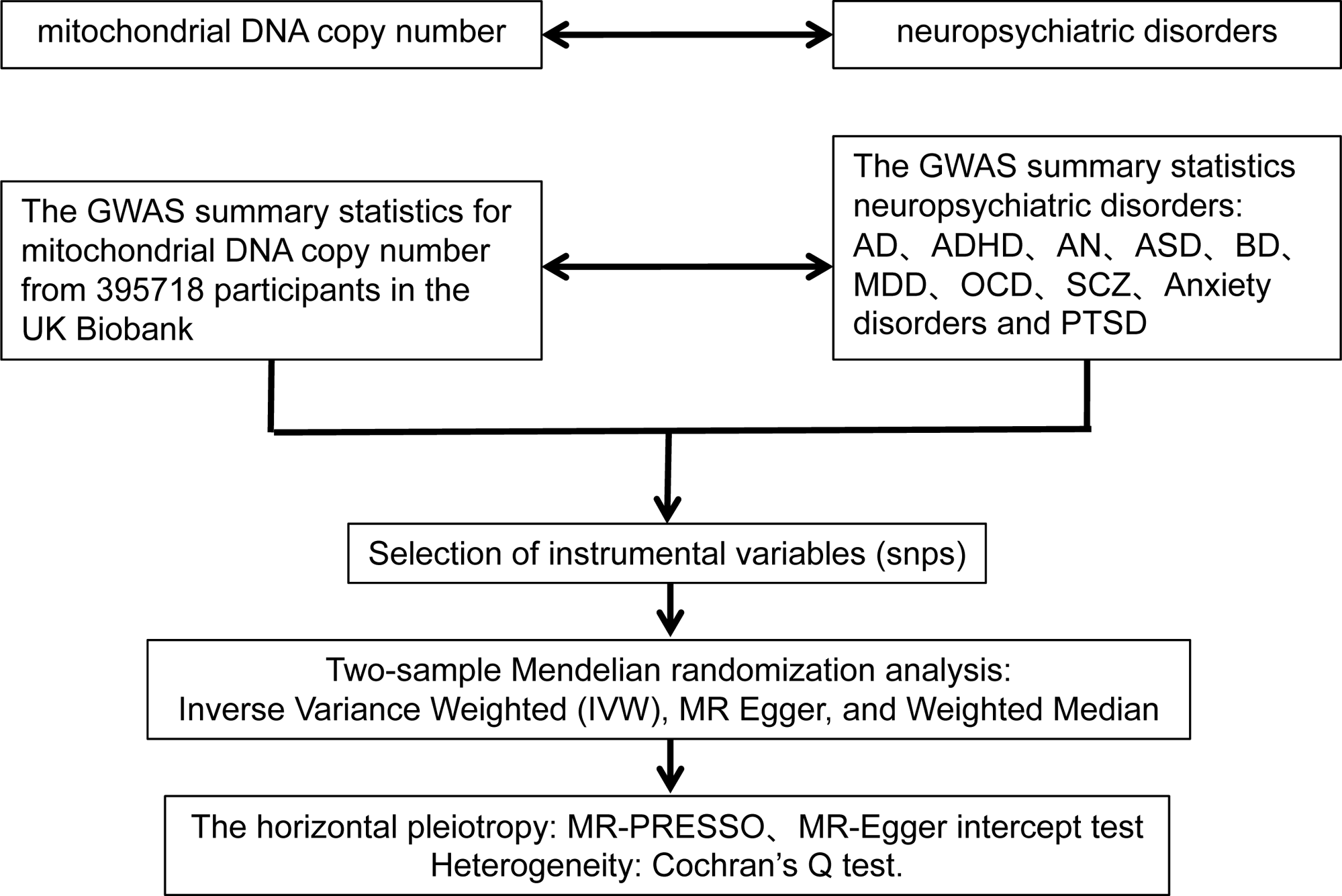
The flowchart of bidirectional Mendelian randomization study explores the causal relationship between mitochondrial DNA copy number and neuropsychiatric disorders. AD: Alzheimer’s disease; ADHD: Attention-deficit/hyperactivity disorder; AN: Anorexia nervosa; ASD: Autism spectrum disorder; BD: Bipolar disorder; MDD: Major depressive disorder; OCD: Obsessive compulsive disorder; SCZ: Schizophrenia; PTSD: Post-traumatic stress disorder.

### 2.2. Data Sources

**Table 1** presents the GWAS meta-analysis information regarding mtDNA-CN, involving 395,718 participants from the UK Biobank of European ancestries. It used a novel approach called “AutoMitoC” developed by Chong et al. (43) to identify novel common and rare genetic determinants of mtDNA-CN. This represents the most extensive genetic evaluation published to date.

**Table 1.** Characteristics of the used genome-wide association study in the study.

| Phenotypes | Study/consortium | Cases/ controls | Ancestry | PubMed ID/Website |
| --- | --- | --- | --- | --- |
| Mitochondrial DNA copy number | UK Biobank | 395,718 | European | 35023831 |
| AD: Alzheimer's disease | PGC | 71,880/383,378 | European | 30617256 |
| ADHD: Attention-deficit/hyperactivity disorder | PGC | 20,183/35,191 | European | 29325848 |
| AN: Anorexia nervosa | PGC | 3,495/10,982 | European | 28494655 |
| ASD: Autism spectrum disorder | PGC | 18,381/27,969 | European | 30804558 |
| BD: Bipolar disorder | PGC | 41,917/371,549 | European | 34002096 |
| MDD: Major depressive disorder | PGC | 59,851/113,154 | European | 29700475 |
| OCD: Obsessive compulsive disorder | PGC | 2,688/7,037 | European | 28761083 |
| SCZ: Schizophrenia | PGC | 52,017/75,889 | European | 35396580 |
| Anxiety disorders | FinnGen | 44,663/301,879 | European | https://storage.googleapis.com/finngen-public-data-r10/summary_stats/finngen_R10_KRA_PSY_ANXIETY_EXMORE.gz |
| PTSD:<br>Post-traumatic stress disorder | FinnGen | 2,615/368,054 | European | https://storage.googleapis.com/finngen-public-data-r10/summary_stats/finngen_R10_F5_PTSD.gz |

PGC, Psychiatric Genomics Consortium.

Information about neuropsychiatric disorders related to GWAS summary statistics are shown in **Table 1** and sourced from the Psychiatric Genomics Consortium (PGC: https://www.med.unc.edu/pgc/). Sample sizes are detailed as follows: AD (71,880 cases and 383,378 controls) (44), ADHD (20,183 cases and 35,191 controls) (45), AN (3,495 cases and 10,982 controls) (46), ASD (18,381 cases and 27,969 controls) (47), BD (41,917 cases and 371,549 controls) (48), MDD (59,851 cases and 113,154 controls) (49), OCD (2,688 cases and 7,037 controls) (50), SCZ (52,017 cases and 75,889 controls) (51). Summary statistics for anxiety disorders (44,663 cases and 301,879 controls) and PTSD (2,615 cases and 368,054 controls) were published in 2023 by the FinnGen Consortium (https://r10.finngen.fi/). The study exclusively used European population data to minimize population stratification biases.

### 2.3. Selection of Instrumental Variables

Strict criteria were employed to select suitable instrumental variables. (1) All SNPs achieving genome-wide significance (P < 5 × 10^−8^) were included. When the number of suitable instrumental variables was restricted to 10 or fewer, the p-value threshold was relaxed to 1 × 10^−6^ (52). (2) To ensure the independence of the instruments used for exposure, SNPs in linkage disequilibrium (LD) (r^2^ < 0.001 within a 10,000 kb window) were excluded (52). (3) Palindromic SNPs with a medium allele frequency (MAF) were removed (53). (4) The LDtrait Tool (https://ldlink.nih.gov) (54) was used to assess potential confounders including alcohol consumption, smoking behavior, socioeconomic status, and education (55). (5) In addition, the proportion of variance explained (PVE) by SNPS was assessed using the following formula: PVE = 2 × MAF × (1 − MAF) × beta^2^ (56), and the F-statistic was calculated to determine the strength of the SNP using the following formula: F = PVE × (N − 2)/(1 − PVE) (56). In cases where MAF data were not provided in the original study, an alternative formula was used: F = beta^2^/se^2^ (57). The statistical power of F > 10 indicates a strong correlation (58).

### 2.4. Statistical Analysis

Three MR methods were used to investigate the causal relationship between mtDNA-CN and neuropsychiatric disorders: Inverse Variance Weighted (IVW), MR-Egger, and Weighted Median (WM). The Cochran’s Q test within the IVW method was used to detect heterogeneity among SNPs, with a significance level set at P < 0.05. The MR-PRESSO analysis identified and corrected for potential pleiotropic outliers, and the intercept P-value in the MR-Egger analysis evaluated horizontal pleiotropy, with significance considered at P < 0.05 (59). The leave-one-out method was implemented to determine whether the estimates were driven by any single SNP (55).

To ensure the reliability of the final analysis results, the following screening criteria were used as filters to identify robust significant causality: (1) IVW method is considered the primary approach for determining the association between exposures and outcomes. MR-Egger and WM methods reinforce the IVW results by analyzing the link between mtDNA-CN and neuropsychiatric disorders from multiple perspectives. (2) The direction of the MR analysis results (beta values) is consistent across all three methods. (3) Given our reliance on the IVW method, we adjust all P values derived from IVW using the False Discovery Rate (FDR) correction. The adjusted results are denoted as P*. A statistical significance threshold is set at 0.05. A P value ≤ 0.05 with P* ≤ 0.05 indicates a significant causal relationship, whereas a P value ≤ 0.05 with P* > 0.05 suggests evidence of a potential causal relationship.

Statistical analyses were performed using the statistical software R (version 4.3.2) with packages including MendelianRandomization, MR-PRESSO, and TwoSampleMR. For binary outcomes, odds ratios (OR) and their 95% confidence intervals (CI) quantified the strength of the causal relationships. For continuous outcomes, results were reported as beta and standard error (SE) along with their respective 95% CIs.

### 2.5. Ethical Approval and Consent to Participate

This study was reported according to the Strengthening the Reporting of Observational Studies in Epidemiology Using MR (STROBE-MR) checklist (60). Our analyses were based on publicly available data that had been approved by relevant review boards. Therefore, no additional ethical approval is required.

## 3. Results

### 3.1. Genetic prediction of the relationship between mtDNA-CN and neuropsychiatric disorders

#### 3.1.1. Selection of Instrumental Variables

To ensure precision in our analysis, the initial pool of 11,173,383 SNPs related to mtDNA-CN was reduced following criteria for significance (P < 5 × 10^−8^) and LD (r^2^ < 0.001 within a 10,000 kb window). This refinement process left 67 SNPs eligible for the main analysis, as detailed in **Additional file 1: Table S1**.

For AD, 64 of these SNPs were successfully extracted. After removing palindromic sequences with MAF, 55 SNPs remained. These had an F-statistic 88.18 (range of 32.25 to 461.23). In ADHD, from the original 67 SNPs, 58 were extracted and 51 remained after the same exclusion criteria, showing an F-statistic 86.69 (range from 32.25 to 461.22). Similar analysis in AN and ASD resulted in 63 initial SNPs, with 56 remaining post-exclusion, and F-statistics 86.76 (from 32.25 to 461.22). For BD, we started with 60 SNPs and retained 53, with F-statistics 89.43 (ranging from 32.25 to 461.22). This pattern was mirrored in MDD and OCD where from initial figures of 61 and 60 SNPs, 53 were retained in each case, both with an F-statistic 88.46 (range of 32.25 to 461.22). SCZ saw 60 initial SNPs with 53 remaining, exhibiting an F-statistic 89.43 (range of 32.25 to 461.22). For Anxiety Disorders and PTSD, 61 SNPs were identified, and 53 remained after exclusions, with an F-statistic 86.08 (ranging from 32.25 to 461.23). Additionally, no significant associations with confounding factors such as alcohol consumption, smoking, socioeconomic status, and education were found when these SNPs were assessed using the LDtrait Tool (https://ldlink.nih.gov). Detailed information on the SNPs finally used as instrumental variables to explore the relationship between mtDNA-CN and neuropsychiatric disorders is available in **Additional file 1: Tables S2-S11**.

#### 3.1.2 MR Analysis Results

As shown in **Table 2**, the primary IVW analysis indicated a statistically significant causal relationship between mtDNA-CN and ASD (OR = 0.735, 95%CI: 0.597 to 0.905; P = 0.004). The secondary analysis methods WM also showed statistical significance (OR = 0.735, 95%CI: 0.550 to 0.982; P = 0.037). However, no evidence was found for a causal relationship between mtDNA-CN and AD, ADHD, AN, BD, MDD, OCD, SCZ, Anxiety Disorders, or PTSD across all three MR analysis methods (IVW, MR-Egger, WM). Sensitivity analyses with the MR-PRESSO method detected horizontal pleiotropic outliers only in AD, with no outliers found for other disorders. After excluding the outlier SNP, the MR-Egger intercept of all neuropsychiatric disorders showed no evidence of horizontal pleiotropy (P > 0.05). The Cochran’s Q test for heterogeneity identified significant heterogeneity only among SNPs for BD, MDD, and SCZ; no significant heterogeneity was observed for other disorders. The leave-one-out method confirmed that the causal relationships were not driven by any individual SNP. Scatter plots of the forward analysis and the leave-one-out analysis graphs for each SNP-neuropsychiatric disorder association are summarized in **Figure 2** and **Additional file 1: Figure S1**, respectively.

**Figure 2.**
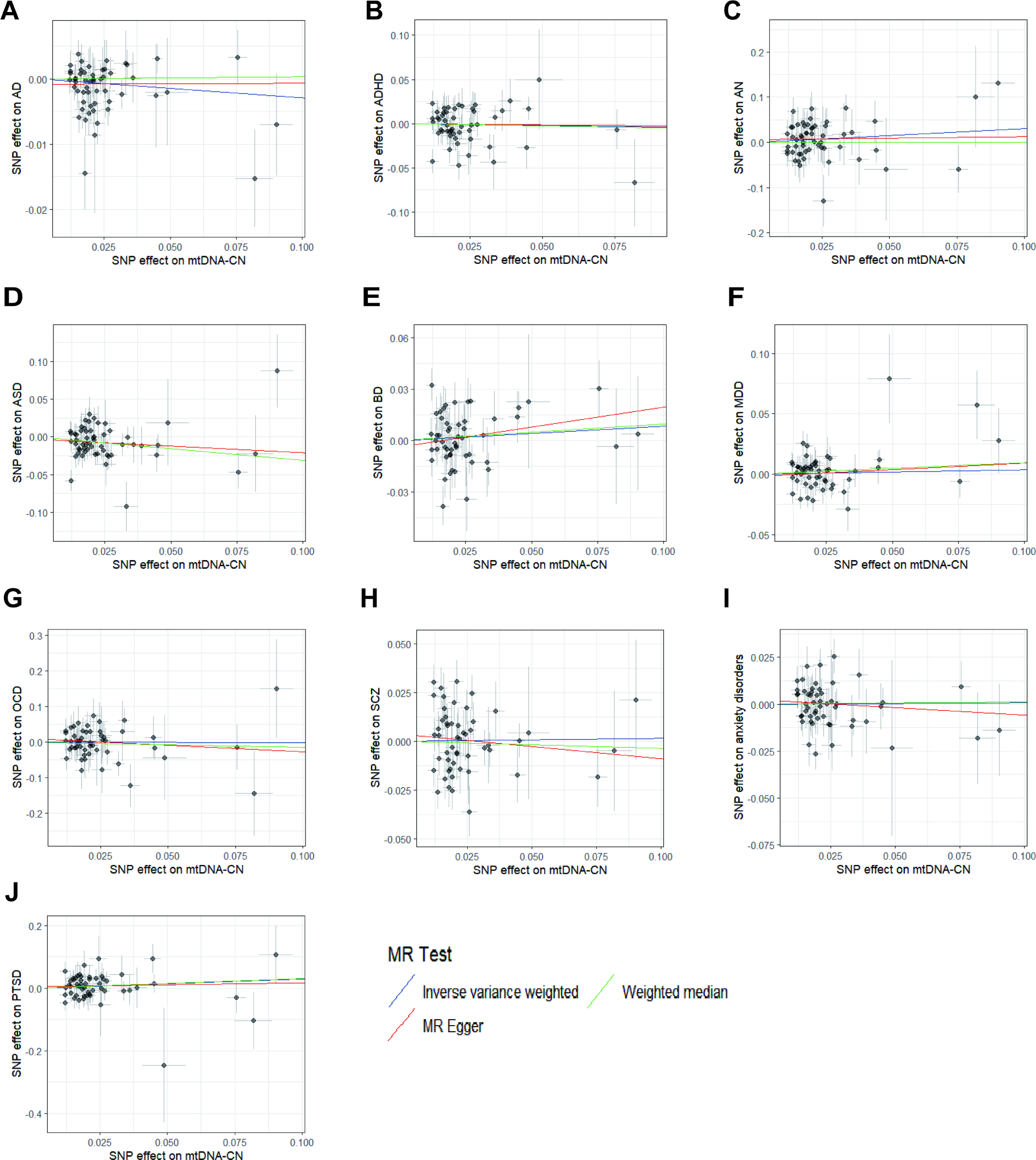
The forward MR analyses: Scatter plots of the association between mtDNA copy number and neuropsychiatric disorders. A Alzheimer’s disease, B attention-deficit/hyperactivity disorder, C anorexia nervosa, D autism spectrum disorder, E bipolar disorder, F major depressive disorder, G obsessive compulsive disorder, H Schizophrenia, I anxiety disorders, J post-traumatic stress disorder. Lines in blue, red, and green represent IVW, MR-Egger, and weighted median methods. IVW inverse variance weighting, mtDNA-CN mtDNA copy number, SNP single nucleotide polymorphism.

**Table 2.** Mendelian randomization estimates between genetically predicted mtDNA copy number and the risk of neuropsychiatric disorders.

| Exposure | Outcome | No. of SNPs | Methods | OR | Lower 95% CI | Upper 95% CI | P | P* | Cochran's Q test (I<sup>2</sup>) | P | MR-Egger intercept (P value) | Outliers from MR-PRESSO |
| --- | --- | --- | --- | --- | --- | --- | --- | --- | --- | --- | --- | --- |
| mtDNA-CN | AD | 54 | IVW | 0.971 | 0.940 | 1.004 | 0.088 | 0.438 | 59.968 | 0.238 | 0.400 | rs1065853 |
|  |  |  | MR-Egger | 1.002 | 0.926 | 1.083 | 0.968 |  |  |  |  |  |
|  |  |  | WM | 1.004 | 0.955 | 1.054 | 0.888 |  |  |  |  |  |
| mtDNA-CN | ADHD | 51 | IVW | 0.960 | 0.776 | 1.187 | 0.705 | 1.000 | 61.105 | 0.135 | 0.946 | NA |
|  |  |  | MR-Egger | 0.975 | 0.589 | 1.615 | 0.922 |  |  |  |  |  |
|  |  |  | WM | 0.945 | 0.700 | 1.275 | 0.710 |  |  |  |  |  |
| mtDNA-CN | AN | 56 | IVW | 1.353 | 0.907 | 2.019 | 0.139 | 0.462 | 53.013 | 0.551 | 0.594 | NA |
|  |  |  | MR-Egger | 1.070 | 0.415 | 2.760 | 0.890 |  |  |  |  |  |
|  |  |  | WM | 1.001 | 0.533 | 1.882 | 0.996 |  |  |  |  |  |
| mtDNA-CN | ASD | 56 | IVW | 0.735 | 0.597 | 0.905 | 0.004 | 0.038 | 66.714 | 0.134 | 0.563 | NA |
|  |  |  | MR-Egger | 0.836 | 0.516 | 1.355 | 0.470 |  |  |  |  |  |
|  |  |  | WM | 0.735 | 0.550 | 0.982 | 0.037 |  |  |  |  |  |
| mtDNA-CN | BD | 53 | IVW | 1.088 | 0.921 | 1.286 | 0.321 | 0.642 | 85.458 | 0.002 | 0.422 | NA |
|  |  |  | MR-Egger | 1.260 | 0.851 | 1.866 | 0.253 |  |  |  |  |  |
|  |  |  | WM | 1.100 | 0.894 | 1.354 | 0.369 |  |  |  |  |  |
| mtDNA-CN | MDD | 53 | IVW | 1.035 | 0.904 | 1.185 | 0.617 | 1.000 | 75.320 | 0.019 | 0.608 | NA |
|  |  |  | MR-Egger | 1.115 | 0.815 | 1.526 | 0.499 |  |  |  |  |  |
|  |  |  | WM | 1.099 | 0.919 | 1.314 | 0.300 |  |  |  |  |  |
| mtDNA-CN | OCD | 53 | IVW | 0.997 | 0.617 | 1.612 | 0.990 | 0.990 | 37.926 | 0.928 | 0.485 | NA |
|  |  |  | MR-Egger | 0.691 | 0.224 | 2.135 | 0.524 |  |  |  |  |  |
|  |  |  | WM | 0.872 | 0.416 | 1.827 | 0.717 |  |  |  |  |  |
| mtDNA-CN | SCZ | 53 | IVW | 1.015 | 0.846 | 1.218 | 0.871 | 1.000 | 119.179 | < 0.001 | 0.484 | NA |
|  |  |  | MR-Egger | 0.882 | 0.572 | 1.359 | 0.571 |  |  |  |  |  |
|  |  |  | WM | 0.964 | 0.789 | 1.177 | 0.717 |  |  |  |  |  |
| mtDNA-CN | Anxiety disorders | 53 | IVW | 1.008 | 0.895 | 1.135 | 0.896 | 0.995 | 62.448 | 0.152 | 0.484 | NA |
|  |  |  | MR-Egger | 0.923 | 0.703 | 1.212 | 0.567 |  |  |  |  |  |
|  |  |  | WM | 1.011 | 0.861 | 1.187 | 0.892 |  |  |  |  |  |
| mtDNA-CN | PTSD | 53 | IVW | 1.345 | 0.901 | 2.008 | 0.147 | 0.368 | 40.390 | 0.879 | 0.696 | NA |
|  |  |  | MR-Egger | 1.138 | 0.452 | 2.868 | 0.785 |  |  |  |  |  |
|  |  |  | WM | 1.369 | 0.753 | 2.488 | 0.303 |  |  |  |  |  |

mtDNA-CN: mitochondrial DNA copy number; AD: Alzheimer's disease; ADHD: Attention-deficit/hyperactivity disorder; AN: Anorexia nervosa; ASD: Autism spectrum disorder; BD: Bipolar disorder; MDD: Major depressive disorder; OCD: Obsessive compulsive disorder; SCZ:Schizophrenia; PTSD: Post-traumatic stress disorder; SNPs single nucleotide polymorphisms.
The random-effects inverse variance-weighted (IVW) method was used as the primary approach, while other methods including MR-Egger, weighted median (WM) methods.Odds ratios (OR) for associations between genetically predicted mtDNA copy number and mental disorders. P\* represents the P adjusted by False discovery rate (FDR). Cochran's Q-test is used for heterogeneity testing, and a $P < 0.05$ is considered significant. MR-PRESSO analysis is used to identify and correct potential level pleiotropy outliers. MR-Egger intercept analysis was used to evaluate level pleiotropy, and P value $< 0.05$ was considered significant.

In our analysis examining the causal relationship between mtDNA-CN and ASD, the directionality (beta values) of the MR results were consistent across three different methodologies. Furthermore, upon adjusting the P-values from the IVW method using the FDR, the adjusted P-value (P*) was 0.038. This suggests a statistically significant causal association between mtDNA-CN and ASD.

### 3.2. Genetic prediction of the relationship between neuropsychiatric disorders and mtDNA-CN

#### 3.2.1 Selection of Instrumental Variables

In our reverse causality analysis of the relationship between neuropsychiatric disorders and mtDNA-CN, SNPs were rigorously selected based on statistical significance (P < 5 × 10^-8^) and LD (r^2^ < 0.001 within a 10,000 kb window). This selection resulted in the retention of 30 SNPs for AD, 12 for ADHD, 52 for BD, and 158 for SCZ. After this filtering, 29 SNPs for AD were extracted from the SNPs of mtDNA-CN and further reduced to 23 post-exclusion of palindromic sequences with MAF, yielding F-statistics 93.03 (ranging from 30.29 to 670.46). For ADHD, of the 12 extracted SNPs, 11 remained after similar exclusions, with F-statistics 34.11 (ranging from 30.57 to 52.71). In BD, 51 SNPs were initially extracted, and 44 remained, showing F-statistics 38.11 (range from 30.08 to 79.65). For SCZ, 158 SNPs were extracted, and 130 remained post-exclusion, with F-statistics 45.35 (ranging from 29.46 to 175.27).

For AN, ASD, MDD, OCD, anxiety disorders, and PTSD, where the number of suitable instrumental variables was fewer than 10, the IV selection p-value threshold was relaxed to 1 × 10^-6^. This included 5 SNPs for AN, 16 for ASD, 22 for MDD, 2 for OCD, 50 for anxiety disorders, and 2 for PTSD. After this filtering, 4 SNPs for AN were extracted from the SNPs of mtDNA-CN and further reduced to 4 post-exclusion of palindromic sequences with MAF, yielding F-statistics 28.02 (ranging from 24.11 to 34.41). For ASD, of the 15 extracted SNPs, 14 remained after similar exclusions, with F-statistics 26.59 (ranging from 24.07 to 35.77). In MDD, 22 SNPs were initially extracted, and 17 remained, showing F-statistics 28.54 (range from 24.57 to 43.66). For OCD, 2 SNPs were extracted, and 2 remained post-exclusion, with F-statistics 25.09 (ranging from 24.62 to 25.57). In anxiety disorders, 50 SNPs were initially extracted, and 41 remained, showing F-statistics 28.26 (range from 24.01 to 46.75). For PTSD, 2 SNPs were extracted, and 2 remained post-exclusion, with F-statistics 27.58 (ranging from 27.49 to 27.66). Additionally, no significant associations with confounding factors such as alcohol consumption, smoking, socioeconomic status, and education were found when these SNPs were assessed using the LDtrait Tool (https://ldlink.nih.gov). Detailed information on the genetic variants (SNPs) finally used as instrumental variables for the reverse relationship between neuropsychiatric disorders and mtDNA-CN is available in **Additional file 1: Table S12-21**.

#### 3.2.2 MR Analysis Results

As shown in **Table 3**, the primary IVW analysis revealed a statistically significant causal relationship between anxiety disorders and mtDNA-CN (β = 0.029, 95%CI: 0.010 to 0.048; P = 0.003). However, no evidence of a causal relationship was found between mtDNA-CN and AD, ADHD, AN, ASD, BD, MDD, OCD, SCZ, or PTSD across all three MR analysis methods (IVW, MR-Egger, WM). Sensitivity analyses with the MR-PRESSO method detected one horizontal pleiotropic outlier in AD, BD, SCZ, and two in anxiety disorders, with no outliers observed for other disorders. After excluding these outliers, the MR-Egger intercept of all outcomes showed no evidence of horizontal pleiotropy (P > 0.05). The Cochran’s Q test for heterogeneity identified significant heterogeneity only among SNPs for AD, BD, MDD, and SCZ; no significant heterogeneity was observed for other disorders. Meanwhile, the “leave one” method confirms that a single SNP does not affect causal relationships. The reverse analysis scatter plots and the leave-one-out analysis graphs for each SNP of neuropsychiatric disorder association with mtDNA-CN are summarized in **Figure 3** and **Additional file 1: Figure S2**, respectively. For OCD and PTSD, only 2 SNPs were available for analysis, which was insufficient for comprehensive MR-Egger, WM, MR-PRESSO methods, and leave-one-out analysis.

**Figure 3.**
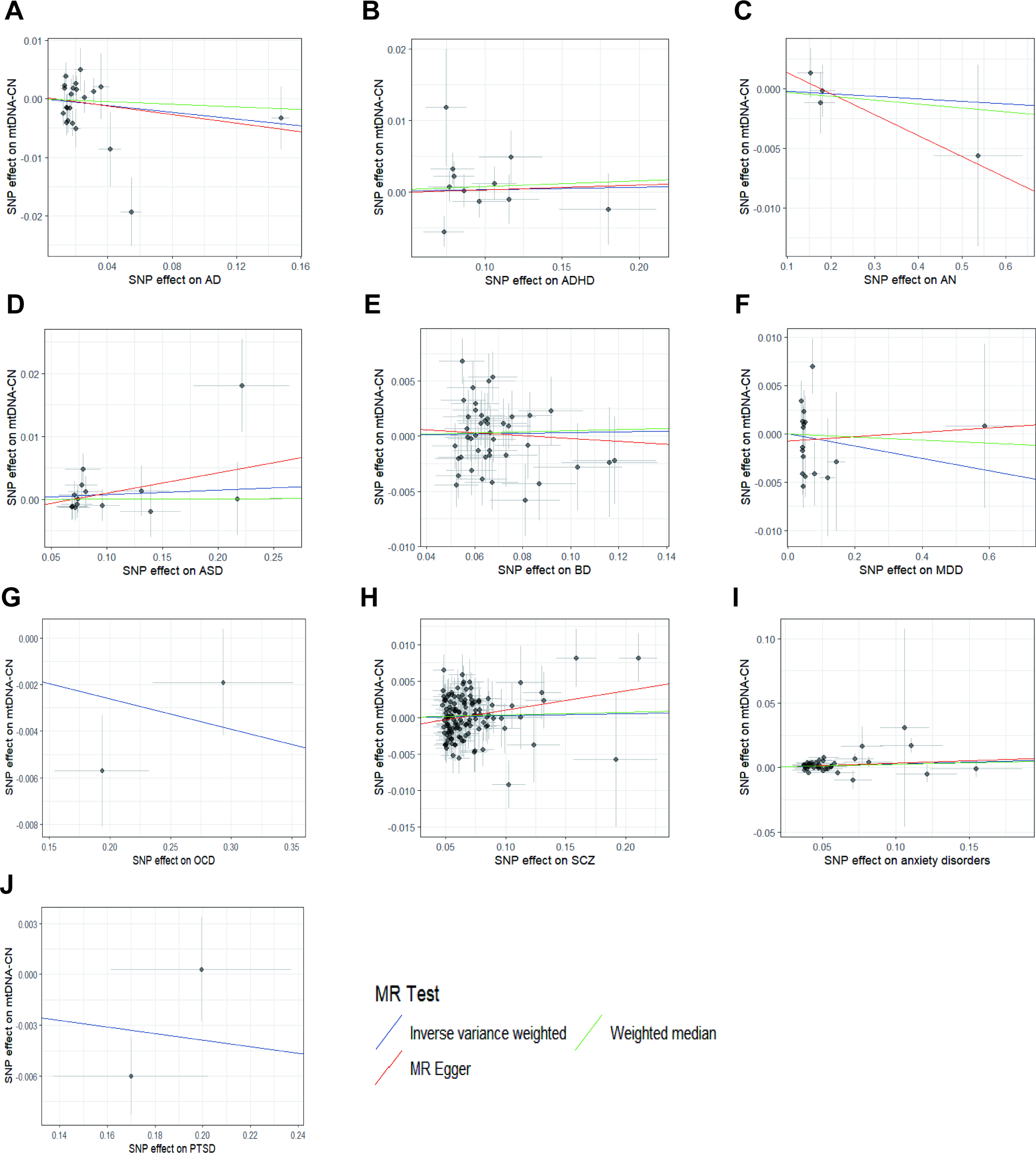
The reverse MR analyses: Scatter plots of the association between neuropsychiatric disorders and mtDNA copy number. A Alzheimer’s disease, B attention-deficit/hyperactivity disorder, C anorexia nervosa, D autism spectrum disorder, E bipolar disorder, F major depressive disorder, G obsessive compulsive disorder, H Schizophrenia, I anxiety disorders, J post-traumatic stress disorder. Lines in blue, red, and green represent IVW, MR-Egger, and weighted median methods. IVW inverse variance weighting, mtDNA-CN mtDNA copy number, SNP single nucleotide polymorphism.

**Table 3.** Reverse mendelian randomization estimates on associations of genetically predicted neuropsychiatric disorders and mtDNA copy number.

| Exposure | Outcome | No. of SNPs | Methods | beta | SE | Lower 95% CI | Upper 95% CI | P | P* | Cochran's Q test (I2) | P | MR-Egger intercept (P value) | Outliers from MR-PRESSO |
| --- | --- | --- | --- | --- | --- | --- | --- | --- | --- | --- | --- | --- | --- |
| AD | mtDNA-CN | 22 | IVW | -0.029 | 0.029 | -0.087 | 0.029 | 0.325 | 0.814 | 35.996 | 0.022 | 0.848 | rs41290120 |
|  |  |  | MR-Egger | -0.037 | 0.050 | -0.135 | 0.062 | 0.474 |  |  |  |  |  |
|  |  |  | WM | -0.011 | 0.032 | -0.075 | 0.052 | 0.730 |  |  |  |  |  |
| ADHD | mtDNA-CN | 11 | IVW | 0.003 | 0.010 | -0.017 | 0.024 | 0.748 | 0.831 | 14.926 | 0.135 | 0.949 | NA |
|  |  |  | MR-Egger | 0.007 | 0.051 | -0.093 | 0.106 | 0.900 |  |  |  |  |  |
|  |  |  | WM | 0.008 | 0.012 | -0.015 | 0.031 | 0.502 |  |  |  |  |  |
| AN | mtDNA-CN | 4 | IVW | -0.002 | 0.007 | -0.015 | 0.011 | 0.757 | 0.757 | 1.051 | 0.789 | 0.508 | NA |
|  |  |  | MR-Egger | -0.018 | 0.021 | -0.058 | 0.023 | 0.482 |  |  |  |  |  |
|  |  |  | WM | -0.003 | 0.008 | -0.019 | 0.012 | 0.684 |  |  |  |  |  |
| ASD | mtDNA-CN | 14 | IVW | 0.007 | 0.008 | -0.008 | 0.023 | 0.362 | 0.724 | 12.195 | 0.512 | 0.340 | NA |
|  |  |  | MR-Egger | 0.032 | 0.026 | -0.019 | 0.084 | 0.245 |  |  |  |  |  |
|  |  |  | WM | 0.001 | 0.011 | -0.020 | 0.022 | 0.960 |  |  |  |  |  |
| BD | mtDNA-CN | 43 | IVW | 0.003 | 0.006 | -0.009 | 0.016 | 0.613 | 0.766 | 58.538 | 0.046 | 0.670 | rs41315395 |
|  |  |  | MR-Egger | -0.013 | 0.038 | -0.088 | 0.062 | 0.737 |  |  |  |  |  |
|  |  |  | WM | 0.005 | 0.008 | -0.010 | 0.020 | 0.520 |  |  |  |  |  |
| MDD | mtDNA-CN | 17 | IVW | -0.006 | 0.012 | -0.030 | 0.017 | 0.598 | 0.854 | 27.340 | 0.038 | 0.590 | NA |
|  |  |  | MR-Egger | 0.002 | 0.020 | -0.037 | 0.041 | 0.911 |  |  |  |  |  |
|  |  |  | WM | -0.002 | 0.014 | -0.029 | 0.026 | 0.911 |  |  |  |  |  |
| OCD | mtDNA-CN | 2 | IVW | -0.013 | 0.010 | -0.033 | 0.007 | 0.209 | 1.000 | 2.452 | 0.117 | - | - |
|  |  |  | MR-Egger |  |  |  |  |  |  |  |  |  |  |
|  |  |  | WM |  |  |  |  |  |  |  |  |  |  |
| SCZ | mtDNA-CN | 129 | IVW | 0.003 | 0.004 | -0.004 | 0.010 | 0.452 | 0.753 | 162.225 | 0.022 | 0.072 | rs34555420 |
|  |  |  | MR-Egger | 0.026 | 0.014 | 0.000 | 0.053 | 0.054 |  |  |  |  |  |
|  |  |  | WM | 0.004 | 0.005 | -0.006 | 0.013 | 0.454 |  |  |  |  |  |
| Anxiety disorders | mtDNA-CN | 39 | IVW | 0.029 | 0.010 | 0.010 | 0.048 | 0.003 | 0.030 | 46.918 | 0.152 | 0.798 | rs4955420、<br>rs6123193 |
|  |  |  | MR-Egger | 0.038 | 0.038 | -0.036 | 0.112 | 0.318 |  |  |  |  |  |
|  |  |  | WM | 0.024 | 0.013 | -0.001 | 0.050 | 0.062 |  |  |  |  |  |
| PTSD | mtDNA-CN | 2 | IVW | -0.019 | 0.018 | -0.055 | 0.016 | 0.286 | 0.955 | 3.190 | 0.074 | - | - |
|  |  |  | MR-Egger |  |  |  |  |  |  |  |  |  |  |
|  |  |  | WM |  |  |  |  |  |  |  |  |  |  |

In examining the statistically significant causal relationship between anxiety disorders and mtDNA-CN, the directional consistency (beta values) of the MR results was observed across three different methodologies. Furthermore, after adjusting the P-values obtained from the IVW method using the FDR, the adjusted P-value (P*) was 0.030. This indicates a significant causal association between anxiety disorders and mtDNA-CN.

## 4. Discussion

Leveraging a vast array of publicly available genetic data, our study explored the causal relationships between mtDNA-CN and neuropsychiatric disorders. To our knowledge, this is the first comprehensive study to utilize bidirectional two-sample MR to investigate the association between mtDNA-CN and various neuropsychiatric disorders from a genetic predictive perspective. We found significant causal effects of mtDNA-CN on ASD and of anxiety disorders on mtDNA-CN. No other forward or reverse causal relationships were identified between mtDNA-CN and other neuropsychiatric disorders (including AD, ADHD, AN, BD, MDD, OCD, SCZ, PTSD).

This MR study revealed that a decrease in mtDNA-CN could increase the likelihood of developing ASD. Two observational studies observed a reduction in mtDNA-CN in peripheral blood mononuclear cells and leukocytes among patients with ASD, consistent with our findings (14, 15). A reduction in mtDNA-CN implies a lower DNA copy number per mitochondrion, potentially leading to diminished ATP synthesis capacity, thereby compromising the energy supply to neuronal cells, affecting their normal function and development, particularly during early brain development stages (61–63). According to a recent gene network analysis that examined mRNA expression in the brains of children with ASD, it was found that ASD is associated with mitochondrial dysfunctions, specifically in the electron transport chain and ATP production (64). Insufficient energy supply may also trigger programmed cell death (apoptosis), especially in neuronal cells, impacting the structural and functional development of the brain and further relating to autism symptoms (61, 63). Mitochondrial dysfunction may affect the synthesis and release of neurotransmitters such as glutamate and GABA, key chemicals in regulating mood, cognition, and behavior, potentially linked to the social and communication challenges observed in autism (62, 63). However, Yoo et al. (16) observed an increase in peripheral blood mtDNA-CN in patients with ASD, which was associated with neurological manifestations and communication; similarly, Chen et al. (17) observed a higher relative mtDNA-CN in children with autism compared to healthy controls. These observations do not contradict our findings; in fact, they may reveal the complex regulatory mechanisms of mitochondria in autism. In patients with established autism, certain tissues may exhibit mitochondrial dysfunction due to genetic or environmental factors. To compensate for this energy production shortfall, other tissues (such as blood cells) may increase mtDNA-CN to maintain normal physiological functions (65, 66). Furthermore, patients with autism may exhibit a chronic low-grade inflammatory state, where inflammation can induce mitochondrial biogenesis and mtDNA replication as part of the immune cell activation and cellular stress response (65–67). Additionally, cells may increase mtDNA-CN in response to stress (such as oxidative stress) to enhance their energy production capacity, aiming to protect cells from further damage (65, 66).

Our study also discovered that the occurrence of anxiety disorders leads to an increase in mtDNA-CN. Previous research supports our findings; Tymofiyeva et al. (33) found that salivary mtDNA-CN was associated with the severity of anxiety symptoms in adolescent samples. In another study, higher levels of blood leukocyte mtDNA-CN were associated with a lifetime history of anxiety disorders (34). Firstly, chronic stress, including induced glucocorticoids, pro-inflammatory cytokines, and oxidative stress activation, may damage mitochondria and stimulate new mitochondrial generation and DNA replication as a compensatory mechanism to maintain energy output (65, 66, 68–70). Meanwhile, chronic stress also stimulates the body’s HPA axis (hypothalamic-pituitary-adrenal axis), leading to elevated cortisol levels. Cortisol acts on coordinated physiological systems, inducing metabolic stress and affecting mitochondrial density and function by altering the expression of mitochondrial and nuclear genes that impact mitochondria. Glucocorticoid receptors bind to mitochondrial membranes to regulate membrane potential (71–74). Additionally, anxiety disorders may increase mtDNA-CN by affecting mitochondrial-related cellular signaling pathways, including AMPK (AMP-activated protein kinase) and sirtuins, which are key proteins active during cellular energy crises and stress (75–79).

Our bidirectional MR analysis did not identify any forward or reverse causal relationships between mtDNA-CN and other neuropsychiatric disorders (including AD, ADHD, AN, BD, MDD, OCD, SCZ, PTSD). Previous observational studies have explored the relationships between mtDNA-CN and AD (9, 10), ADHD (11–13), BD (18–21), MDD (18, 22–27), SCZ (28–32), and PTSD (35–37), with limited and highly controversial findings. Similar to our results, a community-based cohort meta-analysis using MR analysis did not reveal a causal relationship between blood mtDNA-CN and cognition (9). Other studies indicated a significant reduction in mtDNA-CN in the brains of AD patients (10). Three observational studies reported elevated mtDNA-CN in ADHD (11–13). A recent meta-analysis found no predictable relationship between BD diagnosis and mtDNA-CN, regardless of BD subtype; however, BD type 1 patients showed a decrease, while BD type 2 patients showed an increase in mtDNA-CN (18). Fries et al. (19) and Rollins et al. (20) found a significant increase in mtDNA-CN among BD patients compared to healthy controls, whereas de Sousa et al. (21) observed no significant difference in mtDNA-CN between BD patients and healthy controls. In a recent longitudinal study by Verhoeven et al. (22), no evidence of an association between depressive symptoms and mtDNA-CN was found in a community-based adult sample of individuals with depression. He et al. (23) also found no relationship between white blood cell mtDNA-CN and MDD in young people. However, some studies suggest a high level of mtDNA-CN is associated with depression (18, 24–26); in another study, a decrease in mtDNA-CN in adult MDD was observed (27). Li et al. (28) and Shivakumar et al. (29) reported a significant reduction in peripheral blood mtDNA-CN in patients with SCZ; some reported a decrease in mtDNA-CN in the brains of SCZ patients (30, 31); while others reported no differences (32). In Cai et al.’s study, Holocaust survivors with PTSD showed no differences in mitochondrial DNA copy number compared to healthy controls (35). Some studies have observed a reduction in mtDNA-CN in PTSD patients (36, 37). Our study is the first to use MR analysis to explore the bidirectional causal relationships between mtDNA-CN and other neuropsychiatric disorders (including AD, ADHD, AN, BD, MDD, OCD, SCZ, PTSD) from a genetic prediction standpoint, and our findings did not reveal any associations.

The current study has several strengths. Firstly, it included mtDNA-CN and 10 neuropsychiatric disorders, making it the first comprehensive MR study on the relationship between mtDNA-CN and neuropsychiatric disorders. Secondly, our conclusions are based on genetic instrumental variables and multiple MR analysis methods for causal inference. The results are robust, not confounded by horizontal pleiotropy or other factors. However, our study also has limitations. Firstly, due to the lack of individual information, we could not further stratify the population for analysis. Secondly, the study is based on European databases, therefore our conclusions cannot be extended to other ethnic groups, limiting the generalizability of our results. Future research could include populations with different characteristics (such as race and age) in MR studies may increase the representation of different populations. Lastly, we used more relaxed thresholds to assess results, which may increase some false positives while allowing a more comprehensive assessment of strong associations between mtDNA-CN and neuropsychiatric disorders.

## 5. Conclusion

In this study, potential causal relationships between mtDNA-CN and both ASD and anxiety disorders were established. Our findings indicate that a reduction in mtDNA-CN can increase the likelihood of developing ASD; the occurrence of anxiety disorders can lead to an increase in mtDNA-CN. These findings provide insights into the biological mechanisms of mtDNA-CN on ASD and underscore the significance of mtDNA-CN as a potential biomarker for anxiety disorders.

## Data Availability

The datasets used and/or analyzed during the current study are presented in the manuscript. Summary statistics for GWAS are publicly available.

## Ethical statement

Our analyses were based on publicly available data that had been approved by relevant review boards. Therefore, no additional ethical approval is required.

## Acknowledgment

We thank the participants and investigators of the UK Biobank, Psychiatric Genomics Consortium and FinnGen Consortium.

## Author’s contributions

All the contributing authors have participated in the preparation of the manuscript.

