## Supplemental Data 1 for "The role of mitochondrial DNA copy number in neuropsychiatric disorders: A bidirectional two-sample Mendelian randomization study"

**Additional file 1**

China

**Michael Maes Google Scholar profile**

Prof. Dr. Michael Maes, M.D., Ph.D.

https://scholar.google.co.th/citations?user=1wzMZ7UAAAAJ&hl=th&oi=ao

Highly cited author: 2003-2023 (ISI, Clarivate)

ScholarGPS: Worldwide #1 in molecular neuroscience; #1/4 in pathophysiology

Expert worldwide medical expertise ranking, Expertscape (December 2022), worldwide:

#1 in CFS, #1 in oxidative stress, #1 in encephalomyelitis, #1 in nitrosative stress, #1 in nitrosation,

#1 in tryptophan, #1 in aromatic amino acids, #1 in stress (physiological), #1 in neuroimmune;

#2 in bacterial translocation; #3 in inflammation, #4-5: in depression, fatigue and psychiatry.

Table S1. Genetic variants used as instrumental variables for mitochondrial DNA copy number.

Table S2. Genetic variants used as instrumental variables for the relationship between mitochondrial DNA copy number and Alzheimer's disease.

Table S3. Genetic variants used as instrumental variables for the relationship between mitochondrial DNA copy number and attention-deficit/hyperactivity disorder.

Table S4. Genetic variants used as instrumental variables for the relationship between mitochondrial DNA copy number and anorexia nervosa.

Table S5. Genetic variants used as instrumental variables for the relationship between mitochondrial DNA copy number and autism spectrum disorder.

Table S6. Genetic variants used as instrumental variables for the relationship between mitochondrial DNA copy number and bipolar disorder.

Table S7. Genetic variants used as instrumental variables for the relationship between mitochondrial DNA copy number and major depressive disorder.

Table S8. Genetic variants used as instrumental variables for the relationship between mitochondrial DNA copy number and obsessive compulsive disorder.

Table S9. Genetic variants used as instrumental variables for the relationship between mitochondrial DNA copy number and Schizophrenia.

Table S10. Genetic variants used as instrumental variables for the relationship between mitochondrial DNA copy number and anxiety disorders.

Table S11. Genetic variants used as instrumental variables for the relationship between mitochondrial DNA copy number and post-traumatic stress disorder.

Table S12. Genetic variants used as instrumental variables for the relationship between Alzheimer’s disease and mitochondrial DNA copy number.

Table S13. Genetic variants used as instrumental variables for the relationship between attention-deficit/hyperactivity disorder and mitochondrial DNA copy number.

Table S14. Genetic variants used as instrumental variables for the relationship between anorexia nervosa and mitochondrial DNA copy number.

Table S15. Genetic variants used as instrumental variables for the relationship between autism spectrum disorder and mitochondrial DNA copy number.

Table S16. Genetic variants used as instrumental variables for the relationship between bipolar disorder and mitochondrial DNA copy number.

Table S17. Genetic variants used as instrumental variables for the relationship between major depressive disorder and mitochondrial DNA copy number.

Table S18. Genetic variants used as instrumental variables for the relationship between obsessive compulsive disorder and mitochondrial DNA copy number.

Table S19. Genetic variants used as instrumental variables for the relationship between Schizophrenia and mitochondrial DNA copy number.

Table S20. Genetic variants used as instrumental variables for the relationship between anxiety disorders and mitochondrial DNA copy number.

Table S21. Genetic variants used as instrumental variables for the relationship between post-traumatic stress disorder and mitochondrial DNA copy number.

**[Figure S1](https://europepmc.org/articles/PMC9349767/figure/jmv28008-fig-0003/" \t "figure)** **The forward MR analyses: Plots of “leave-one-out” analyses for MR analyses of the causal effect of mtDNA copy number with the risk of neuropsychiatric disorders.** A Alzheimer’s disease, B attention-deficit/hyperactivity disorder, C anorexia nervosa, D autism spectrum disorder, E bipolar disorder, F major depressive disorder, G obsessive compulsive disorder, H Schizophrenia, I anxiety disorders, J post-traumatic stress disorder. The horizontal lines in the figure represents beta value and its 95% confidence interval [CI] of causal inference, which indicates the genetic effect of the SNP on neuropsychiatric disorders.

**[Figure S2](https://europepmc.org/articles/PMC9349767/figure/jmv28008-fig-0003/" \t "figure)** **The reverse MR analyses: Plots of “leave-one-out” analyses for MR analyses of the causal effect of neuropsychiatric disorders on mtDNA copy number .** A Alzheimer’s disease, B attention-deficit/hyperactivity disorder, C anorexia nervosa, D anxiety disorders, E autism spectrum disorder, F bipolar disorder, G major depressive disorder, H Schizophrenia. The horizontal lines in the figure represents beta value and its 95% confidence interval [CI] of causal inference, which indicates the genetic effect of the SNP on mtDNA copy number.

**Table S1. Genetic variants used as instrumental variables for mitochondrial DNA copy number.**

| **SNPs** | **Effect allele** | **Other allele** | **Beta** | **SE** | ***P* value** |
| --- | --- | --- | --- | --- | --- |
| rs1569419 | C | T | 0.0189 | 0.0025 | 6.87E-14 |
| rs182346769 | A | G | 0.0213 | 0.0027 | 7.87E-15 |
| rs2038479 | A | C | 0.0162 | 0.0027 | 1.47E-09 |
| rs10749636 | A | G | 0.0155 | 0.0025 | 5.43E-10 |
| rs2977608 | C | A | 0.0236 | 0.0025 | 2.58E-21 |
| rs72660908 | G | C | 0.0457 | 0.0021 | 8.24E-101 |
| rs2274319 | C | T | -0.0139 | 0.0022 | 4.09E-10 |
| rs143989240 | TTG | T | 0.0127 | 0.0023 | 2.61E-08 |
| rs4846082 | T | C | -0.0179 | 0.0021 | 3.33E-17 |
| rs62641680 | A | G | -0.0903 | 0.0063 | 4.63E-47 |
| rs12052715 | G | C | -0.0133 | 0.0024 | 1.72E-08 |
| rs74874677 | G | A | -0.0819 | 0.0071 | 3.62E-31 |
| rs78909033 | A | G | -0.021 | 0.0031 | 1.00E-11 |
| rs1354034 | C | T | -0.0268 | 0.0022 | 2.05E-35 |
| rs13084580 | T | C | 0.0244 | 0.0033 | 2.19E-13 |
| rs9844549 | G | A | 0.0171 | 0.0025 | 1.16E-11 |
| rs34778241 | TG | T | 0.014 | 0.0023 | 2.02E-09 |
| rs10013187 | A | C | -0.0121 | 0.0021 | 9.74E-09 |
| rs114694170 | C | T | 0.0331 | 0.0045 | 3.04E-13 |
| rs7705526 | A | C | 0.0178 | 0.0023 | 6.46E-15 |
| rs36009521 | T | TA | -0.0181 | 0.0031 | 6.27E-09 |
| rs9494142 | C | T | 0.018 | 0.0024 | 1.93E-13 |
| rs212938 | G | A | 0.0149 | 0.0025 | 3.21E-09 |
| rs5745587 | G | A | 0.021 | 0.0028 | 3.53E-14 |
| rs4720497 | A | G | 0.0164 | 0.0025 | 8.71E-11 |
| rs28851188 | A | C | -0.012 | 0.0021 | 1.92E-08 |
| rs445 | T | C | 0.0207 | 0.0036 | 8.85E-09 |
| rs6959832 | A | G | -0.021 | 0.0021 | 2.10E-23 |
| rs342292 | G | C | 0.0283 | 0.0021 | 1.41E-40 |
| rs77236693 | T | C | 0.036 | 0.0036 | 2.08E-23 |
| rs4284061 | A | T | -0.0186 | 0.0022 | 2.97E-17 |
| rs453301 | G | T | 0.0121 | 0.0021 | 1.09E-08 |
| rs2322718 | G | T | 0.0136 | 0.0021 | 1.53E-10 |
| rs385893 | C | T | 0.0153 | 0.0021 | 5.96E-13 |
| rs12247015 | G | A | 0.0337 | 0.0021 | 1.28E-55 |
| rs80140716 | C | A | -0.0252 | 0.004 | 2.18E-10 |
| rs138055405 | T | G | 0.0488 | 0.0082 | 2.65E-09 |
| rs73349121 | C | G | -0.1083 | 0.0081 | 1.08E-40 |
| rs7080386 | A | C | -0.0451 | 0.0021 | 9.58E-98 |
| rs10835226 | T | C | 0.0184 | 0.0023 | 4.83E-15 |
| rs4388979 | T | G | -0.0141 | 0.0021 | 4.47E-11 |
| rs11553699 | G | A | 0.0445 | 0.0032 | 1.45E-43 |
| rs11064881 | A | G | 0.0255 | 0.004 | 2.58E-10 |
| rs11064074 | T | C | 0.0197 | 0.0021 | 4.87E-20 |
| rs5012419 | G | A | 0.0247 | 0.0022 | 2.66E-29 |
| rs1127787 | A | G | -0.0159 | 0.0028 | 1.49E-08 |
| rs2015599 | A | G | 0.0123 | 0.0021 | 6.03E-09 |
| rs72698722 | T | C | -0.0192 | 0.0027 | 1.25E-12 |
| rs1760940 | C | A | 0.0263 | 0.0024 | 5.20E-27 |
| rs289713 | A | T | -0.0149 | 0.0027 | 4.14E-08 |
| rs17850455 | G | C | 0.091 | 0.0104 | 1.81E-18 |
| rs2290507 | A | G | 0.0223 | 0.0031 | 3.64E-13 |
| rs1967556 | G | T | 0.0175 | 0.0021 | 1.97E-16 |
| rs11082396 | C | T | 0.0258 | 0.0031 | 2.54E-16 |
| rs12604328 | C | T | -0.0172 | 0.0024 | 1.24E-12 |
| rs4807780 | C | T | -0.0191 | 0.0022 | 1.61E-17 |
| rs4808075 | C | T | -0.0274 | 0.0023 | 3.19E-32 |
| rs1065853 | T | G | 0.0388 | 0.0039 | 1.58E-23 |
| rs11085147 | T | C | 0.0756 | 0.0036 | 1.54E-95 |
| rs142158911 | A | G | 0.0192 | 0.0033 | 7.39E-09 |
| rs1613662 | A | G | -0.0167 | 0.0028 | 3.90E-09 |
| rs4814778 | C | G | -0.0297 | 0.0022 | 5.27E-40 |
| rs2263663 | T | C | 0.02 | 0.0024 | 1.97E-16 |
| rs156355 | C | T | 0.0214 | 0.0022 | 6.41E-23 |
| rs8121099 | C | A | 0.0221 | 0.0021 | 1.63E-25 |
| rs5759176 | T | C | 0.0317 | 0.0023 | 1.51E-44 |
| rs140522 | C | T | -0.0142 | 0.0022 | 2.84E-10 |

Abbreviations: SE, Standard Error; SNPs, single nucleotide polymorphisms.

Table S2. Genetic variants used as instrumental variables for the relationship between mitochondrial DNA copy number and Alzheimer's disease.

| **SNPs** | **EA 1** | **OA 1** | **EA 2** | **OA 2** | **Beta 1** | **Beta 2** | **SE 1** | **SE 2** | ***P* value 1** | ***P* value 2** | ***F*** |
| --- | --- | --- | --- | --- | --- | --- | --- | --- | --- | --- | --- |
| rs10013187 | A | C | A | C | -0.0121 | -0.000937754 | 0.0021 | 0.002159462 | 9.74E-09 | 0.664104281 | 33.19954649 |
| rs1065853 | T | G | T | G | 0.0388 | -0.155245111 | 0.0039 | 0.0220456 | 1.58E-23 | 1.90E-12 | 98.97698882 |
| rs10749636 | A | G | A | G | 0.0155 | -0.005829331 | 0.0025 | 0.002518806 | 5.43E-10 | 0.020649982 | 38.44 |
| rs10835226 | T | C | T | C | 0.0184 | -0.001335377 | 0.0023 | 0.002371953 | 4.83E-15 | 0.57344417 | 64 |
| rs11064074 | T | C | T | C | 0.0197 | -0.000429055 | 0.0021 | 0.005344709 | 4.87E-20 | 0.936017309 | 88.00226757 |
| rs11064881 | A | G | A | G | 0.0255 | -0.001802804 | 0.004 | 0.004211874 | 2.58E-10 | 0.668630053 | 40.640625 |
| rs11082396 | C | T | C | T | 0.0258 | 0.002764858 | 0.0031 | 0.003137629 | 2.54E-16 | 0.378213122 | 69.2653486 |
| rs11085147 | T | C | T | C | 0.0756 | 0.003349505 | 0.0036 | 0.004151406 | 1.54E-95 | 0.419760734 | 441 |
| rs1127787 | A | G | A | G | -0.0159 | -0.000818481 | 0.0028 | 0.002942955 | 1.49E-08 | 0.780923757 | 32.24617347 |
| rs114694170 | C | T | C | T | 0.0331 | 0.002399326 | 0.0045 | 0.004988337 | 3.04E-13 | 0.630525584 | 54.10419753 |
| rs11553699 | G | A | G | A | 0.0445 | -0.002587797 | 0.0032 | 0.007925689 | 1.45E-43 | 0.744040372 | 193.3837891 |
| rs12247015 | G | A | G | A | 0.0337 | 0.002213767 | 0.0021 | 0.002188108 | 1.28E-55 | 0.311668818 | 257.5260771 |
| rs12604328 | C | T | C | T | -0.0172 | 0.002001646 | 0.0024 | 0.002467364 | 1.24E-12 | 0.417222855 | 51.36111111 |
| rs13084580 | T | C | T | C | 0.0244 | 0.001377198 | 0.0033 | 0.003407286 | 2.19E-13 | 0.686071549 | 54.67033976 |
| rs1354034 | C | T | C | T | -0.0268 | 0.003464761 | 0.0022 | 0.002190075 | 2.05E-35 | 0.113642979 | 148.3966942 |
| rs138055405 | T | G | T | G | 0.0488 | -0.001992798 | 0.0082 | 0.008220173 | 2.65E-09 | 0.808448712 | 35.41701368 |
| rs140522 | C | T | C | T | -0.0142 | -5.16E-05 | 0.0022 | 0.002273739 | 2.84E-10 | 0.981886753 | 41.66115702 |
| rs142158911 | A | G | A | G | 0.0192 | -0.005274115 | 0.0033 | 0.0033879 | 7.39E-09 | 0.119529705 | 33.85123967 |
| rs156355 | C | T | C | T | 0.0214 | -0.003716981 | 0.0022 | 0.005283906 | 6.41E-23 | 0.4817733 | 94.61983471 |
| rs1569419 | C | T | C | T | 0.0189 | -0.002035939 | 0.0025 | 0.002550001 | 6.87E-14 | 0.424634213 | 57.1536 |
| rs1613662 | A | G | A | G | -0.0167 | 0.00465454 | 0.0028 | 0.003007611 | 3.90E-09 | 0.121721745 | 35.57270408 |
| rs1760940 | C | A | C | A | 0.0263 | -0.004617589 | 0.0024 | 0.002542005 | 5.20E-27 | 0.069291512 | 120.0850694 |
| rs182346769 | A | G | A | G | 0.0213 | -0.008635135 | 0.0027 | 0.011987158 | 7.87E-15 | 0.4713 | 62.2345679 |
| rs1967556 | G | T | G | T | 0.0175 | 0.001001994 | 0.0021 | 0.002183617 | 1.97E-16 | 0.646328088 | 69.44444444 |
| rs2015599 | A | G | A | G | 0.0123 | 0.002141367 | 0.0021 | 0.002168252 | 6.03E-09 | 0.32334841 | 34.30612245 |
| rs2038479 | A | C | A | C | 0.0162 | 0.001422367 | 0.0027 | 0.00274869 | 1.47E-09 | 0.604827499 | 36 |
| rs212938 | G | A | G | A | 0.0149 | -0.001882572 | 0.0025 | 0.002614793 | 3.21E-09 | 0.471543454 | 35.5216 |
| rs2263663 | T | C | T | C | 0.02 | -0.004122657 | 0.0024 | 0.002568036 | 1.97E-16 | 0.108411653 | 69.44444444 |
| rs2274319 | C | T | C | T | -0.0139 | 5.22E-05 | 0.0022 | 0.002289742 | 4.09E-10 | 0.981806355 | 39.91942149 |
| rs2290507 | A | G | A | G | 0.0223 | -0.006807708 | 0.0031 | 0.0032186 | 3.64E-13 | 0.03442016 | 51.7471384 |
| rs2322718 | G | T | G | T | 0.0136 | -0.001185548 | 0.0021 | 0.002167284 | 1.53E-10 | 0.584364888 | 41.94104308 |
| rs28851188 | A | C | A | C | -0.012 | 0.000156346 | 0.0021 | 0.002186164 | 1.92E-08 | 0.942986954 | 32.65306122 |
| rs2977608 | C | A | C | A | 0.0236 | -6.49E-05 | 0.0025 | 0.002429334 | 2.58E-21 | 0.978673528 | 89.1136 |
| rs385893 | C | T | C | T | 0.0153 | 0.003794919 | 0.0021 | 0.002157054 | 5.96E-13 | 0.078525389 | 53.08163265 |
| rs4388979 | T | G | T | G | -0.0141 | 0.000440289 | 0.0021 | 0.002186257 | 4.47E-11 | 0.840394156 | 45.08163265 |
| rs445 | T | C | T | C | 0.0207 | 0.00055757 | 0.0036 | 0.003404827 | 8.85E-09 | 0.86992107 | 33.0625 |
| rs453301 | G | T | G | T | 0.0121 | 0.001177576 | 0.0021 | 0.002163992 | 1.09E-08 | 0.586325467 | 33.19954649 |
| rs4720497 | A | G | A | G | 0.0164 | 0.002706647 | 0.0025 | 0.002628769 | 8.71E-11 | 0.303186034 | 43.0336 |
| rs4808075 | C | T | C | T | -0.0274 | -0.000878534 | 0.0023 | 0.002433072 | 3.19E-32 | 0.718039638 | 141.9206049 |
| rs4846082 | T | C | T | C | -0.0179 | 0.000180482 | 0.0021 | 0.002158299 | 3.33E-17 | 0.933356881 | 72.6553288 |
| rs5012419 | G | A | G | A | 0.0247 | 0.000250866 | 0.0022 | 0.002217016 | 2.66E-29 | 0.909907976 | 126.0516529 |
| rs5745587 | G | A | G | A | 0.021 | -0.004828362 | 0.0028 | 0.002682157 | 3.53E-14 | 0.071832434 | 56.25 |
| rs5759176 | T | C | T | C | 0.0317 | -0.002360551 | 0.0023 | 0.00226328 | 1.51E-44 | 0.296958632 | 189.9603025 |
| rs62641680 | A | G | A | G | -0.0903 | 0.007035333 | 0.0063 | 0.007843256 | 4.63E-47 | 0.369723558 | 205.4444444 |
| rs6959832 | A | G | A | G | -0.021 | -1.88E-05 | 0.0021 | 0.002163451 | 2.10E-23 | 0.993056091 | 100 |
| rs7080386 | A | C | A | C | -0.0451 | -0.003121113 | 0.0021 | 0.002185661 | 9.58E-98 | 0.153293333 | 461.2267574 |
| rs72698722 | T | C | T | C | -0.0192 | -0.001875311 | 0.0027 | 0.00279259 | 1.25E-12 | 0.501882292 | 50.56790123 |
| rs74874677 | G | A | G | A | -0.0819 | 0.015238347 | 0.0071 | 0.007474723 | 3.62E-31 | 0.041484922 | 133.061099 |
| rs7705526 | A | C | A | C | 0.0178 | -0.014494578 | 0.0023 | 0.005620796 | 6.46E-15 | 0.009916113 | 59.89413989 |
| rs77236693 | T | C | T | C | 0.036 | 0.000195347 | 0.0036 | 0.003860018 | 2.08E-23 | 0.959638021 | 100 |
| rs78909033 | A | G | A | G | -0.021 | 0.000313753 | 0.0031 | 0.00342341 | 1.00E-11 | 0.926976686 | 45.88969823 |
| rs80140716 | C | A | C | A | -0.0252 | -0.001479149 | 0.004 | 0.004441478 | 2.18E-10 | 0.739111088 | 39.69 |
| rs8121099 | C | A | C | A | 0.0221 | -0.001849254 | 0.0021 | 0.002164482 | 1.63E-25 | 0.392903858 | 110.7505669 |
| rs9494142 | C | T | C | T | 0.018 | -0.006243923 | 0.0024 | 0.002517316 | 1.93E-13 | 0.01312391 | 56.25 |
| rs9844549 | G | A | G | A | 0.0171 | -0.003535783 | 0.0025 | 0.00265798 | 1.16E-11 | 0.183435319 | 46.7856 |

Abbreviations: 1, mitochondrial DNA copy number; 2, Alzheimer's disease; SE, Standard Error; EA, Effect allele;OA, Other allele; SNPs, single nucleotide polymorphisms.

Table S3. Genetic variants used as instrumental variables for the relationship between mitochondrial DNA copy number and attention-deficit/hyperactivity disorder.

| **SNPs** | **EA 1** | **OA 1** | **EA 2** | **OA 2** | **Beta 1** | **Beta 2** | **SE 1** | **SE 2** | ***P* value 1** | ***P* value 2** | ***F*** |
| --- | --- | --- | --- | --- | --- | --- | --- | --- | --- | --- | --- |
| rs10013187 | A | C | A | C | -0.0121 | 0.0428966 | 0.0021 | 0.0133 | 9.74E-09 | 0.00130299 | 33.19937333 |
| rs1065853 | T | G | T | G | 0.0388 | 0.0255995 | 0.0039 | 0.0248 | 1.58E-23 | 0.3023 | 98.97647261 |
| rs10749636 | A | G | A | G | 0.0155 | -0.00810274 | 0.0025 | 0.0163 | 5.43E-10 | 0.619001 | 38.43979952 |
| rs10835226 | T | C | T | C | 0.0184 | -0.00739729 | 0.0023 | 0.0145 | 4.83E-15 | 0.6107 | 63.99966621 |
| rs11064074 | T | C | T | C | 0.0197 | 0.0167981 | 0.0021 | 0.0151 | 4.87E-20 | 0.2676 | 88.0018086 |
| rs11064881 | A | G | A | G | 0.0255 | -0.00160128 | 0.004 | 0.0274 | 2.58E-10 | 0.9524 | 40.64041304 |
| rs11082396 | C | T | C | T | 0.0258 | 0.0166986 | 0.0031 | 0.0196 | 2.54E-16 | 0.3943 | 69.26498735 |
| rs11085147 | T | C | T | C | 0.0756 | -0.00670241 | 0.0036 | 0.0225 | 1.54E-95 | 0.767201 | 440.9977 |
| rs1127787 | A | G | A | G | -0.0159 | -0.012599 | 0.0028 | 0.0181 | 1.49E-08 | 0.487401 | 32.24600529 |
| rs114694170 | C | T | C | T | 0.0331 | -0.0437968 | 0.0045 | 0.0311 | 3.04E-13 | 0.1597 | 54.10391535 |
| rs11553699 | G | A | G | A | 0.0445 | -0.0267004 | 0.0032 | 0.0246 | 1.45E-43 | 0.279 | 193.3827805 |
| rs12247015 | G | A | G | A | 0.0337 | 0.00709511 | 0.0021 | 0.0138 | 1.28E-55 | 0.6078 | 257.524734 |
| rs12604328 | C | T | C | T | -0.0172 | 0.00820356 | 0.0024 | 0.0159 | 1.24E-12 | 0.6056 | 51.36084324 |
| rs13084580 | T | C | T | C | 0.0244 | -0.0356997 | 0.0033 | 0.0218 | 2.19E-13 | 0.1012 | 54.67005463 |
| rs1354034 | C | T | C | T | -0.0268 | -0.0213015 | 0.0022 | 0.0137 | 2.05E-35 | 0.1206 | 148.3959203 |
| rs138055405 | T | G | T | G | 0.0488 | 0.0499038 | 0.0082 | 0.0567 | 2.65E-09 | 0.3787 | 35.41682897 |
| rs140522 | C | T | C | T | -0.0142 | -0.00479847 | 0.0022 | 0.0143 | 2.84E-10 | 0.735099 | 41.66093974 |
| rs142158911 | A | G | A | G | 0.0192 | 0.00679685 | 0.0033 | 0.0219 | 7.39E-09 | 0.7576 | 33.85106312 |
| rs156355 | C | T | C | T | 0.0214 | -0.00970278 | 0.0022 | 0.0161 | 6.41E-23 | 0.5478 | 94.61934123 |
| rs1569419 | C | T | C | T | 0.0189 | -0.00610135 | 0.0025 | 0.0188 | 6.87E-14 | 0.745799 | 57.15330192 |
| rs1613662 | A | G | A | G | -0.0167 | -0.0135007 | 0.0028 | 0.0179 | 3.90E-09 | 0.4517 | 35.57251855 |
| rs1760940 | C | A | C | A | 0.0263 | 0.0142003 | 0.0024 | 0.0204 | 5.20E-27 | 0.4847 | 120.0844431 |
| rs1967556 | G | T | G | T | 0.0175 | 0.0182964 | 0.0021 | 0.0135 | 1.97E-16 | 0.1758 | 69.44408226 |
| rs2015599 | A | G | A | G | 0.0123 | 0.001998 | 0.0021 | 0.0132 | 6.03E-09 | 0.8812 | 34.30594353 |
| rs2038479 | A | C | A | C | 0.0162 | 0.0176041 | 0.0027 | 0.017 | 1.47E-09 | 0.3011 | 35.99981224 |
| rs212938 | G | A | G | A | 0.0149 | -0.00699547 | 0.0025 | 0.0159 | 3.21E-09 | 0.6575 | 35.52141474 |
| rs2263663 | T | C | T | C | 0.02 | -0.0163022 | 0.0024 | 0.0155 | 1.97E-16 | 0.2916 | 69.44408226 |
| rs2274319 | C | T | C | T | -0.0139 | -0.00639948 | 0.0022 | 0.0142 | 4.09E-10 | 0.649901 | 39.91921329 |
| rs2290507 | A | G | A | G | 0.0223 | 0.0199987 | 0.0031 | 0.0202 | 3.64E-13 | 0.3221 | 51.74686851 |
| rs2322718 | G | T | G | T | 0.0136 | 0.0135007 | 0.0021 | 0.0133 | 1.53E-10 | 0.311 | 41.94082434 |
| rs28851188 | A | C | A | C | -0.012 | -0.00549507 | 0.0021 | 0.0132 | 1.92E-08 | 0.6783 | 32.65289092 |
| rs36009521 | T | TA | T | TA | -0.0181 | 0.0140015 | 0.0031 | 0.0192 | 6.27E-09 | 0.4674 | 34.0903529 |
| rs385893 | C | T | C | T | 0.0153 | 0.00920221 | 0.0021 | 0.0137 | 5.96E-13 | 0.501601 | 53.08135581 |
| rs4388979 | T | G | T | G | -0.0141 | 0.0105046 | 0.0021 | 0.0137 | 4.47E-11 | 0.4433 | 45.08139753 |
| rs445 | T | C | T | C | 0.0207 | -0.0226035 | 0.0036 | 0.0236 | 8.85E-09 | 0.3365 | 33.06232756 |
| rs453301 | G | T | G | T | 0.0121 | 0.023197 | 0.0021 | 0.0136 | 1.09E-08 | 0.0882999 | 33.19937333 |
| rs4720497 | A | G | A | G | 0.0164 | 0.0034042 | 0.0025 | 0.0156 | 8.71E-11 | 0.8286 | 43.03337556 |
| rs4808075 | C | T | C | T | -0.0274 | 0.000700245 | 0.0023 | 0.0147 | 3.19E-32 | 0.9605 | 141.9198647 |
| rs4846082 | T | C | T | C | -0.0179 | 0.00929665 | 0.0021 | 0.0136 | 3.33E-17 | 0.4926 | 72.65494987 |
| rs5012419 | G | A | G | A | 0.0247 | -0.00740253 | 0.0022 | 0.0137 | 2.66E-29 | 0.587801 | 126.0509955 |
| rs5745587 | G | A | G | A | 0.021 | -0.0473987 | 0.0028 | 0.0163 | 3.53E-14 | 0.00358699 | 56.24970663 |
| rs5759176 | T | C | T | C | 0.0317 | -0.026498 | 0.0023 | 0.0143 | 1.51E-44 | 0.0629303 | 189.9593117 |
| rs6959832 | A | G | A | G | -0.021 | -0.0214995 | 0.0021 | 0.0136 | 2.10E-23 | 0.1146 | 99.99947845 |
| rs7080386 | A | C | A | C | -0.0451 | -0.0170037 | 0.0021 | 0.0135 | 9.58E-98 | 0.2066 | 461.2243519 |
| rs72698722 | T | C | T | C | -0.0192 | 0.011296 | 0.0027 | 0.0166 | 1.25E-12 | 0.494899 | 50.5676375 |
| rs74874677 | G | A | G | A | -0.0819 | 0.0668024 | 0.0071 | 0.0494 | 3.62E-31 | 0.1767 | 133.060405 |
| rs77236693 | T | C | T | C | 0.036 | 0.0145043 | 0.0036 | 0.0233 | 2.08E-23 | 0.5334 | 99.99947845 |
| rs80140716 | C | A | C | A | -0.0252 | 0.0175023 | 0.004 | 0.0288 | 2.18E-10 | 0.5448 | 39.689793 |
| rs8121099 | C | A | C | A | 0.0221 | -0.0030952 | 0.0021 | 0.0132 | 1.63E-25 | 0.8169 | 110.7499893 |
| rs9494142 | C | T | C | T | 0.018 | -0.0291995 | 0.0024 | 0.0155 | 1.93E-13 | 0.0598301 | 56.24970663 |
| rs9844549 | G | A | G | A | 0.0171 | -0.001998 | 0.0025 | 0.0159 | 1.16E-11 | 0.8982 | 46.78535599 |

Abbreviations: 1, mitochondrial DNA copy number; 2, Attention-deficit/hyperactivity disorder; SE, Standard Error; EA, Effect allele;OA, Other allele; SNPs, single nucleotide polymorphisms.

Table S4. Genetic variants used as instrumental variables for the relationship between mitochondrial DNA copy number and anorexia nervosa.

| **SNPs** | **EA 1** | **OA 1** | **EA 2** | **OA 2** | **Beta 1** | **Beta 2** | **SE 1** | **SE 2** | ***P* value 1** | ***P* value 2** | ***F*** |
| --- | --- | --- | --- | --- | --- | --- | --- | --- | --- | --- | --- |
| rs10013187 | A | C | A | C | -0.0121 | 0.0188021 | 0.0021 | 0.0293 | 9.74E-09 | 0.5212 | 33.19937333 |
| rs1065853 | T | G | T | G | 0.0388 | -0.0378992 | 0.0039 | 0.0563 | 1.58E-23 | 0.5008 | 98.97647261 |
| rs10749636 | A | G | A | G | 0.0155 | 0.00639948 | 0.0025 | 0.035 | 5.43E-10 | 0.8555 | 38.43979952 |
| rs10835226 | T | C | T | C | 0.0184 | 0.0371031 | 0.0023 | 0.0316 | 4.83E-15 | 0.2407 | 63.99966621 |
| rs11064074 | T | C | T | C | 0.0197 | 0.0394995 | 0.0021 | 0.0304 | 4.87E-20 | 0.1935 | 88.0018086 |
| rs11064881 | A | G | A | G | 0.0255 | -0.129005 | 0.004 | 0.0567 | 2.58E-10 | 0.0227599 | 40.64041304 |
| rs11082396 | C | T | C | T | 0.0258 | -0.012699 | 0.0031 | 0.0435 | 2.54E-16 | 0.770999 | 69.26498735 |
| rs11085147 | T | C | T | C | 0.0756 | -0.0591027 | 0.0036 | 0.0524 | 1.54E-95 | 0.2594 | 440.9977 |
| rs1127787 | A | G | A | G | -0.0159 | -0.0390007 | 0.0028 | 0.0397 | 1.49E-08 | 0.326 | 32.24600529 |
| rs114694170 | C | T | C | T | 0.0331 | 0.0182047 | 0.0045 | 0.0685 | 3.04E-13 | 0.7906 | 54.10391535 |
| rs11553699 | G | A | G | A | 0.0445 | 0.0460963 | 0.0032 | 0.0453 | 1.45E-43 | 0.3094 | 193.3827805 |
| rs12247015 | G | A | G | A | 0.0337 | 0.0754027 | 0.0021 | 0.0296 | 1.28E-55 | 0.01079 | 257.524734 |
| rs12604328 | C | T | C | T | -0.0172 | 0.0258 | 0.0024 | 0.0327 | 1.24E-12 | 0.43 | 51.36084324 |
| rs13084580 | T | C | T | C | 0.0244 | 0.00039992 | 0.0033 | 0.0455 | 2.19E-13 | 0.9925 | 54.67005463 |
| rs1354034 | C | T | C | T | -0.0268 | -0.0456997 | 0.0022 | 0.0297 | 2.05E-35 | 0.1241 | 148.3959203 |
| rs138055405 | T | G | T | G | 0.0488 | -0.0594953 | 0.0082 | 0.113 | 2.65E-09 | 0.5985 | 35.41682897 |
| rs140522 | C | T | C | T | -0.0142 | -0.0683967 | 0.0022 | 0.0308 | 2.84E-10 | 0.0263299 | 41.66093974 |
| rs142158911 | A | G | A | G | 0.0192 | 0.0202045 | 0.0033 | 0.045 | 7.39E-09 | 0.653501 | 33.85106312 |
| rs156355 | C | T | C | T | 0.0214 | 0.0326988 | 0.0022 | 0.0309 | 6.41E-23 | 0.2913 | 94.61934123 |
| rs1569419 | C | T | C | T | 0.0189 | -0.0202045 | 0.0025 | 0.0341 | 6.87E-14 | 0.5525 | 57.15330192 |
| rs1613662 | A | G | A | G | -0.0167 | 0.0507977 | 0.0028 | 0.0398 | 3.90E-09 | 0.2017 | 35.57251855 |
| rs1760940 | C | A | C | A | 0.0263 | 0.037297 | 0.0024 | 0.0364 | 5.20E-27 | 0.3055 | 120.0844431 |
| rs1967556 | G | T | G | T | 0.0175 | 0.0117993 | 0.0021 | 0.0294 | 1.97E-16 | 0.6875 | 69.44408226 |
| rs2015599 | A | G | A | G | 0.0123 | -0.00969686 | 0.0021 | 0.0295 | 6.03E-09 | 0.742 | 34.30594353 |
| rs2038479 | A | C | A | C | 0.0162 | -0.0418016 | 0.0027 | 0.0369 | 1.47E-09 | 0.2574 | 35.99981224 |
| rs212938 | G | A | G | A | 0.0149 | -0.0399032 | 0.0025 | 0.0354 | 3.21E-09 | 0.2599 | 35.52141474 |
| rs2263663 | T | C | T | C | 0.02 | -0.0151035 | 0.0024 | 0.0334 | 1.97E-16 | 0.6517 | 69.44408226 |
| rs2274319 | C | T | C | T | -0.0139 | -0.0338022 | 0.0022 | 0.0306 | 4.09E-10 | 0.2692 | 39.91921329 |
| rs2290507 | A | G | A | G | 0.0223 | 0.0358979 | 0.0031 | 0.0435 | 3.64E-13 | 0.4091 | 51.74686851 |
| rs2322718 | G | T | G | T | 0.0136 | 0.0340016 | 0.0021 | 0.0292 | 1.53E-10 | 0.2446 | 41.94082434 |
| rs28851188 | A | C | A | C | -0.012 | -0.0191013 | 0.0021 | 0.0292 | 1.92E-08 | 0.5134 | 32.65289092 |
| rs2977608 | C | A | C | A | 0.0236 | -0.00560427 | 0.0025 | 0.0376 | 2.58E-21 | 0.8813 | 89.11313523 |
| rs34778241 | TG | T | TG | T | 0.014 | -0.0214973 | 0.0023 | 0.0335 | 2.02E-09 | 0.5223 | 37.05084646 |
| rs36009521 | T | TA | T | TA | -0.0181 | 0.0358014 | 0.0031 | 0.0421 | 6.27E-09 | 0.3948 | 34.0903529 |
| rs385893 | C | T | C | T | 0.0153 | -0.00850374 | 0.0021 | 0.0292 | 5.96E-13 | 0.771699 | 53.08135581 |
| rs4388979 | T | G | T | G | -0.0141 | -0.0121029 | 0.0021 | 0.0297 | 4.47E-11 | 0.6851 | 45.08139753 |
| rs445 | T | C | T | C | 0.0207 | 0.0530003 | 0.0036 | 0.0458 | 8.85E-09 | 0.2472 | 33.06232756 |
| rs453301 | G | T | G | T | 0.0121 | -0.0247024 | 0.0021 | 0.0293 | 1.09E-08 | 0.3995 | 33.19937333 |
| rs4720497 | A | G | A | G | 0.0164 | -0.0253999 | 0.0025 | 0.0356 | 8.71E-11 | 0.4762 | 43.03337556 |
| rs4808075 | C | T | C | T | -0.0274 | 0.0435027 | 0.0023 | 0.0317 | 3.19E-32 | 0.1708 | 141.9198647 |
| rs4846082 | T | C | T | C | -0.0179 | -0.0446 | 0.0021 | 0.0293 | 3.33E-17 | 0.1277 | 72.65494987 |
| rs5012419 | G | A | G | A | 0.0247 | -0.0115036 | 0.0022 | 0.0299 | 2.66E-29 | 0.7002 | 126.0509955 |
| rs5745587 | G | A | G | A | 0.021 | 0.074896 | 0.0028 | 0.0357 | 3.53E-14 | 0.0357396 | 56.24970663 |
| rs5759176 | T | C | T | C | 0.0317 | -0.010505 | 0.0023 | 0.0313 | 1.51E-44 | 0.737999 | 189.9593117 |
| rs62641680 | A | G | A | G | -0.0903 | -0.130895 | 0.0063 | 0.119 | 4.63E-47 | 0.2713 | 205.443373 |
| rs6959832 | A | G | A | G | -0.021 | -0.019295 | 0.0021 | 0.0292 | 2.10E-23 | 0.508 | 99.99947845 |
| rs7080386 | A | C | A | C | -0.0451 | 0.0178989 | 0.0021 | 0.0292 | 9.58E-98 | 0.5399 | 461.2243519 |
| rs72698722 | T | C | T | C | -0.0192 | -0.0208049 | 0.0027 | 0.0378 | 1.25E-12 | 0.5827 | 50.5676375 |
| rs74874677 | G | A | G | A | -0.0819 | -0.101202 | 0.0071 | 0.1122 | 3.62E-31 | 0.3671 | 133.060405 |
| rs7705526 | A | C | A | C | 0.0178 | -0.00020002 | 0.0023 | 0.033 | 6.46E-15 | 0.9947 | 59.89382751 |
| rs77236693 | T | C | T | C | 0.036 | 0.0221039 | 0.0036 | 0.0509 | 2.08E-23 | 0.6637 | 99.99947845 |
| rs78909033 | A | G | A | G | -0.021 | -0.0313049 | 0.0031 | 0.0488 | 1.00E-11 | 0.5213 | 45.8894589 |
| rs80140716 | C | A | C | A | -0.0252 | -0.0147999 | 0.004 | 0.0642 | 2.18E-10 | 0.8178 | 39.689793 |
| rs8121099 | C | A | C | A | 0.0221 | 0.00160128 | 0.0021 | 0.029 | 1.63E-25 | 0.957 | 110.7499893 |
| rs9494142 | C | T | C | T | 0.018 | -0.00760104 | 0.0024 | 0.0339 | 1.93E-13 | 0.8215 | 56.24970663 |
| rs9844549 | G | A | G | A | 0.0171 | 0.0154995 | 0.0025 | 0.034 | 1.16E-11 | 0.649101 | 46.78535599 |

Abbreviations: 1, mitochondrial DNA copy number; 2, Anorexia nervosa; SE, Standard Error; EA, Effect allele;OA, Other allele; SNPs, single nucleotide polymorphisms.

Table S5. Genetic variants used as instrumental variables for the relationship between mitochondrial DNA copy number and autism spectrum disorder.

| **SNPs** | **EA 1** | **OA 1** | **EA 2** | **OA 2** | **Beta 1** | **Beta 2** | **SE 1** | **SE 2** | ***P* value 1** | ***P* value 2** | ***F*** |
| --- | --- | --- | --- | --- | --- | --- | --- | --- | --- | --- | --- |
| rs10013187 | A | C | A | C | -0.0121 | 0.00600195 | 0.0021 | 0.014 | 9.74E-09 | 0.6664 | 33.19937333 |
| rs1065853 | T | G | T | G | 0.0388 | -0.0120017 | 0.0039 | 0.0255 | 1.58E-23 | 0.6367 | 98.97647261 |
| rs10749636 | A | G | A | G | 0.0155 | -0.016902 | 0.0025 | 0.0167 | 5.43E-10 | 0.3114 | 38.43979952 |
| rs10835226 | T | C | T | C | 0.0184 | -0.0160989 | 0.0023 | 0.0152 | 4.83E-15 | 0.2909 | 63.99966621 |
| rs11064074 | T | C | T | C | 0.0197 | 0.00840458 | 0.0021 | 0.0143 | 4.87E-20 | 0.5549 | 88.0018086 |
| rs11064881 | A | G | A | G | 0.0255 | -0.036498 | 0.004 | 0.0283 | 2.58E-10 | 0.1966 | 40.64041304 |
| rs11082396 | C | T | C | T | 0.0258 | -0.0240973 | 0.0031 | 0.021 | 2.54E-16 | 0.2495 | 69.26498735 |
| rs11085147 | T | C | T | C | 0.0756 | -0.0465048 | 0.0036 | 0.023 | 1.54E-95 | 0.0428697 | 440.9977 |
| rs1127787 | A | G | A | G | -0.0159 | -0.00830439 | 0.0028 | 0.0188 | 1.49E-08 | 0.658901 | 32.24600529 |
| rs114694170 | C | T | C | T | 0.0331 | -0.0924971 | 0.0045 | 0.0326 | 3.04E-13 | 0.00457004 | 54.10391535 |
| rs11553699 | G | A | G | A | 0.0445 | -0.0244975 | 0.0032 | 0.0223 | 1.45E-43 | 0.2704 | 193.3827805 |
| rs12247015 | G | A | G | A | 0.0337 | -0.0009995 | 0.0021 | 0.0143 | 1.28E-55 | 0.9434 | 257.524734 |
| rs12604328 | C | T | C | T | -0.0172 | 0.0153979 | 0.0024 | 0.0165 | 1.24E-12 | 0.3505 | 51.36084324 |
| rs13084580 | T | C | T | C | 0.0244 | -0.0181029 | 0.0033 | 0.0226 | 2.19E-13 | 0.4246 | 54.67005463 |
| rs1354034 | C | T | C | T | -0.0268 | -0.0077995 | 0.0022 | 0.0142 | 2.05E-35 | 0.5823 | 148.3959203 |
| rs138055405 | T | G | T | G | 0.0488 | 0.0189003 | 0.0082 | 0.0571 | 2.65E-09 | 0.7399 | 35.41682897 |
| rs140522 | C | T | C | T | -0.0142 | 0.00450011 | 0.0022 | 0.0148 | 2.84E-10 | 0.762901 | 41.66093974 |
| rs142158911 | A | G | A | G | 0.0192 | 0.0301023 | 0.0033 | 0.0222 | 7.39E-09 | 0.1751 | 33.85106312 |
| rs156355 | C | T | C | T | 0.0214 | -0.0231988 | 0.0022 | 0.0148 | 6.41E-23 | 0.1162 | 94.61934123 |
| rs1569419 | C | T | C | T | 0.0189 | 0.0203048 | 0.0025 | 0.0183 | 6.87E-14 | 0.268 | 57.15330192 |
| rs1613662 | A | G | A | G | -0.0167 | 0.0100988 | 0.0028 | 0.0186 | 3.90E-09 | 0.5871 | 35.57251855 |
| rs1760940 | C | A | C | A | 0.0263 | -0.0255995 | 0.0024 | 0.0181 | 5.20E-27 | 0.1573 | 120.0844431 |
| rs1967556 | G | T | G | T | 0.0175 | -0.0194006 | 0.0021 | 0.0139 | 1.97E-16 | 0.1631 | 69.44408226 |
| rs2015599 | A | G | A | G | 0.0123 | 0.00489799 | 0.0021 | 0.0139 | 6.03E-09 | 0.723801 | 34.30594353 |
| rs2038479 | A | C | A | C | 0.0162 | 0.0135971 | 0.0027 | 0.0179 | 1.47E-09 | 0.4479 | 35.99981224 |
| rs212938 | G | A | G | A | 0.0149 | -0.0214973 | 0.0025 | 0.0164 | 3.21E-09 | 0.1901 | 35.52141474 |
| rs2263663 | T | C | T | C | 0.02 | -0.0129029 | 0.0024 | 0.0159 | 1.97E-16 | 0.4175 | 69.44408226 |
| rs2274319 | C | T | C | T | -0.0139 | 0.00930314 | 0.0022 | 0.0146 | 4.09E-10 | 0.5248 | 39.91921329 |
| rs2290507 | A | G | A | G | 0.0223 | 0.0184979 | 0.0031 | 0.0216 | 3.64E-13 | 0.3915 | 51.74686851 |
| rs2322718 | G | T | G | T | 0.0136 | 0.0037972 | 0.0021 | 0.0139 | 1.53E-10 | 0.783201 | 41.94082434 |
| rs28851188 | A | C | A | C | -0.012 | -0.00700447 | 0.0021 | 0.0139 | 1.92E-08 | 0.611599 | 32.65289092 |
| rs2977608 | C | A | C | A | 0.0236 | -0.0125015 | 0.0025 | 0.0178 | 2.58E-21 | 0.484299 | 89.11313523 |
| rs34778241 | TG | T | TG | T | 0.014 | -0.025697 | 0.0023 | 0.0156 | 2.02E-09 | 0.1005 | 37.05084646 |
| rs36009521 | T | TA | T | TA | -0.0181 | 0.00139902 | 0.0031 | 0.0203 | 6.27E-09 | 0.944 | 34.0903529 |
| rs385893 | C | T | C | T | 0.0153 | -0.0150955 | 0.0021 | 0.0138 | 5.96E-13 | 0.2725 | 53.08135581 |
| rs4388979 | T | G | T | G | -0.0141 | -0.00270365 | 0.0021 | 0.0141 | 4.47E-11 | 0.851 | 45.08139753 |
| rs445 | T | C | T | C | 0.0207 | -0.00110061 | 0.0036 | 0.0232 | 8.85E-09 | 0.9613 | 33.06232756 |
| rs453301 | G | T | G | T | 0.0121 | -0.0582972 | 0.0021 | 0.0139 | 1.09E-08 | 2.88E-05 | 33.19937333 |
| rs4720497 | A | G | A | G | 0.0164 | -0.0251024 | 0.0025 | 0.0162 | 8.71E-11 | 0.1221 | 43.03337556 |
| rs4808075 | C | T | C | T | -0.0274 | 0.0246001 | 0.0023 | 0.015 | 3.19E-32 | 0.1017 | 141.9198647 |
| rs4846082 | T | C | T | C | -0.0179 | -0.00719583 | 0.0021 | 0.0141 | 3.33E-17 | 0.609399 | 72.65494987 |
| rs5012419 | G | A | G | A | 0.0247 | 0.00329542 | 0.0022 | 0.0144 | 2.66E-29 | 0.8211 | 126.0509955 |
| rs5745587 | G | A | G | A | 0.021 | 0.000100005 | 0.0028 | 0.017 | 3.53E-14 | 0.9958 | 56.24970663 |
| rs5759176 | T | C | T | C | 0.0317 | -0.0101008 | 0.0023 | 0.015 | 1.51E-44 | 0.4987 | 189.9593117 |
| rs62641680 | A | G | A | G | -0.0903 | -0.0872041 | 0.0063 | 0.0487 | 4.63E-47 | 0.0736495 | 205.443373 |
| rs6959832 | A | G | A | G | -0.021 | -0.0247026 | 0.0021 | 0.014 | 2.10E-23 | 0.0770992 | 99.99947845 |
| rs7080386 | A | C | A | C | -0.0451 | 0.0108015 | 0.0021 | 0.014 | 9.58E-98 | 0.4416 | 461.2243519 |
| rs72698722 | T | C | T | C | -0.0192 | 0.00819632 | 0.0027 | 0.0176 | 1.25E-12 | 0.641599 | 50.5676375 |
| rs74874677 | G | A | G | A | -0.0819 | 0.0226956 | 0.0071 | 0.0505 | 3.62E-31 | 0.6529 | 133.060405 |
| rs7705526 | A | C | A | C | 0.0178 | 0.0219963 | 0.0023 | 0.0158 | 6.46E-15 | 0.1641 | 59.89382751 |
| rs77236693 | T | C | T | C | 0.036 | -0.010505 | 0.0036 | 0.0241 | 2.08E-23 | 0.6629 | 99.99947845 |
| rs78909033 | A | G | A | G | -0.021 | 0.00410158 | 0.0031 | 0.0223 | 1.00E-11 | 0.8536 | 45.8894589 |
| rs80140716 | C | A | C | A | -0.0252 | -0.0178989 | 0.004 | 0.03 | 2.18E-10 | 0.5508 | 39.689793 |
| rs8121099 | C | A | C | A | 0.0221 | -0.0219963 | 0.0021 | 0.0138 | 1.63E-25 | 0.112 | 110.7499893 |
| rs9494142 | C | T | C | T | 0.018 | -0.00990083 | 0.0024 | 0.0164 | 1.93E-13 | 0.5438 | 56.24970663 |
| rs9844549 | G | A | G | A | 0.0171 | 0.0112025 | 0.0025 | 0.0163 | 1.16E-11 | 0.491501 | 46.78535599 |

Abbreviations: 1, mitochondrial DNA copy number; 2, autism spectrum disorder; SE, Standard Error; EA, Effect allele;OA, Other allele; SNPs, single nucleotide polymorphisms.

Table S6. Genetic variants used as instrumental variables for the relationship between mitochondrial DNA copy number and bipolar disorder.

| **SNPs** | **EA 1** | **OA 1** | **EA 2** | **OA 2** | **Beta 1** | **Beta 2** | **SE 1** | **SE 2** | ***P* value 1** | ***P* value 2** | ***F*** |
| --- | --- | --- | --- | --- | --- | --- | --- | --- | --- | --- | --- |
| rs10013187 | A | C | A | C | -0.0121 | 0.00559563 | 0.0021 | 0.0094 | 9.74E-09 | 0.555199 | 33.19937333 |
| rs10749636 | A | G | A | G | 0.0155 | 0.0166986 | 0.0025 | 0.0109 | 5.43E-10 | 0.1261 | 38.43979952 |
| rs10835226 | T | C | T | C | 0.0184 | 0.00559563 | 0.0023 | 0.0105 | 4.83E-15 | 0.595701 | 63.99966621 |
| rs11064074 | T | C | T | C | 0.0197 | -0.00539542 | 0.0021 | 0.0097 | 4.87E-20 | 0.575999 | 88.0018086 |
| rs11064881 | A | G | A | G | 0.0255 | -0.0343048 | 0.004 | 0.0186 | 2.58E-10 | 0.0648799 | 40.64041304 |
| rs11082396 | C | T | C | T | 0.0258 | 0.0226956 | 0.0031 | 0.0137 | 2.54E-16 | 0.0966696 | 69.26498735 |
| rs11085147 | T | C | T | C | 0.0756 | 0.0300984 | 0.0036 | 0.0164 | 1.54E-95 | 0.0671707 | 440.9977 |
| rs1127787 | A | G | A | G | -0.0159 | -0.0128965 | 0.0028 | 0.0126 | 1.49E-08 | 0.3076 | 32.24600529 |
| rs114694170 | C | T | C | T | 0.0331 | -0.012699 | 0.0045 | 0.0204 | 3.04E-13 | 0.536 | 54.10391535 |
| rs11553699 | G | A | G | A | 0.0445 | 0.0136021 | 0.0032 | 0.0153 | 1.45E-43 | 0.3756 | 193.3827805 |
| rs12247015 | G | A | G | A | 0.0337 | -0.0168964 | 0.0021 | 0.0095 | 1.28E-55 | 0.0766302 | 257.524734 |
| rs12604328 | C | T | C | T | -0.0172 | 0.0037972 | 0.0024 | 0.0111 | 1.24E-12 | 0.7335 | 51.36084324 |
| rs13084580 | T | C | T | C | 0.0244 | 0.00690378 | 0.0033 | 0.0157 | 2.19E-13 | 0.658901 | 54.67005463 |
| rs1354034 | C | T | C | T | -0.0268 | -0.0230034 | 0.0022 | 0.0098 | 2.05E-35 | 0.0181401 | 148.3959203 |
| rs138055405 | T | G | T | G | 0.0488 | 0.0226956 | 0.0082 | 0.0392 | 2.65E-09 | 0.562 | 35.41682897 |
| rs140522 | C | T | C | T | -0.0142 | -0.0030952 | 0.0022 | 0.0102 | 2.84E-10 | 0.7625 | 41.66093974 |
| rs142158911 | A | G | A | G | 0.0192 | -0.0193025 | 0.0033 | 0.0157 | 7.39E-09 | 0.2182 | 33.85106312 |
| rs156355 | C | T | C | T | 0.0214 | -0.00380276 | 0.0022 | 0.0099 | 6.41E-23 | 0.697301 | 94.61934123 |
| rs1569419 | C | T | C | T | 0.0189 | 0.00729656 | 0.0025 | 0.0119 | 6.87E-14 | 0.5396 | 57.15330192 |
| rs1613662 | A | G | A | G | -0.0167 | -0.018704 | 0.0028 | 0.0127 | 3.90E-09 | 0.1406 | 35.57251855 |
| rs1760940 | C | A | C | A | 0.0263 | -0.00819632 | 0.0024 | 0.0116 | 5.20E-27 | 0.4777 | 120.0844431 |
| rs182346769 | A | G | A | G | 0.0213 | -0.00299551 | 0.0027 | 0.0135 | 7.87E-15 | 0.8244 | 62.23424332 |
| rs1967556 | G | T | G | T | 0.0175 | 0.0204987 | 0.0021 | 0.0099 | 1.97E-16 | 0.0372598 | 69.44408226 |
| rs2015599 | A | G | A | G | 0.0123 | -0.0117013 | 0.0021 | 0.0094 | 6.03E-09 | 0.2133 | 34.30594353 |
| rs2038479 | A | C | A | C | 0.0162 | -0.0009995 | 0.0027 | 0.0118 | 1.47E-09 | 0.932 | 35.99981224 |
| rs212938 | G | A | G | A | 0.0149 | 0.00300451 | 0.0025 | 0.0113 | 3.21E-09 | 0.7867 | 35.52141474 |
| rs2263663 | T | C | T | C | 0.02 | -0.0172995 | 0.0024 | 0.0108 | 1.97E-16 | 0.1109 | 69.44408226 |
| rs2274319 | C | T | C | T | -0.0139 | -0.0154993 | 0.0022 | 0.0098 | 4.09E-10 | 0.1144 | 39.91921329 |
| rs2290507 | A | G | A | G | 0.0223 | 0.00920221 | 0.0031 | 0.0141 | 3.64E-13 | 0.5125 | 51.74686851 |
| rs2322718 | G | T | G | T | 0.0136 | 0.000100005 | 0.0021 | 0.0094 | 1.53E-10 | 0.9951 | 41.94082434 |
| rs28851188 | A | C | A | C | -0.012 | -0.0036035 | 0.0021 | 0.0094 | 1.92E-08 | 0.7032 | 32.65289092 |
| rs2977608 | C | A | C | A | 0.0236 | 0.00150113 | 0.0025 | 0.0131 | 2.58E-21 | 0.9084 | 89.11313523 |
| rs4388979 | T | G | T | G | -0.0141 | 0.00520351 | 0.0021 | 0.0096 | 4.47E-11 | 0.5836 | 45.08139753 |
| rs453301 | G | T | G | T | 0.0121 | 0.032203 | 0.0021 | 0.0094 | 1.09E-08 | 0.000640501 | 33.19937333 |
| rs4720497 | A | G | A | G | 0.0164 | -0.0382972 | 0.0025 | 0.0112 | 8.71E-11 | 0.000629695 | 43.03337556 |
| rs4807780 | C | T | C | T | -0.0191 | 0.00849599 | 0.0022 | 0.0105 | 1.61E-17 | 0.4211 | 75.37357383 |
| rs4808075 | C | T | C | T | -0.0274 | 0.012599 | 0.0023 | 0.0102 | 3.19E-32 | 0.2193 | 141.9198647 |
| rs4846082 | T | C | T | C | -0.0179 | 0.00490199 | 0.0021 | 0.0095 | 3.33E-17 | 0.606301 | 72.65494987 |
| rs5012419 | G | A | G | A | 0.0247 | 0.0111014 | 0.0022 | 0.0097 | 2.66E-29 | 0.2524 | 126.0509955 |
| rs5745587 | G | A | G | A | 0.021 | 0.0222967 | 0.0028 | 0.0116 | 3.53E-14 | 0.0551404 | 56.24970663 |
| rs5759176 | T | C | T | C | 0.0317 | 0.00300451 | 0.0023 | 0.01 | 1.51E-44 | 0.7673 | 189.9593117 |
| rs62641680 | A | G | A | G | -0.0903 | -0.00390238 | 0.0063 | 0.0332 | 4.63E-47 | 0.9056 | 205.443373 |
| rs6959832 | A | G | A | G | -0.021 | 0.0182047 | 0.0021 | 0.0095 | 2.10E-23 | 0.05466 | 99.99947845 |
| rs7080386 | A | C | A | C | -0.0451 | -0.0189984 | 0.0021 | 0.0095 | 9.58E-98 | 0.0444304 | 461.2243519 |
| rs72698722 | T | C | T | C | -0.0192 | 0.00830439 | 0.0027 | 0.0118 | 1.25E-12 | 0.4802 | 50.5676375 |
| rs74874677 | G | A | G | A | -0.0819 | 0.00359646 | 0.0071 | 0.0337 | 3.62E-31 | 0.9155 | 133.060405 |
| rs7705526 | A | C | A | C | 0.0178 | -0.00970278 | 0.0023 | 0.0105 | 6.46E-15 | 0.3568 | 59.89382751 |
| rs77236693 | T | C | T | C | 0.036 | 0.0127003 | 0.0036 | 0.016 | 2.08E-23 | 0.4269 | 99.99947845 |
| rs78909033 | A | G | A | G | -0.021 | 0.00150113 | 0.0031 | 0.0144 | 1.00E-11 | 0.9167 | 45.8894589 |
| rs80140716 | C | A | C | A | -0.0252 | 0.00879859 | 0.004 | 0.0182 | 2.18E-10 | 0.6264 | 39.689793 |
| rs8121099 | C | A | C | A | 0.0221 | 0.00210221 | 0.0021 | 0.0094 | 1.63E-25 | 0.8226 | 110.7499893 |
| rs9494142 | C | T | C | T | 0.018 | 0.00999983 | 0.0024 | 0.0108 | 1.93E-13 | 0.3554 | 56.24970663 |
| rs9844549 | G | A | G | A | 0.0171 | -0.0227981 | 0.0025 | 0.0111 | 1.16E-11 | 0.0398098 | 46.78535599 |

Abbreviations: 1, mitochondrial DNA copy number; 2, bipolar disorder; SE, Standard Error; EA, Effect allele;OA, Other allele; SNPs, single nucleotide polymorphisms.

Table S7. Genetic variants used as instrumental variables for the relationship between mitochondrial DNA copy number and major depressive disorder.

| **SNPs** | **EA 1** | **OA 1** | **EA 2** | **OA 2** | **Beta 1** | **Beta 2** | **SE 1** | **SE 2** | ***P* value 1** | ***P* value 2** | ***F*** |
| --- | --- | --- | --- | --- | --- | --- | --- | --- | --- | --- | --- |
| rs10013187 | A | C | A | C | -0.0121 | 0.0165031 | 0.0021 | 0.0079 | 9.74E-09 | 0.0374197 | 33.19937333 |
| rs10749636 | A | G | A | G | 0.0155 | 1.00E-04 | 0.0025 | 0.0095 | 5.43E-10 | 0.9954 | 38.43979952 |
| rs10835226 | T | C | T | C | 0.0184 | -0.00280393 | 0.0023 | 0.0087 | 4.83E-15 | 0.7471 | 63.99966621 |
| rs11064074 | T | C | T | C | 0.0197 | 0.0077995 | 0.0021 | 0.0082 | 4.87E-20 | 0.3405 | 88.0018086 |
| rs11064881 | A | G | A | G | 0.0255 | 0.0148985 | 0.004 | 0.0154 | 2.58E-10 | 0.3332 | 40.64041304 |
| rs11082396 | C | T | C | T | 0.0258 | 0.0034961 | 0.0031 | 0.0117 | 2.54E-16 | 0.7671 | 69.26498735 |
| rs11085147 | T | C | T | C | 0.0756 | -0.00579677 | 0.0036 | 0.0139 | 1.54E-95 | 0.677901 | 440.9977 |
| rs1127787 | A | G | A | G | -0.0159 | -0.00310482 | 0.0028 | 0.0107 | 1.49E-08 | 0.7742 | 32.24600529 |
| rs114694170 | C | T | C | T | 0.0331 | -0.0291995 | 0.0045 | 0.0181 | 3.04E-13 | 0.1061 | 54.10391535 |
| rs11553699 | G | A | G | A | 0.0445 | 0.00530404 | 0.0032 | 0.0138 | 1.45E-43 | 0.7014 | 193.3827805 |
| rs12247015 | G | A | G | A | 0.0337 | -0.00430074 | 0.0021 | 0.0081 | 1.28E-55 | 0.5995 | 257.524734 |
| rs12604328 | C | T | C | T | -0.0172 | 0.00800193 | 0.0024 | 0.0092 | 1.24E-12 | 0.3859 | 51.36084324 |
| rs13084580 | T | C | T | C | 0.0244 | -0.00480151 | 0.0033 | 0.0127 | 2.19E-13 | 0.7035 | 54.67005463 |
| rs1354034 | C | T | C | T | -0.0268 | 0.0120017 | 0.0022 | 0.0081 | 2.05E-35 | 0.1384 | 148.3959203 |
| rs138055405 | T | G | T | G | 0.0488 | 0.0789036 | 0.0082 | 0.0369 | 2.65E-09 | 0.0323601 | 35.41682897 |
| rs140522 | C | T | C | T | -0.0142 | -0.00600195 | 0.0022 | 0.009 | 2.84E-10 | 0.5011 | 41.66093974 |
| rs142158911 | A | G | A | G | 0.0192 | -0.0112025 | 0.0033 | 0.0135 | 7.39E-09 | 0.4049 | 33.85106312 |
| rs156355 | C | T | C | T | 0.0214 | -0.00269636 | 0.0022 | 0.0086 | 6.41E-23 | 0.754699 | 94.61934123 |
| rs1569419 | C | T | C | T | 0.0189 | 0.00579677 | 0.0025 | 0.0108 | 6.87E-14 | 0.5903 | 57.15330192 |
| rs1613662 | A | G | A | G | -0.0167 | -0.00240288 | 0.0028 | 0.0107 | 3.90E-09 | 0.821 | 35.57251855 |
| rs1760940 | C | A | C | A | 0.0263 | 0.0129029 | 0.0024 | 0.0099 | 5.20E-27 | 0.1928 | 120.0844431 |
| rs1967556 | G | T | G | T | 0.0175 | 0.0151035 | 0.0021 | 0.008 | 1.97E-16 | 0.0596705 | 69.44408226 |
| rs2015599 | A | G | A | G | 0.0123 | 0.0082955 | 0.0021 | 0.0079 | 6.03E-09 | 0.2926 | 34.30594353 |
| rs2038479 | A | C | A | C | 0.0162 | 0.025502 | 0.0027 | 0.0101 | 1.47E-09 | 0.0113201 | 35.99981224 |
| rs212938 | G | A | G | A | 0.0149 | 0.00540458 | 0.0025 | 0.0095 | 3.21E-09 | 0.568 | 35.52141474 |
| rs2263663 | T | C | T | C | 0.02 | -0.0217958 | 0.0024 | 0.0091 | 1.97E-16 | 0.01679 | 69.44408226 |
| rs2274319 | C | T | C | T | -0.0139 | 0.00549507 | 0.0022 | 0.0086 | 4.09E-10 | 0.5251 | 39.91921329 |
| rs2290507 | A | G | A | G | 0.0223 | -0.002002 | 0.0031 | 0.0136 | 3.64E-13 | 0.8823 | 51.74686851 |
| rs2322718 | G | T | G | T | 0.0136 | 0.00130085 | 0.0021 | 0.0079 | 1.53E-10 | 0.8649 | 41.94082434 |
| rs28851188 | A | C | A | C | -0.012 | -0.00979784 | 0.0021 | 0.008 | 1.92E-08 | 0.218 | 32.65289092 |
| rs2977608 | C | A | C | A | 0.0236 | -0.0129952 | 0.0025 | 0.0117 | 2.58E-21 | 0.2667 | 89.11313523 |
| rs385893 | C | T | C | T | 0.0153 | -0.0208021 | 0.0021 | 0.008 | 5.96E-13 | 0.00939896 | 53.08135581 |
| rs4388979 | T | G | T | G | -0.0141 | -0.00619918 | 0.0021 | 0.0081 | 4.47E-11 | 0.4458 | 45.08139753 |
| rs445 | T | C | T | C | 0.0207 | 0.00919757 | 0.0036 | 0.0132 | 8.85E-09 | 0.487401 | 33.06232756 |
| rs453301 | G | T | G | T | 0.0121 | -0.00299551 | 0.0021 | 0.008 | 1.09E-08 | 0.7021 | 33.19937333 |
| rs4720497 | A | G | A | G | 0.0164 | 0.00509699 | 0.0025 | 0.0096 | 8.71E-11 | 0.5945 | 43.03337556 |
| rs4808075 | C | T | C | T | -0.0274 | 0.00859685 | 0.0023 | 0.0091 | 3.19E-32 | 0.3456 | 141.9198647 |
| rs4846082 | T | C | T | C | -0.0179 | 0.0195967 | 0.0021 | 0.0081 | 3.33E-17 | 0.01562 | 72.65494987 |
| rs5012419 | G | A | G | A | 0.0247 | -0.00630011 | 0.0022 | 0.0082 | 2.66E-29 | 0.4429 | 126.0509955 |
| rs5745587 | G | A | G | A | 0.021 | -0.0082955 | 0.0028 | 0.01 | 3.53E-14 | 0.4092 | 56.24970663 |
| rs5759176 | T | C | T | C | 0.0317 | -0.014596 | 0.0023 | 0.01 | 1.51E-44 | 0.1461 | 189.9593117 |
| rs62641680 | A | G | A | G | -0.0903 | -0.0275048 | 0.0063 | 0.0271 | 4.63E-47 | 0.3103 | 205.443373 |
| rs6959832 | A | G | A | G | -0.021 | -0.0102019 | 0.0021 | 0.008 | 2.10E-23 | 0.2015 | 99.99947845 |
| rs7080386 | A | C | A | C | -0.0451 | -0.0122042 | 0.0021 | 0.008 | 9.58E-98 | 0.1277 | 461.2243519 |
| rs72698722 | T | C | T | C | -0.0192 | -0.00399798 | 0.0027 | 0.0102 | 1.25E-12 | 0.695401 | 50.5676375 |
| rs74874677 | G | A | G | A | -0.0819 | -0.0574006 | 0.0071 | 0.0283 | 3.62E-31 | 0.0426599 | 133.060405 |
| rs7705526 | A | C | A | C | 0.0178 | 0.0235017 | 0.0023 | 0.0096 | 6.46E-15 | 0.01414 | 59.89382751 |
| rs77236693 | T | C | T | C | 0.036 | 0.00269636 | 0.0036 | 0.0137 | 2.08E-23 | 0.8451 | 99.99947845 |
| rs78909033 | A | G | A | G | -0.021 | -0.00690378 | 0.0031 | 0.0129 | 1.00E-11 | 0.5904 | 45.8894589 |
| rs80140716 | C | A | C | A | -0.0252 | -0.0030952 | 0.004 | 0.0172 | 2.18E-10 | 0.8554 | 39.689793 |
| rs8121099 | C | A | C | A | 0.0221 | 0.00280393 | 0.0021 | 0.0081 | 1.63E-25 | 0.7256 | 110.7499893 |
| rs9494142 | C | T | C | T | 0.018 | 0.00470103 | 0.0024 | 0.0092 | 1.93E-13 | 0.6108 | 56.24970663 |
| rs9844549 | G | A | G | A | 0.0171 | 0.00579677 | 0.0025 | 0.0095 | 1.16E-11 | 0.539499 | 46.78535599 |

Abbreviations: 1, mitochondrial DNA copy number; 2, major depressive disorder; SE, Standard Error; EA, Effect allele;OA, Other allele; SNPs, single nucleotide polymorphisms.

Table S8. Genetic variants used as instrumental variables for the relationship between mitochondrial DNA copy number and obsessive compulsive disorder.

| **SNPs** | **EA 1** | **OA 1** | **EA 2** | **OA 2** | **Beta 1** | **Beta 2** | **SE 1** | **SE 2** | ***P* value 1** | ***P* value 2** | ***F*** |
| --- | --- | --- | --- | --- | --- | --- | --- | --- | --- | --- | --- |
| rs10013187 | A | C | A | C | -0.0121 | -0.00369984 | 0.0021 | 0.0339 | 9.74E-09 | 0.9128 | 33.19937333 |
| rs10749636 | A | G | A | G | 0.0155 | -0.0151004 | 0.0025 | 0.0398 | 5.43E-10 | 0.704301 | 38.43979952 |
| rs10835226 | T | C | T | C | 0.0184 | 0.0321961 | 0.0023 | 0.0376 | 4.83E-15 | 0.3911 | 63.99966621 |
| rs11064074 | T | C | T | C | 0.0197 | 0.0291995 | 0.0021 | 0.0351 | 4.87E-20 | 0.4054 | 88.0018086 |
| rs11064881 | A | G | A | G | 0.0255 | -0.0763002 | 0.004 | 0.0673 | 2.58E-10 | 0.2571 | 40.64041304 |
| rs11082396 | C | T | C | T | 0.0258 | 0.00599996 | 0.0031 | 0.0504 | 2.54E-16 | 0.905 | 69.26498735 |
| rs11085147 | T | C | T | C | 0.0756 | -0.0142003 | 0.0036 | 0.0625 | 1.54E-95 | 0.8201 | 440.9977 |
| rs1127787 | A | G | A | G | -0.0159 | 0.0133998 | 0.0028 | 0.046 | 1.49E-08 | 0.770699 | 32.24600529 |
| rs114694170 | C | T | C | T | 0.0331 | 0.0306005 | 0.0045 | 0.0846 | 3.04E-13 | 0.7173 | 54.10391535 |
| rs11553699 | G | A | G | A | 0.0445 | 0.0131004 | 0.0032 | 0.0592 | 1.45E-43 | 0.8243 | 193.3827805 |
| rs12247015 | G | A | G | A | 0.0337 | 0.0612001 | 0.0021 | 0.0338 | 1.28E-55 | 0.0702296 | 257.524734 |
| rs12604328 | C | T | C | T | -0.0172 | -0.0036035 | 0.0024 | 0.0376 | 1.24E-12 | 0.9237 | 51.36084324 |
| rs13084580 | T | C | T | C | 0.0244 | 0.0582029 | 0.0033 | 0.0528 | 2.19E-13 | 0.2703 | 54.67005463 |
| rs1354034 | C | T | C | T | -0.0268 | -0.00499749 | 0.0022 | 0.0343 | 2.05E-35 | 0.8842 | 148.3959203 |
| rs138055405 | T | G | T | G | 0.0488 | -0.0435998 | 0.0082 | 0.1196 | 2.65E-09 | 0.715599 | 35.41682897 |
| rs140522 | C | T | C | T | -0.0142 | 0.0144996 | 0.0022 | 0.0354 | 2.84E-10 | 0.6827 | 41.66093974 |
| rs142158911 | A | G | A | G | 0.0192 | -0.0473002 | 0.0033 | 0.0542 | 7.39E-09 | 0.3827 | 33.85106312 |
| rs156355 | C | T | C | T | 0.0214 | -0.00239712 | 0.0022 | 0.0381 | 6.41E-23 | 0.9494 | 94.61934123 |
| rs1569419 | C | T | C | T | 0.0189 | -0.0369007 | 0.0025 | 0.0391 | 6.87E-14 | 0.3454 | 57.15330192 |
| rs1613662 | A | G | A | G | -0.0167 | -0.0544995 | 0.0028 | 0.0454 | 3.90E-09 | 0.23 | 35.57251855 |
| rs1760940 | C | A | C | A | 0.0263 | 0.0130001 | 0.0024 | 0.0412 | 5.20E-27 | 0.7519 | 120.0844431 |
| rs1967556 | G | T | G | T | 0.0175 | -0.0124028 | 0.0021 | 0.0343 | 1.97E-16 | 0.7172 | 69.44408226 |
| rs2015599 | A | G | A | G | 0.0123 | -0.0452997 | 0.0021 | 0.034 | 6.03E-09 | 0.1822 | 34.30594353 |
| rs2038479 | A | C | A | C | 0.0162 | 0.00430074 | 0.0027 | 0.0435 | 1.47E-09 | 0.9218 | 35.99981224 |
| rs212938 | G | A | G | A | 0.0149 | 0.0450005 | 0.0025 | 0.0417 | 3.21E-09 | 0.2801 | 35.52141474 |
| rs2263663 | T | C | T | C | 0.02 | 0.047103 | 0.0024 | 0.0392 | 1.97E-16 | 0.2292 | 69.44408226 |
| rs2274319 | C | T | C | T | -0.0139 | -0.00320486 | 0.0022 | 0.0358 | 4.09E-10 | 0.929 | 39.91921329 |
| rs2290507 | A | G | A | G | 0.0223 | 0.074003 | 0.0031 | 0.0493 | 3.64E-13 | 0.1334 | 51.74686851 |
| rs2322718 | G | T | G | T | 0.0136 | -0.0204005 | 0.0021 | 0.034 | 1.53E-10 | 0.549301 | 41.94082434 |
| rs28851188 | A | C | A | C | -0.012 | -0.0313999 | 0.0021 | 0.0339 | 1.92E-08 | 0.3544 | 32.65289092 |
| rs2977608 | C | A | C | A | 0.0236 | -0.0221039 | 0.0025 | 0.1005 | 2.58E-21 | 0.826 | 89.11313523 |
| rs385893 | C | T | C | T | 0.0153 | 0.0175002 | 0.0021 | 0.0336 | 5.96E-13 | 0.6027 | 53.08135581 |
| rs4388979 | T | G | T | G | -0.0141 | -0.0418997 | 0.0021 | 0.0342 | 4.47E-11 | 0.2194 | 45.08139753 |
| rs445 | T | C | T | C | 0.0207 | -0.0329003 | 0.0036 | 0.0517 | 8.85E-09 | 0.524499 | 33.06232756 |
| rs453301 | G | T | G | T | 0.0121 | 0.0262002 | 0.0021 | 0.035 | 1.09E-08 | 0.454099 | 33.19937333 |
| rs4720497 | A | G | A | G | 0.0164 | -0.00380021 | 0.0025 | 0.0413 | 8.71E-11 | 0.9269 | 43.03337556 |
| rs4808075 | C | T | C | T | -0.0274 | 0.0265 | 0.0023 | 0.0373 | 3.19E-32 | 0.4783 | 141.9198647 |
| rs4846082 | T | C | T | C | -0.0179 | -0.0118004 | 0.0021 | 0.0336 | 3.33E-17 | 0.726001 | 72.65494987 |
| rs5012419 | G | A | G | A | 0.0247 | 0.0303005 | 0.0022 | 0.0348 | 2.66E-29 | 0.3832 | 126.0509955 |
| rs5745587 | G | A | G | A | 0.021 | 0.0307005 | 0.0028 | 0.0414 | 3.53E-14 | 0.4575 | 56.24970663 |
| rs5759176 | T | C | T | C | 0.0317 | -0.0605996 | 0.0023 | 0.0357 | 1.51E-44 | 0.0892997 | 189.9593117 |
| rs62641680 | A | G | A | G | -0.0903 | -0.1511 | 0.0063 | 0.1393 | 4.63E-47 | 0.2778 | 205.443373 |
| rs6959832 | A | G | A | G | -0.021 | 0.0009995 | 0.0021 | 0.0338 | 2.10E-23 | 0.9769 | 99.99947845 |
| rs7080386 | A | C | A | C | -0.0451 | 0.0156962 | 0.0021 | 0.034 | 9.58E-98 | 0.642899 | 461.2243519 |
| rs72698722 | T | C | T | C | -0.0192 | 0.0208021 | 0.0027 | 0.0438 | 1.25E-12 | 0.6353 | 50.5676375 |
| rs74874677 | G | A | G | A | -0.0819 | 0.1441 | 0.0071 | 0.1193 | 3.62E-31 | 0.2271 | 133.060405 |
| rs7705526 | A | C | A | C | 0.0178 | -0.0783995 | 0.0023 | 0.0534 | 6.46E-15 | 0.1418 | 59.89382751 |
| rs77236693 | T | C | T | C | 0.036 | -0.1219 | 0.0036 | 0.0599 | 2.08E-23 | 0.0417802 | 99.99947845 |
| rs78909033 | A | G | A | G | -0.021 | 0.0110982 | 0.0031 | 0.0584 | 1.00E-11 | 0.8498 | 45.8894589 |
| rs80140716 | C | A | C | A | -0.0252 | -0.0199987 | 0.004 | 0.0808 | 2.18E-10 | 0.8044 | 39.689793 |
| rs8121099 | C | A | C | A | 0.0221 | -0.00939572 | 0.0021 | 0.0344 | 1.63E-25 | 0.784099 | 110.7499893 |
| rs9494142 | C | T | C | T | 0.018 | -0.049704 | 0.0024 | 0.04 | 1.93E-13 | 0.2139 | 56.24970663 |
| rs9844549 | G | A | G | A | 0.0171 | 0.00269964 | 0.0025 | 0.0396 | 1.16E-11 | 0.9452 | 46.78535599 |

Abbreviations: 1, mitochondrial DNA copy number; 2, obsessive compulsive disorder; SE, Standard Error; EA, Effect allele;OA, Other allele; SNPs, single nucleotide polymorphisms.

Table S9. Genetic variants used as instrumental variables for the relationship between mitochondrial DNA copy number and Schizophrenia.

| **SNPs** | **EA 1** | **OA 1** | **EA 2** | **OA 2** | **Beta 1** | **Beta 2** | **SE 1** | **SE 2** | ***P* value 1** | ***P* value 2** | ***F*** |
| --- | --- | --- | --- | --- | --- | --- | --- | --- | --- | --- | --- |
| rs10013187 | A | C | A | C | -0.0121 | -0.0305001 | 0.0021 | 0.0087 | 9.74E-09 | 0.000419199 | 33.19937333 |
| rs10749636 | A | G | A | G | 0.0155 | 0.0107981 | 0.0025 | 0.0102 | 5.43E-10 | 0.2894 | 38.43979952 |
| rs10835226 | T | C | T | C | 0.0184 | -0.0141987 | 0.0023 | 0.0095 | 4.83E-15 | 0.1356 | 63.99966621 |
| rs11064074 | T | C | T | C | 0.0197 | -0.0109993 | 0.0021 | 0.0089 | 4.87E-20 | 0.2175 | 88.0018086 |
| rs11064881 | A | G | A | G | 0.0255 | -0.0154008 | 0.004 | 0.0171 | 2.58E-10 | 0.3671 | 40.64041304 |
| rs11082396 | C | T | C | T | 0.0258 | -0.0361005 | 0.0031 | 0.0128 | 2.54E-16 | 0.00495302 | 69.26498735 |
| rs11085147 | T | C | T | C | 0.0756 | -0.0182033 | 0.0036 | 0.0157 | 1.54E-95 | 0.2449 | 440.9977 |
| rs1127787 | A | G | A | G | -0.0159 | -0.0231011 | 0.0028 | 0.0117 | 1.49E-08 | 0.0494595 | 32.24600529 |
| rs114694170 | C | T | C | T | 0.0331 | -0.001998 | 0.0045 | 0.0196 | 3.04E-13 | 0.9198 | 54.10391535 |
| rs11553699 | G | A | G | A | 0.0445 | -0.0172012 | 0.0032 | 0.0143 | 1.45E-43 | 0.2272 | 193.3827805 |
| rs12247015 | G | A | G | A | 0.0337 | -0.0044003 | 0.0021 | 0.0088 | 1.28E-55 | 0.613999 | 257.524734 |
| rs12604328 | C | T | C | T | -0.0172 | -0.00919757 | 0.0024 | 0.0101 | 1.24E-12 | 0.362 | 51.36084324 |
| rs13084580 | T | C | T | C | 0.0244 | 0.0037972 | 0.0033 | 0.014 | 2.19E-13 | 0.7863 | 54.67005463 |
| rs1354034 | C | T | C | T | -0.0268 | -0.0247999 | 0.0022 | 0.0089 | 2.05E-35 | 0.00504301 | 148.3959203 |
| rs138055405 | T | G | T | G | 0.0488 | 0.00429923 | 0.0082 | 0.0339 | 2.65E-09 | 0.8986 | 35.41682897 |
| rs140522 | C | T | C | T | -0.0142 | -0.0028958 | 0.0022 | 0.0093 | 2.84E-10 | 0.7546 | 41.66093974 |
| rs142158911 | A | G | A | G | 0.0192 | -0.0180953 | 0.0033 | 0.0141 | 7.39E-09 | 0.1992 | 33.85106312 |
| rs156355 | C | T | C | T | 0.0214 | 0.00800193 | 0.0022 | 0.0091 | 6.41E-23 | 0.3769 | 94.61934123 |
| rs1569419 | C | T | C | T | 0.0189 | 0.00900038 | 0.0025 | 0.0105 | 6.87E-14 | 0.3891 | 57.15330192 |
| rs1613662 | A | G | A | G | -0.0167 | -0.0124028 | 0.0028 | 0.0118 | 3.90E-09 | 0.2943 | 35.57251855 |
| rs1760940 | C | A | C | A | 0.0263 | 0.00589736 | 0.0024 | 0.0107 | 5.20E-27 | 0.5822 | 120.0844431 |
| rs182346769 | A | G | A | G | 0.0213 | 0.002002 | 0.0027 | 0.0123 | 7.87E-15 | 0.8692 | 62.23424332 |
| rs1967556 | G | T | G | T | 0.0175 | 0.00810274 | 0.0021 | 0.0087 | 1.97E-16 | 0.3497 | 69.44408226 |
| rs2015599 | A | G | A | G | 0.0123 | 0.00329542 | 0.0021 | 0.0086 | 6.03E-09 | 0.7047 | 34.30594353 |
| rs2038479 | A | C | A | C | 0.0162 | 0.0168003 | 0.0027 | 0.011 | 1.47E-09 | 0.129 | 35.99981224 |
| rs212938 | G | A | G | A | 0.0149 | 0.0275048 | 0.0025 | 0.0105 | 3.21E-09 | 0.00867501 | 35.52141474 |
| rs2263663 | T | C | T | C | 0.02 | 0.0198966 | 0.0024 | 0.01 | 1.97E-16 | 0.0466799 | 69.44408226 |
| rs2274319 | C | T | C | T | -0.0139 | -0.00639948 | 0.0022 | 0.0091 | 4.09E-10 | 0.4781 | 39.91921329 |
| rs2290507 | A | G | A | G | 0.0223 | -0.0141001 | 0.0031 | 0.0128 | 3.64E-13 | 0.2729 | 51.74686851 |
| rs2322718 | G | T | G | T | 0.0136 | -0.0259991 | 0.0021 | 0.0087 | 1.53E-10 | 0.002733 | 41.94082434 |
| rs28851188 | A | C | A | C | -0.012 | 0.0149005 | 0.0021 | 0.0086 | 1.92E-08 | 0.0846292 | 32.65289092 |
| rs2977608 | C | A | C | A | 0.0236 | 0.00439966 | 0.0025 | 0.0118 | 2.58E-21 | 0.7072 | 89.11313523 |
| rs4388979 | T | G | T | G | -0.0141 | 0.00889948 | 0.0021 | 0.0088 | 4.47E-11 | 0.312 | 45.08139753 |
| rs453301 | G | T | G | T | 0.0121 | 0.0236986 | 0.0021 | 0.0087 | 1.09E-08 | 0.00633797 | 33.19937333 |
| rs4720497 | A | G | A | G | 0.0164 | -0.0193025 | 0.0025 | 0.0104 | 8.71E-11 | 0.0633695 | 43.03337556 |
| rs4807780 | C | T | C | T | -0.0191 | 0.0253999 | 0.0022 | 0.0096 | 1.61E-17 | 0.00856998 | 75.37357383 |
| rs4808075 | C | T | C | T | -0.0274 | -0.0102968 | 0.0023 | 0.0095 | 3.19E-32 | 0.2812 | 141.9198647 |
| rs4846082 | T | C | T | C | -0.0179 | 0.0151035 | 0.0021 | 0.0087 | 3.33E-17 | 0.0837896 | 72.65494987 |
| rs5012419 | G | A | G | A | 0.0247 | 0.0172988 | 0.0022 | 0.0089 | 2.66E-29 | 0.0520104 | 126.0509955 |
| rs5745587 | G | A | G | A | 0.021 | 0.0309026 | 0.0028 | 0.0106 | 3.53E-14 | 0.00358204 | 56.24970663 |
| rs5759176 | T | C | T | C | 0.0317 | -0.00330453 | 0.0023 | 0.0092 | 1.51E-44 | 0.721601 | 189.9593117 |
| rs62641680 | A | G | A | G | -0.0903 | -0.0213994 | 0.0063 | 0.0308 | 4.63E-47 | 0.4875 | 205.443373 |
| rs6959832 | A | G | A | G | -0.021 | 0.00569619 | 0.0021 | 0.0087 | 2.10E-23 | 0.5096 | 99.99947845 |
| rs7080386 | A | C | A | C | -0.0451 | -0.00019998 | 0.0021 | 0.0088 | 9.58E-98 | 0.9795 | 461.2243519 |
| rs72698722 | T | C | T | C | -0.0192 | 0.00290421 | 0.0027 | 0.011 | 1.25E-12 | 0.7921 | 50.5676375 |
| rs74874677 | G | A | G | A | -0.0819 | 0.00490199 | 0.0071 | 0.0308 | 3.62E-31 | 0.8741 | 133.060405 |
| rs7705526 | A | C | A | C | 0.0178 | -0.00950469 | 0.0023 | 0.0096 | 6.46E-15 | 0.3246 | 59.89382751 |
| rs77236693 | T | C | T | C | 0.036 | 0.0157026 | 0.0036 | 0.0148 | 2.08E-23 | 0.289 | 99.99947845 |
| rs78909033 | A | G | A | G | -0.021 | 0.00640044 | 0.0031 | 0.0136 | 1.00E-11 | 0.634501 | 45.8894589 |
| rs80140716 | C | A | C | A | -0.0252 | 0.0075988 | 0.004 | 0.0173 | 2.18E-10 | 0.6604 | 39.689793 |
| rs8121099 | C | A | C | A | 0.0221 | 0.00130085 | 0.0021 | 0.0086 | 1.63E-25 | 0.8807 | 110.7499893 |
| rs9494142 | C | T | C | T | 0.018 | -0.0238044 | 0.0024 | 0.01 | 1.93E-13 | 0.0172401 | 56.24970663 |
| rs9844549 | G | A | G | A | 0.0171 | -0.00590255 | 0.0025 | 0.0102 | 1.16E-11 | 0.5611 | 46.78535599 |

Abbreviations: 1, mitochondrial DNA copy number; 2, Schizophrenia; SE, Standard Error; EA, Effect allele;OA, Other allele; SNPs, single nucleotide polymorphisms.

Table S10. Genetic variants used as instrumental variables for the relationship between mitochondrial DNA copy number and anxiety disorders.

| **SNPs** | **EA 1** | **OA 1** | **EA 2** | **OA 2** | **Beta 1** | **Beta 2** | **SE 1** | **SE 2** | ***P* value 1** | ***P* value 2** | ***F*** |
| --- | --- | --- | --- | --- | --- | --- | --- | --- | --- | --- | --- |
| rs10013187 | A | C | A | C | -0.0121 | -0.00523214 | 0.0021 | 0.00761964 | 9.74E-09 | 0.492293 | 33.19954649 |
| rs1065853 | T | G | T | G | 0.0388 | -0.00929887 | 0.0039 | 0.0171056 | 1.58E-23 | 0.586707 | 98.97698882 |
| rs10749636 | A | G | A | G | 0.0155 | -0.00375531 | 0.0025 | 0.00924146 | 5.43E-10 | 0.684482 | 38.44 |
| rs10835226 | T | C | T | C | 0.0184 | 0.000980954 | 0.0023 | 0.00784375 | 4.83E-15 | 0.900475 | 64 |
| rs11064074 | T | C | T | C | 0.0197 | 0.0047937 | 0.0021 | 0.00772287 | 4.87E-20 | 0.534787 | 88.00226757 |
| rs11064881 | A | G | A | G | 0.0255 | -0.0220263 | 0.004 | 0.0135913 | 2.58E-10 | 0.105099 | 40.640625 |
| rs11082396 | C | T | C | T | 0.0258 | 0.0113463 | 0.0031 | 0.0114538 | 2.54E-16 | 0.321872 | 69.2653486 |
| rs11085147 | T | C | T | C | 0.0756 | 0.0093175 | 0.0036 | 0.0137712 | 1.54E-95 | 0.498664 | 441 |
| rs1127787 | A | G | A | G | -0.0159 | -0.0201448 | 0.0028 | 0.0107382 | 1.49E-08 | 0.0606569 | 32.24617347 |
| rs114694170 | C | T | C | T | 0.0331 | -0.0117358 | 0.0045 | 0.0166314 | 3.04E-13 | 0.480412 | 54.10419753 |
| rs11553699 | G | A | G | A | 0.0445 | -0.00116005 | 0.0032 | 0.0124213 | 1.45E-43 | 0.925593 | 193.3837891 |
| rs12247015 | G | A | G | A | 0.0337 | -0.00857735 | 0.0021 | 0.00812862 | 1.28E-55 | 0.291333 | 257.5260771 |
| rs12604328 | C | T | C | T | -0.0172 | -0.0047521 | 0.0024 | 0.00938487 | 1.24E-12 | 0.612606 | 51.36111111 |
| rs13084580 | T | C | T | C | 0.0244 | -0.0105583 | 0.0033 | 0.0190675 | 2.19E-13 | 0.579761 | 54.67033976 |
| rs1354034 | C | T | C | T | -0.0268 | 0.00167033 | 0.0022 | 0.00837226 | 2.05E-35 | 0.841866 | 148.3966942 |
| rs138055405 | T | G | T | G | 0.0488 | -0.023469 | 0.0082 | 0.0467132 | 2.65E-09 | 0.615382 | 35.41701368 |
| rs140522 | C | T | C | T | -0.0142 | 0.00915444 | 0.0022 | 0.00822617 | 2.84E-10 | 0.265776 | 41.66115702 |
| rs142158911 | A | G | A | G | 0.0192 | -0.00358157 | 0.0033 | 0.0125683 | 7.39E-09 | 0.775668 | 33.85123967 |
| rs156355 | C | T | C | T | 0.0214 | 0.00575656 | 0.0022 | 0.00777582 | 6.41E-23 | 0.459109 | 94.61983471 |
| rs1569419 | C | T | C | T | 0.0189 | -0.00461902 | 0.0025 | 0.00832035 | 6.87E-14 | 0.578794 | 57.1536 |
| rs1613662 | A | G | A | G | -0.0167 | 0.0213894 | 0.0028 | 0.0117312 | 3.90E-09 | 0.068259 | 35.57270408 |
| rs1760940 | C | A | C | A | 0.0263 | 0.0255218 | 0.0024 | 0.00922012 | 5.20E-27 | 0.00563923 | 120.0850694 |
| rs1967556 | G | T | G | T | 0.0175 | -0.00934404 | 0.0021 | 0.00764168 | 1.97E-16 | 0.221415 | 69.44444444 |
| rs2015599 | A | G | A | G | 0.0123 | 0.00541132 | 0.0021 | 0.00765678 | 6.03E-09 | 0.47973 | 34.30612245 |
| rs2038479 | A | C | A | C | 0.0162 | -0.00176831 | 0.0027 | 0.00924821 | 1.47E-09 | 0.848365 | 36 |
| rs212938 | G | A | G | A | 0.0149 | 0.0067941 | 0.0025 | 0.00837609 | 3.21E-09 | 0.417291 | 35.5216 |
| rs2263663 | T | C | T | C | 0.02 | -0.00738514 | 0.0024 | 0.00873755 | 1.97E-16 | 0.397989 | 69.44444444 |
| rs2274319 | C | T | C | T | -0.0139 | 0.00621429 | 0.0022 | 0.00772207 | 4.09E-10 | 0.420968 | 39.91942149 |
| rs2290507 | A | G | A | G | 0.0223 | -0.00651359 | 0.0031 | 0.010932 | 3.64E-13 | 0.551291 | 51.7471384 |
| rs2322718 | G | T | G | T | 0.0136 | -0.00642541 | 0.0021 | 0.00766492 | 1.53E-10 | 0.401869 | 41.94104308 |
| rs28851188 | A | C | A | C | -0.012 | -0.00760193 | 0.0021 | 0.00776842 | 1.92E-08 | 0.327793 | 32.65306122 |
| rs2977608 | C | A | C | A | 0.0236 | 0.00028993 | 0.0025 | 0.00826923 | 2.58E-21 | 0.972031 | 89.1136 |
| rs385893 | C | T | C | T | 0.0153 | 0.000255005 | 0.0021 | 0.00769158 | 5.96E-13 | 0.973552 | 53.08163265 |
| rs4388979 | T | G | T | G | -0.0141 | -0.0131596 | 0.0021 | 0.00790259 | 4.47E-11 | 0.0958672 | 45.08163265 |
| rs445 | T | C | T | C | 0.0207 | 0.00775792 | 0.0036 | 0.0163357 | 8.85E-09 | 0.634856 | 33.0625 |
| rs453301 | G | T | G | T | 0.0121 | 0.0125122 | 0.0021 | 0.00772819 | 1.09E-08 | 0.105439 | 33.19954649 |
| rs4720497 | A | G | A | G | 0.0164 | 0.00528283 | 0.0025 | 0.00969802 | 8.71E-11 | 0.585937 | 43.0336 |
| rs4807780 | C | T | C | T | -0.0191 | -0.0111452 | 0.0022 | 0.00811101 | 1.61E-17 | 0.169416 | 75.37396694 |
| rs4808075 | C | T | C | T | -0.0274 | -0.000143847 | 0.0023 | 0.00823144 | 3.19E-32 | 0.986057 | 141.9206049 |
| rs4846082 | T | C | T | C | -0.0179 | -0.00833301 | 0.0021 | 0.00782165 | 3.33E-17 | 0.286705 | 72.6553288 |
| rs5012419 | G | A | G | A | 0.0247 | -0.0108823 | 0.0022 | 0.00804551 | 2.66E-29 | 0.176187 | 126.0516529 |
| rs5745587 | G | A | G | A | 0.021 | 0.0209557 | 0.0028 | 0.00845973 | 3.53E-14 | 0.0132449 | 56.25 |
| rs62641680 | A | G | A | G | -0.0903 | 0.0137536 | 0.0063 | 0.0245536 | 4.63E-47 | 0.575379 | 205.4444444 |
| rs6959832 | A | G | A | G | -0.021 | 0.00107072 | 0.0021 | 0.00770726 | 2.10E-23 | 0.889511 | 100 |
| rs7080386 | A | C | A | C | -0.0451 | -0.000936972 | 0.0021 | 0.00786168 | 9.58E-98 | 0.905131 | 461.2267574 |
| rs72698722 | T | C | T | C | -0.0192 | 0.0265016 | 0.0027 | 0.00844691 | 1.25E-12 | 0.00170432 | 50.56790123 |
| rs74874677 | G | A | G | A | -0.0819 | 0.0182076 | 0.0071 | 0.0245363 | 3.62E-31 | 0.458047 | 133.061099 |
| rs7705526 | A | C | A | C | 0.0178 | 0.00649803 | 0.0023 | 0.00817051 | 6.46E-15 | 0.426438 | 59.89413989 |
| rs77236693 | T | C | T | C | 0.036 | 0.0157236 | 0.0036 | 0.0134821 | 2.08E-23 | 0.243511 | 100 |
| rs78909033 | A | G | A | G | -0.021 | -0.00163754 | 0.0031 | 0.011332 | 1.00E-11 | 0.885101 | 45.88969823 |
| rs80140716 | C | A | C | A | -0.0252 | -0.00157966 | 0.004 | 0.0274413 | 2.18E-10 | 0.954095 | 39.69 |
| rs9494142 | C | T | C | T | 0.018 | -0.0102767 | 0.0024 | 0.00820655 | 1.93E-13 | 0.210476 | 56.25 |
| rs9844549 | G | A | G | A | 0.0171 | -0.00637238 | 0.0025 | 0.00814372 | 1.16E-11 | 0.433926 | 46.7856 |

Abbreviations: 1, mitochondrial DNA copy number; 2, anxiety disorders; SE, Standard Error; EA, Effect allele;OA, Other allele; SNPs, single nucleotide polymorphisms.

Table S11. Genetic variants used as instrumental variables for the relationship between mitochondrial DNA copy number and post-traumatic stress disorder.

| **SNPs** | **EA 1** | **OA 1** | **EA 2** | **OA 2** | **Beta 1** | **Beta 2** | **SE 1** | **SE 2** | ***P* value 1** | ***P* value 2** | ***F*** |
| --- | --- | --- | --- | --- | --- | --- | --- | --- | --- | --- | --- |
| rs10013187 | A | C | A | C | -0.0121 | 0.0463346 | 0.0021 | 0.0281904 | 9.74E-09 | 0.100252 | 33.19954649 |
| rs1065853 | T | G | T | G | 0.0388 | 0.00107912 | 0.0039 | 0.0646904 | 1.58E-23 | 0.986691 | 98.97698882 |
| rs10749636 | A | G | A | G | 0.0155 | -0.0310819 | 0.0025 | 0.0342547 | 5.43E-10 | 0.364208 | 38.44 |
| rs10835226 | T | C | T | C | 0.0184 | -0.00273716 | 0.0023 | 0.02897 | 4.83E-15 | 0.924726 | 64 |
| rs11064074 | T | C | T | C | 0.0197 | 0.0147621 | 0.0021 | 0.0286048 | 4.87E-20 | 0.605804 | 88.00226757 |
| rs11064881 | A | G | A | G | 0.0255 | -4.29E-05 | 0.004 | 0.0498188 | 2.58E-10 | 0.999313 | 40.640625 |
| rs11082396 | C | T | C | T | 0.0258 | 0.0379559 | 0.0031 | 0.042581 | 2.54E-16 | 0.372725 | 69.2653486 |
| rs11085147 | T | C | T | C | 0.0756 | -0.0293243 | 0.0036 | 0.0516042 | 1.54E-95 | 0.569864 | 441 |
| rs1127787 | A | G | A | G | -0.0159 | -0.0328935 | 0.0028 | 0.0395677 | 1.49E-08 | 0.405792 | 32.24617347 |
| rs114694170 | C | T | C | T | 0.0331 | 0.0439933 | 0.0045 | 0.0615985 | 3.04E-13 | 0.475107 | 54.10419753 |
| rs11553699 | G | A | G | A | 0.0445 | 0.094278 | 0.0032 | 0.0461451 | 1.45E-43 | 0.041045 | 193.3837891 |
| rs12247015 | G | A | G | A | 0.0337 | -0.00951114 | 0.0021 | 0.0301074 | 1.28E-55 | 0.752073 | 257.5260771 |
| rs12604328 | C | T | C | T | -0.0172 | -0.00534994 | 0.0024 | 0.0346239 | 1.24E-12 | 0.877203 | 51.36111111 |
| rs13084580 | T | C | T | C | 0.0244 | 0.0947917 | 0.0033 | 0.0724504 | 2.19E-13 | 0.190749 | 54.67033976 |
| rs1354034 | C | T | C | T | -0.0268 | 0.00823661 | 0.0022 | 0.0310706 | 2.05E-35 | 0.790937 | 148.3966942 |
| rs138055405 | T | G | T | G | 0.0488 | -0.246236 | 0.0082 | 0.183308 | 2.65E-09 | 0.179176 | 35.41701368 |
| rs140522 | C | T | C | T | -0.0142 | 0.0146149 | 0.0022 | 0.0305061 | 2.84E-10 | 0.631881 | 41.66115702 |
| rs142158911 | A | G | A | G | 0.0192 | 0.0729877 | 0.0033 | 0.046188 | 7.39E-09 | 0.114054 | 33.85123967 |
| rs156355 | C | T | C | T | 0.0214 | -0.0228418 | 0.0022 | 0.0287597 | 6.41E-23 | 0.427061 | 94.61983471 |
| rs1569419 | C | T | C | T | 0.0189 | -0.0329726 | 0.0025 | 0.0307146 | 6.87E-14 | 0.283039 | 57.1536 |
| rs1613662 | A | G | A | G | -0.0167 | -0.0401097 | 0.0028 | 0.0433257 | 3.90E-09 | 0.354564 | 35.57270408 |
| rs1760940 | C | A | C | A | 0.0263 | 0.0292024 | 0.0024 | 0.0342342 | 5.20E-27 | 0.39365 | 120.0850694 |
| rs1967556 | G | T | G | T | 0.0175 | 0.0432037 | 0.0021 | 0.0281894 | 1.97E-16 | 0.125369 | 69.44444444 |
| rs2015599 | A | G | A | G | 0.0123 | 0.00238765 | 0.0021 | 0.0283796 | 6.03E-09 | 0.932951 | 34.30612245 |
| rs2038479 | A | C | A | C | 0.0162 | -0.0234924 | 0.0027 | 0.0342226 | 1.47E-09 | 0.492425 | 36 |
| rs212938 | G | A | G | A | 0.0149 | 0.0308588 | 0.0025 | 0.0309145 | 3.21E-09 | 0.318183 | 35.5216 |
| rs2263663 | T | C | T | C | 0.02 | -0.0227842 | 0.0024 | 0.0323049 | 1.97E-16 | 0.480633 | 69.44444444 |
| rs2274319 | C | T | C | T | -0.0139 | -0.00546236 | 0.0022 | 0.0285551 | 4.09E-10 | 0.848297 | 39.91942149 |
| rs2290507 | A | G | A | G | 0.0223 | -0.00570387 | 0.0031 | 0.0404507 | 3.64E-13 | 0.887863 | 51.7471384 |
| rs2322718 | G | T | G | T | 0.0136 | 0.0103461 | 0.0021 | 0.0284232 | 1.53E-10 | 0.715857 | 41.94104308 |
| rs28851188 | A | C | A | C | -0.012 | 0.0211187 | 0.0021 | 0.0288046 | 1.92E-08 | 0.463455 | 32.65306122 |
| rs2977608 | C | A | C | A | 0.0236 | 0.0313451 | 0.0025 | 0.0305225 | 2.58E-21 | 0.304444 | 89.1136 |
| rs385893 | C | T | C | T | 0.0153 | 0.0265994 | 0.0021 | 0.0283746 | 5.96E-13 | 0.348534 | 53.08163265 |
| rs4388979 | T | G | T | G | -0.0141 | -0.0226957 | 0.0021 | 0.0293067 | 4.47E-11 | 0.438683 | 45.08163265 |
| rs445 | T | C | T | C | 0.0207 | -0.0124536 | 0.0036 | 0.0601784 | 8.85E-09 | 0.836053 | 33.0625 |
| rs453301 | G | T | G | T | 0.0121 | 0.0546947 | 0.0021 | 0.0286289 | 1.09E-08 | 0.0560725 | 33.19954649 |
| rs4720497 | A | G | A | G | 0.0164 | 0.0125969 | 0.0025 | 0.0356956 | 8.71E-11 | 0.724166 | 43.0336 |
| rs4807780 | C | T | C | T | -0.0191 | 0.0264819 | 0.0022 | 0.0299989 | 1.61E-17 | 0.377364 | 75.37396694 |
| rs4808075 | C | T | C | T | -0.0274 | -0.0228418 | 0.0023 | 0.0305131 | 3.19E-32 | 0.454104 | 141.9206049 |
| rs4846082 | T | C | T | C | -0.0179 | -0.025995 | 0.0021 | 0.0289159 | 3.33E-17 | 0.368661 | 72.6553288 |
| rs5012419 | G | A | G | A | 0.0247 | 0.0148908 | 0.0022 | 0.0298024 | 2.66E-29 | 0.617321 | 126.0516529 |
| rs5745587 | G | A | G | A | 0.021 | 0.0356572 | 0.0028 | 0.0313018 | 3.53E-14 | 0.254643 | 56.25 |
| rs62641680 | A | G | A | G | -0.0903 | -0.107083 | 0.0063 | 0.0939384 | 4.63E-47 | 0.254316 | 205.4444444 |
| rs6959832 | A | G | A | G | -0.021 | 0.0200396 | 0.0021 | 0.0286085 | 2.10E-23 | 0.483631 | 100 |
| rs7080386 | A | C | A | C | -0.0451 | -0.0143222 | 0.0021 | 0.0290679 | 9.58E-98 | 0.622213 | 461.2267574 |
| rs72698722 | T | C | T | C | -0.0192 | -0.0013703 | 0.0027 | 0.031287 | 1.25E-12 | 0.965066 | 50.56790123 |
| rs74874677 | G | A | G | A | -0.0819 | 0.103585 | 0.0071 | 0.0918959 | 3.62E-31 | 0.259659 | 133.061099 |
| rs7705526 | A | C | A | C | 0.0178 | 0.0115398 | 0.0023 | 0.0302432 | 6.46E-15 | 0.702783 | 59.89413989 |
| rs77236693 | T | C | T | C | 0.036 | -0.00640613 | 0.0036 | 0.0495012 | 2.08E-23 | 0.89703 | 100 |
| rs78909033 | A | G | A | G | -0.021 | -0.0287964 | 0.0031 | 0.0419917 | 1.00E-11 | 0.49286 | 45.88969823 |
| rs80140716 | C | A | C | A | -0.0252 | 0.0524279 | 0.004 | 0.102548 | 2.18E-10 | 0.609176 | 39.69 |
| rs9494142 | C | T | C | T | 0.018 | 0.0369108 | 0.0024 | 0.030241 | 1.93E-13 | 0.222255 | 56.25 |
| rs9844549 | G | A | G | A | 0.0171 | -0.0379862 | 0.0025 | 0.029948 | 1.16E-11 | 0.204653 | 46.7856 |

Abbreviations: 1, mitochondrial DNA copy number; 2, post-traumatic stress disorder; SE, Standard Error; EA, Effect allele;OA, Other allele; SNPs, single nucleotide polymorphisms.

Table S12. Genetic variants used as instrumental variables for the relationship between Alzheimer’s disease and mitochondrial DNA copy number.

| **SNPs** | **EA 1** | **OA 1** | **EA 2** | **OA 2** | **Beta 1** | **Beta 2** | **SE 1** | **SE 2** | ***P* value 1** | ***P* value 2** | ***F*** |
| --- | --- | --- | --- | --- | --- | --- | --- | --- | --- | --- | --- |
| rs11218343 | C | T | C | T | -0.035926385 | -0.0021 | 0.005255086 | 0.0056 | 8.12E-12 | 0.7115 | 46.7376984 |
| rs11257238 | C | T | C | T | 0.012943374 | 0.0023 | 0.002261352 | 0.0022 | 1.04E-08 | 0.2856 | 32.76111574 |
| rs113260531 | A | G | A | G | 0.019985799 | -0.0051 | 0.003251406 | 0.0033 | 7.91E-10 | 0.1186 | 37.78335127 |
| rs118170342 | C | T | C | T | 0.147536684 | -0.0033 | 0.005697881 | 0.0054 | 7.93E-148 | 0.5357 | 670.4607054 |
| rs12590654 | A | G | A | G | -0.014826611 | 0.0015 | 0.002307804 | 0.0022 | 1.32E-10 | 0.5062 | 41.27489318 |
| rs1859788 | A | G | A | G | -0.018395737 | -0.0018 | 0.002312745 | 0.0023 | 1.80E-15 | 0.4174 | 63.26723919 |
| rs204473 | A | G | A | G | -0.041669554 | 0.0086 | 0.006996017 | 0.0064 | 2.58E-09 | 0.178 | 35.47610737 |
| rs2081545 | A | C | A | C | -0.017870026 | 0.0042 | 0.002229747 | 0.0022 | 1.11E-15 | 0.0509003 | 64.23020532 |
| rs28394864 | A | G | A | G | 0.01230151 | -0.0025 | 0.00218049 | 0.0021 | 1.68E-08 | 0.2372 | 31.82793834 |
| rs28399657 | G | A | G | A | -0.054635517 | 0.0193 | 0.006577122 | 0.0059 | 9.82E-17 | 0.001148 | 69.00464349 |
| rs41290120 | A | G | A | G | -0.099050195 | 0.0359 | 0.005778026 | 0.0049 | 7.14E-66 | 2.39E-13 | 293.8676135 |
| rs4236673 | A | G | A | G | -0.020161433 | -0.0026 | 0.002228715 | 0.0021 | 1.48E-19 | 0.232 | 81.83397732 |
| rs442495 | C | T | C | T | -0.013721107 | -0.0039 | 0.002257715 | 0.0023 | 1.22E-09 | 0.0855598 | 36.93515681 |
| rs4575098 | A | G | A | G | 0.016411633 | -0.0016 | 0.002576595 | 0.0025 | 1.90E-10 | 0.5142 | 40.57059016 |
| rs4663105 | C | A | C | A | 0.031095063 | 0.0013 | 0.002220228 | 0.0022 | 1.45E-44 | 0.548001 | 196.1496486 |
| rs59735493 | A | G | A | G | -0.012985226 | -0.0019 | 0.002359548 | 0.0023 | 3.73E-08 | 0.406 | 30.28595537 |
| rs6014724 | G | A | G | A | -0.022894258 | -0.005 | 0.00368812 | 0.0037 | 5.38E-10 | 0.1807 | 38.53391117 |
| rs679515 | T | C | T | C | 0.025417699 | 3.00E-04 | 0.00286314 | 0.0028 | 6.83E-19 | 0.9008 | 78.81111008 |
| rs755951 | C | A | C | A | 0.01500459 | -0.0037 | 0.002210299 | 0.0022 | 1.13E-11 | 0.0823702 | 46.08359815 |
| rs7810606 | T | C | T | C | -0.01451867 | 0.0014 | 0.0021826 | 0.0021 | 2.89E-11 | 0.5014 | 44.24919012 |
| rs846881 | C | A | C | A | -0.01736942 | -8.00E-04 | 0.002685173 | 0.0026 | 9.89E-11 | 0.7502 | 41.84330527 |
| rs867611 | G | A | G | A | -0.020428916 | -0.0016 | 0.002323835 | 0.0023 | 1.48E-18 | 0.482 | 77.28228542 |
| rs9381563 | C | T | C | T | 0.0144515 | -0.0041 | 0.002271607 | 0.0022 | 1.99E-10 | 0.0609705 | 40.47245615 |

Abbreviations: 1, Alzheimer’s disease; 2, mitochondrial DNA copy number; SE, Standard Error; EA, Effect allele;OA, Other allele; SNPs, single nucleotide polymorphisms.

Table S13. Genetic variants used as instrumental variables for the relationship between attention-deficit/hyperactivity disorder and mitochondrial DNA copy number.

| **SNPs** | **EA 1** | **OA 1** | **EA 2** | **OA 2** | **Beta 1** | **Beta 2** | **SE 1** | **SE 2** | ***P* value 1** | ***P* value 2** | ***F*** |
| --- | --- | --- | --- | --- | --- | --- | --- | --- | --- | --- | --- |
| rs10262192 | A | G | A | G | 0.073204 | -0.0055 | 0.0132 | 0.0021 | 2.89E-08 | 0.0102901 | 30.75431626 |
| rs112984125 | A | G | A | G | -0.106005 | -0.0012 | 0.0146 | 0.0023 | 3.58E-13 | 0.6111 | 52.71464705 |
| rs1222063 | A | G | A | G | 0.0962007 | -0.0013 | 0.0174 | 0.0023 | 3.07E-08 | 0.5662 | 30.5662585 |
| rs1427829 | G | A | G | A | -0.0799012 | -0.0022 | 0.0133 | 0.0021 | 1.82E-09 | 0.2892 | 36.09006262 |
| rs212178 | A | G | A | G | -0.1154 | 0.001 | 0.02 | 0.0034 | 7.68E-09 | 0.77 | 33.29169753 |
| rs281324 | C | T | C | T | 0.0744973 | 0.0118 | 0.0134 | 0.0082 | 2.68E-08 | 0.1507 | 30.90692391 |
| rs28411770 | C | T | C | T | -0.0861043 | -2.00E-04 | 0.0151 | 0.0023 | 1.15E-08 | 0.9142 | 32.51472611 |
| rs4858241 | G | T | G | T | -0.0789036 | -0.0033 | 0.014 | 0.0022 | 1.74E-08 | 0.1301 | 31.76302668 |
| rs4916723 | C | A | C | A | 0.0766003 | 8.00E-04 | 0.0135 | 0.0022 | 1.58E-08 | 0.7204 | 32.19420595 |
| rs74760947 | G | A | G | A | 0.179797 | -0.0024 | 0.0317 | 0.005 | 1.35E-08 | 0.6307 | 32.16848971 |
| rs9677504 | A | G | A | G | 0.116903 | 0.0049 | 0.0206 | 0.0037 | 1.39E-08 | 0.1826 | 32.2033599 |

Abbreviations: 1, attention-deficit/hyperactivity disorder; 2, mitochondrial DNA copy number; SE, Standard Error; EA, Effect allele;OA, Other allele; SNPs, single nucleotide polymorphisms.

Table S14. Genetic variants used as instrumental variables for the relationship between anorexia nervosa and mitochondrial DNA copy number.

| **SNPs** | **EA 1** | **OA 1** | **EA 2** | **OA 2** | **Beta 1** | **Beta 2** | **SE 1** | **SE 2** | ***P* value 1** | ***P* value 2** | ***F*** |
| --- | --- | --- | --- | --- | --- | --- | --- | --- | --- | --- | --- |
| rs11174202 | G | A | G | A | -0.152798 | -0.0013 | 0.0299 | 0.0021 | 3.11E-07 | 0.5309 | 26.11156853 |
| rs117957029 | C | T | C | T | 0.536502 | -0.0056 | 0.1024 | 0.0076 | 1.62E-07 | 0.4623 | 27.44623486 |
| rs13125782 | C | T | C | T | -0.174802 | 0.0012 | 0.0356 | 0.0026 | 9.19E-07 | 0.6441 | 24.10642432 |
| rs4622308 | T | C | T | C | -0.180096 | 2.00E-04 | 0.0307 | 0.0022 | 4.25E-09 | 0.9345 | 34.40894691 |

Abbreviations: 1, anorexia nervosa; 2, mitochondrial DNA copy number; SE, Standard Error; EA, Effect allele;OA, Other allele; SNPs, single nucleotide polymorphisms.

Table S15. Genetic variants used as instrumental variables for the relationship between autism spectrum disorder and mitochondrial DNA copy number.

| **SNPs** | **EA 1** | **OA 1** | **EA 2** | **OA 2** | **Beta 1** | **Beta 2** | **SE 1** | **SE 2** | ***P* value 1** | ***P* value 2** | ***F*** |
| --- | --- | --- | --- | --- | --- | --- | --- | --- | --- | --- | --- |
| rs11185408 | A | G | A | G | -0.0686965 | 0.0011 | 0.0138 | 0.0021 | 6.98E-07 | 0.6188 | 24.77948689 |
| rs111931861 | G | A | G | A | 0.216901 | 1.00E-04 | 0.0409 | 0.0058 | 1.12E-07 | 0.9809 | 28.12274784 |
| rs112635299 | T | G | T | G | 0.220997 | 0.0181 | 0.0432 | 0.0074 | 3.04E-07 | 0.0148399 | 26.1689636 |
| rs141455452 | G | T | G | T | -0.0784044 | -0.0048 | 0.0159 | 0.0024 | 8.94E-07 | 0.0412098 | 24.31464219 |
| rs1452075 | T | C | T | C | 0.080704 | 0.0012 | 0.0155 | 0.0024 | 2.07E-07 | 0.6261 | 27.1086559 |
| rs2224274 | T | C | T | C | 0.0709989 | -0.0013 | 0.0138 | 0.0021 | 2.86E-07 | 0.5353 | 26.46831702 |
| rs2391769 | G | A | G | A | 0.0769026 | 0.0023 | 0.0145 | 0.0022 | 1.14E-07 | 0.3008 | 28.12725186 |
| rs325485 | G | A | G | A | -0.0728043 | 8.00E-04 | 0.0143 | 0.0022 | 3.25E-07 | 0.721 | 25.91929869 |
| rs45595836 | T | C | T | C | 0.138996 | -0.0019 | 0.0272 | 0.0041 | 3.13E-07 | 0.643501 | 26.11247619 |
| rs6701243 | C | A | C | A | -0.0735014 | -1.00E-04 | 0.0144 | 0.0022 | 3.07E-07 | 0.953 | 26.05238566 |
| rs72934503 | G | A | G | A | 0.0704976 | 7.00E-04 | 0.0141 | 0.0022 | 5.89E-07 | 0.745799 | 24.99721925 |
| rs78827416 | A | G | A | G | 0.130502 | 0.0013 | 0.0266 | 0.004 | 9.00E-07 | 0.7466 | 24.06868272 |
| rs910805 | A | G | A | G | -0.0956963 | 0.001 | 0.016 | 0.0025 | 2.04E-09 | 0.6938 | 35.77104174 |
| rs9366877 | G | A | G | A | -0.0684994 | 0.0012 | 0.0139 | 0.0021 | 9.05E-07 | 0.5689 | 24.28427793 |

Abbreviations: 1, autism spectrum disorder; 2, mitochondrial DNA copy number; SE, Standard Error; EA, Effect allele;OA, Other allele; SNPs, single nucleotide polymorphisms.

Table S16. Genetic variants used as instrumental variables for the relationship between bipolar disorder and mitochondrial DNA copy number.

| **SNPs** | **EA 1** | **OA 1** | **EA 2** | **OA 2** | **Beta 1** | **Beta 2** | **SE 1** | **SE 2** | ***P* value 1** | ***P* value 2** | ***F*** |
| --- | --- | --- | --- | --- | --- | --- | --- | --- | --- | --- | --- |
| rs10043984 | T | C | T | C | 0.0593042 | 0.0044 | 0.0108 | 0.0024 | 3.71E-08 | 0.0655904 | 30.15235876 |
| rs10255167 | A | G | A | G | 0.0663962 | 3.00E-04 | 0.0118 | 0.0025 | 1.60E-08 | 0.8994 | 31.66068694 |
| rs10737496 | T | C | T | C | -0.0542041 | 0.0019 | 0.0094 | 0.0021 | 7.17E-09 | 0.3712 | 33.2511345 |
| rs10866641 | C | T | C | T | -0.0625991 | -0.0012 | 0.0094 | 0.0021 | 2.79E-11 | 0.5615 | 44.34844235 |
| rs10994415 | C | T | C | T | 0.118097 | -0.0022 | 0.0174 | 0.004 | 1.14E-11 | 0.5861 | 46.06564257 |
| rs112481526 | G | A | G | A | 0.0630996 | -0.0039 | 0.0105 | 0.0024 | 1.86E-09 | 0.1082 | 36.11374386 |
| rs113779084 | A | G | A | G | 0.0754997 | 0.0018 | 0.0102 | 0.0023 | 1.42E-13 | 0.4451 | 54.78832302 |
| rs11764361 | G | A | G | A | -0.0614995 | 0.0013 | 0.0104 | 0.0022 | 3.47E-09 | 0.5652 | 34.96828962 |
| rs12575685 | A | G | A | G | 0.0652001 | 0.0016 | 0.0101 | 0.0023 | 1.24E-10 | 0.4779 | 41.67270343 |
| rs12668848 | A | G | A | G | -0.057004 | 1.00E-04 | 0.0095 | 0.0021 | 1.90E-09 | 0.9465 | 36.00487865 |
| rs13044225 | G | A | G | A | 0.0546991 | 0.0068 | 0.0095 | 0.0021 | 8.50E-09 | 0.00130299 | 33.15210048 |
| rs1487445 | T | C | T | C | 0.0741957 | 9.00E-04 | 0.0093 | 0.0021 | 1.48E-15 | 0.6708 | 63.64869083 |
| rs17183814 | A | G | A | G | -0.102899 | 0.0028 | 0.0185 | 0.004 | 2.68E-08 | 0.4873 | 30.93689696 |
| rs174592 | G | A | G | A | 0.072001 | 0.0012 | 0.0097 | 0.0022 | 9.92E-14 | 0.5817 | 55.09744845 |
| rs2126180 | A | G | A | G | 0.0566021 | 7.00E-04 | 0.0094 | 0.0021 | 1.62E-09 | 0.740699 | 36.25828686 |
| rs2273738 | T | C | T | C | 0.0916987 | 0.0023 | 0.0136 | 0.0031 | 1.63E-11 | 0.4525 | 45.46178043 |
| rs228768 | T | G | T | G | -0.064401 | 0.0019 | 0.0102 | 0.0023 | 2.83E-10 | 0.412 | 39.86417473 |
| rs2336147 | C | T | C | T | -0.067696 | 3.00E-04 | 0.0093 | 0.0021 | 3.61E-13 | 0.8792 | 52.98561971 |
| rs237460 | T | C | T | C | 0.0553013 | 0.0033 | 0.0094 | 0.0021 | 4.25E-09 | 0.1222 | 34.61089847 |
| rs2693698 | G | A | G | A | 0.053 | -0.0036 | 0.0094 | 0.0021 | 1.96E-08 | 0.0919899 | 31.79024912 |
| rs28455634 | A | G | A | G | -0.0627964 | -0.0019 | 0.0099 | 0.0022 | 2.63E-10 | 0.3778 | 40.23435137 |
| rs28565152 | A | G | A | G | 0.0671018 | -0.0042 | 0.0112 | 0.0025 | 1.96E-09 | 0.0853297 | 35.89468896 |
| rs2953928 | A | G | A | G | 0.116096 | -0.0024 | 0.02 | 0.005 | 6.25E-09 | 0.6307 | 33.69554005 |
| rs35306827 | A | G | A | G | -0.0659001 | -0.005 | 0.0112 | 0.0025 | 3.56E-09 | 0.0474602 | 34.62055304 |
| rs35958438 | A | G | A | G | -0.0642041 | -0.0014 | 0.0117 | 0.0025 | 3.83E-08 | 0.580301 | 30.1128389 |
| rs41315395 | A | C | A | C | 0.0719042 | -0.0103 | 0.0127 | 0.003 | 1.48E-08 | 0.000622902 | 32.05523572 |
| rs4447398 | C | A | C | A | -0.0821973 | 8.00E-04 | 0.0138 | 0.0031 | 2.61E-09 | 0.7948 | 35.47764884 |
| rs4619651 | A | G | A | G | -0.0660967 | 0.0017 | 0.0101 | 0.0023 | 4.78E-11 | 0.4491 | 42.82670933 |
| rs4790841 | T | C | T | C | 0.0729041 | -0.0017 | 0.0132 | 0.0029 | 3.14E-08 | 0.5642 | 30.50379986 |
| rs5758064 | C | T | C | T | -0.0524027 | 0.0044 | 0.0093 | 0.0021 | 2.01E-08 | 0.03804 | 31.74967839 |
| rs6104027 | G | A | G | A | -0.0603046 | -0.0024 | 0.0095 | 0.0021 | 1.93E-10 | 0.2576 | 40.29503812 |
| rs61554907 | T | G | T | G | 0.0868005 | -0.0043 | 0.0154 | 0.0034 | 1.64E-08 | 0.2047 | 31.76880737 |
| rs62581014 | T | C | T | C | 0.0652001 | 0.0012 | 0.0117 | 0.0022 | 2.77E-08 | 0.5701 | 31.0543683 |
| rs6806239 | G | T | G | T | -0.0660967 | 0.0013 | 0.0119 | 0.0027 | 2.64E-08 | 0.6391 | 30.85059401 |
| rs6887473 | A | G | A | G | -0.0603046 | -1.00E-04 | 0.0105 | 0.0023 | 8.81E-09 | 0.9645 | 32.98528064 |
| rs6946056 | C | A | C | A | 0.0532004 | -0.002 | 0.0097 | 0.0022 | 3.66E-08 | 0.3574 | 30.08044287 |
| rs6954854 | A | G | A | G | -0.0582972 | 2.00E-04 | 0.0094 | 0.0021 | 5.94E-10 | 0.9176 | 38.46250666 |
| rs696366 | A | C | A | C | -0.0515957 | 9.00E-04 | 0.0094 | 0.0021 | 4.46E-08 | 0.685001 | 30.12792419 |
| rs6992333 | G | A | G | A | 0.060196 | 0.003 | 0.01 | 0.0021 | 1.62E-09 | 0.1534 | 36.23540888 |
| rs7108878 | G | T | G | T | 0.081004 | -0.0058 | 0.0147 | 0.0033 | 3.61E-08 | 0.0801992 | 30.36520096 |
| rs7201930 | C | T | C | T | 0.0586042 | -0.0031 | 0.0104 | 0.0023 | 1.89E-08 | 0.1783 | 31.75328813 |
| rs748455 | C | T | C | T | -0.0674998 | -0.0054 | 0.0103 | 0.0023 | 5.01E-11 | 0.0207401 | 42.94656387 |
| rs7707252 | G | A | G | A | 0.0571949 | 0.0018 | 0.0104 | 0.0024 | 3.64E-08 | 0.4546 | 30.24445971 |
| rs9834970 | C | T | C | T | 0.0830012 | 0.0019 | 0.0093 | 0.0021 | 6.63E-19 | 0.3579 | 79.65274456 |

Abbreviations: 1, bipolar disorder; 2, mitochondrial DNA copy number; SE, Standard Error; EA, Effect allele;OA, Other allele; SNPs, single nucleotide polymorphisms.

Table S17. Genetic variants used as instrumental variables for the relationship between major depressive disorder and mitochondrial DNA copy number.

| **SNPs** | **EA 1** | **OA 1** | **EA 2** | **OA 2** | **Beta 1** | **Beta 2** | **SE 1** | **SE 2** | ***P* value 1** | ***P* value 2** | ***F*** |
| --- | --- | --- | --- | --- | --- | --- | --- | --- | --- | --- | --- |
| rs10825942 | G | T | G | T | 0.0442027 | -0.0014 | 0.0082 | 0.0021 | 7.46E-08 | 0.5085 | 29.05794318 |
| rs117473501 | A | G | A | G | 0.586202 | 8.00E-04 | 0.1172 | 0.0085 | 5.74E-07 | 0.9242 | 25.01694926 |
| rs12129573 | A | C | A | C | 0.0477992 | 0.0023 | 0.0082 | 0.0022 | 5.45E-09 | 0.3075 | 33.97883861 |
| rs12200766 | G | A | G | A | -0.0467977 | -0.001 | 0.0091 | 0.0026 | 3.10E-07 | 0.7136 | 26.44607424 |
| rs12552 | G | A | G | A | -0.0404987 | -0.0034 | 0.008 | 0.0021 | 3.90E-07 | 0.1074 | 25.6269647 |
| rs144895331 | C | T | C | T | 0.0798011 | -0.0041 | 0.0161 | 0.0033 | 6.81E-07 | 0.2154 | 24.56750103 |
| rs1950829 | G | A | G | A | -0.0454035 | -7.00E-04 | 0.0079 | 0.0021 | 8.15E-09 | 0.7307 | 33.03082809 |
| rs2060886 | C | T | C | T | 0.0433982 | 0.0013 | 0.008 | 0.0021 | 5.76E-08 | 0.552199 | 29.4278436 |
| rs2451828 | T | C | T | C | 0.1441 | -0.0029 | 0.0273 | 0.0072 | 1.30E-07 | 0.690699 | 27.86106073 |
| rs2509805 | C | T | C | T | -0.0426954 | 0.0017 | 0.0085 | 0.0023 | 5.28E-07 | 0.446 | 25.23011914 |
| rs2756119 | A | G | A | G | -0.0425005 | 0.0041 | 0.0085 | 0.0022 | 5.96E-07 | 0.0611899 | 25.00029922 |
| rs56016904 | G | A | G | A | 0.0511986 | -0.0044 | 0.0099 | 0.0022 | 2.55E-07 | 0.04596 | 26.74488663 |
| rs58982057 | T | C | T | C | -0.051904 | -0.0012 | 0.01 | 0.0027 | 1.90E-07 | 0.661301 | 26.93994072 |
| rs6905391 | A | G | A | G | -0.0740018 | -0.007 | 0.0112 | 0.0029 | 3.47E-11 | 0.0140799 | 43.6559558 |
| rs74378177 | C | T | C | T | -0.119 | 0.0045 | 0.0239 | 0.0061 | 6.04E-07 | 0.4633 | 24.79094605 |
| rs7531118 | C | T | C | T | 0.0449974 | -0.0023 | 0.008 | 0.0021 | 2.15E-08 | 0.2742 | 31.63660312 |
| rs7856424 | T | C | T | C | -0.0449974 | 0.0054 | 0.0088 | 0.0023 | 3.01E-07 | 0.0215001 | 26.14595299 |

Abbreviations: 1, major depressive disorder; 2, mitochondrial DNA copy number; SE, Standard Error; EA, Effect allele;OA, Other allele; SNPs, single nucleotide polymorphisms.

Table S18. Genetic variants used as instrumental variables for the relationship between obsessive compulsive disorder and mitochondrial DNA copy number.

| **SNPs** | **EA 1** | **OA 1** | **EA 2** | **OA 2** | **Beta 1** | **Beta 2** | **SE 1** | **SE 2** | ***P* value 1** | ***P* value 2** | ***F*** |
| --- | --- | --- | --- | --- | --- | --- | --- | --- | --- | --- | --- |
| rs12568997 | A | G | A | G | -0.2933 | 0.0019 | 0.058 | 0.0023 | 4.23E-07 | 0.416 | 25.57069516 |
| rs4733767 | A | G | A | G | 0.193501 | -0.0057 | 0.039 | 0.0024 | 7.10E-07 | 0.0175299 | 24.61566708 |

Abbreviations: 1, obsessive compulsive disorder; 2, mitochondrial DNA copy number; SE, Standard Error; EA, Effect allele;OA, Other allele; SNPs, single nucleotide polymorphisms.

Table S19. Genetic variants used as instrumental variables for the relationship between Schizophrenia and mitochondrial DNA copy number.

| **SNPs** | **EA 1** | **OA 1** | **EA 2** | **OA 2** | **Beta 1** | **Beta 2** | **SE 1** | **SE 2** | ***P* value 1** | ***P* value 2** | ***F*** |
| --- | --- | --- | --- | --- | --- | --- | --- | --- | --- | --- | --- |
| rs10035564 | G | A | G | A | 0.0668024 | -9.00E-04 | 0.0092 | 0.0023 | 4.38E-13 | 0.6832 | 52.72320805 |
| rs10086619 | G | A | G | A | 0.0722052 | 0.0018 | 0.0116 | 0.0028 | 4.97E-10 | 0.519501 | 38.7448803 |
| rs10117 | A | G | A | G | -0.0549994 | -0.0034 | 0.0088 | 0.0022 | 4.66E-10 | 0.1204 | 39.06104975 |
| rs10861176 | A | G | A | G | 0.0555021 | 2.00E-04 | 0.0098 | 0.0024 | 1.59E-08 | 0.934 | 32.07451006 |
| rs10873538 | G | T | G | T | 0.0665031 | -0.001 | 0.0091 | 0.0022 | 3.01E-13 | 0.6427 | 53.40652825 |
| rs10957321 | A | G | A | G | 0.0475949 | 0.0037 | 0.0086 | 0.0021 | 3.48E-08 | 0.0810905 | 30.62790464 |
| rs11027839 | C | A | C | A | 0.0515038 | -1.00E-04 | 0.0086 | 0.0021 | 2.40E-09 | 0.9644 | 35.86534351 |
| rs11136325 | A | G | A | G | -0.0537967 | -0.0022 | 0.0091 | 0.0022 | 3.05E-09 | 0.312 | 34.94796071 |
| rs11165867 | T | C | T | C | 0.0743034 | -0.0046 | 0.0116 | 0.0029 | 1.30E-10 | 0.1073 | 41.02936037 |
| rs11191580 | C | T | C | T | -0.131703 | -0.0023 | 0.0155 | 0.0039 | 1.77E-17 | 0.552199 | 72.19735554 |
| rs11210892 | A | G | A | G | -0.0635005 | -0.0031 | 0.0091 | 0.0023 | 2.68E-12 | 0.1729 | 48.69281211 |
| rs113264400 | C | T | C | T | 0.112296 | 0.0048 | 0.0202 | 0.005 | 2.86E-08 | 0.3356 | 30.90431959 |
| rs11664298 | A | G | A | G | 0.0773995 | 0.0022 | 0.0108 | 0.0026 | 8.94E-13 | 0.3987 | 51.35966127 |
| rs11693094 | T | C | T | C | -0.054403 | -3.00E-04 | 0.0087 | 0.0021 | 4.29E-10 | 0.8905 | 39.10214163 |
| rs11696755 | C | T | C | T | 0.0636962 | 0.0059 | 0.011 | 0.0027 | 7.26E-09 | 0.0301002 | 33.53011391 |
| rs11941714 | A | G | A | G | -0.0515957 | 0.0031 | 0.0093 | 0.0023 | 3.07E-08 | 0.1764 | 30.77899763 |
| rs12129573 | A | C | A | C | 0.0777994 | 0.0023 | 0.0089 | 0.0022 | 2.28E-18 | 0.3075 | 76.41275067 |
| rs12151767 | A | G | A | G | -0.0611045 | -7.00E-04 | 0.0086 | 0.0021 | 1.31E-12 | 0.734301 | 50.48273068 |
| rs12285419 | A | C | A | C | 0.0849045 | -0.0011 | 0.011 | 0.0027 | 1.05E-14 | 0.6849 | 59.57573358 |
| rs12293670 | G | A | G | A | -0.0704957 | 0.0011 | 0.0092 | 0.0023 | 1.56E-14 | 0.6407 | 58.71417343 |
| rs12489270 | C | T | C | T | 0.0579046 | 0.001 | 0.0089 | 0.0022 | 7.47E-11 | 0.636999 | 42.32914243 |
| rs12652777 | C | T | C | T | -0.0487997 | -0.0023 | 0.0086 | 0.0021 | 1.52E-08 | 0.2809 | 32.19813769 |
| rs12712510 | C | T | C | T | -0.0574006 | 0.0038 | 0.0087 | 0.0021 | 5.14E-11 | 0.0753703 | 43.52990409 |
| rs12771371 | A | G | A | G | -0.0524027 | -2.00E-04 | 0.0093 | 0.0023 | 1.94E-08 | 0.9201 | 31.74934592 |
| rs12833624 | T | C | T | C | 0.0501992 | 0.0023 | 0.009 | 0.0022 | 2.77E-08 | 0.3009 | 31.11013708 |
| rs12883788 | T | C | T | C | 0.0613011 | -0.0016 | 0.0087 | 0.0021 | 1.86E-12 | 0.4507 | 49.64681376 |
| rs13016542 | C | T | C | T | -0.0883039 | -0.0017 | 0.0129 | 0.0032 | 8.28E-12 | 0.584501 | 46.85691595 |
| rs13107325 | T | C | T | C | 0.158703 | 0.0082 | 0.0168 | 0.004 | 2.90E-21 | 0.0406097 | 89.23700862 |
| rs13195636 | C | A | C | A | -0.210504 | -0.0082 | 0.0159 | 0.0033 | 6.55E-40 | 0.0128801 | 175.2749324 |
| rs13233308 | T | C | T | C | -0.0487044 | 0.0021 | 0.0086 | 0.0021 | 1.75E-08 | 0.3104 | 32.07250223 |
| rs132582 | T | C | T | C | -0.0509972 | 0.0011 | 0.0086 | 0.0021 | 3.26E-09 | 0.6019 | 35.16325844 |
| rs1430894 | T | C | T | C | 0.0532953 | 0.0014 | 0.0086 | 0.0021 | 6.15E-10 | 0.513701 | 38.40380637 |
| rs145071536 | C | T | C | T | 0.0851005 | 0.0026 | 0.012 | 0.0027 | 1.62E-12 | 0.3462 | 50.29155717 |
| rs1451488 | G | A | G | A | 0.0708947 | 0.0041 | 0.0087 | 0.0021 | 4.47E-16 | 0.0548403 | 66.40218715 |
| rs149165 | G | T | G | T | -0.0481995 | -0.0065 | 0.0087 | 0.0021 | 3.00E-08 | 0.00214299 | 30.69304049 |
| rs1593304 | G | A | G | A | 0.0641013 | 0.0046 | 0.0111 | 0.0026 | 7.45E-09 | 0.0777392 | 33.34886582 |
| rs1611236 | A | G | A | G | -0.0551035 | 0.0036 | 0.0096 | 0.0023 | 8.47E-09 | 0.1167 | 32.94649771 |
| rs1615350 | T | C | T | C | -0.0736036 | -3.00E-04 | 0.0098 | 0.0024 | 4.92E-14 | 0.8999 | 56.40781964 |
| rs167924 | G | A | G | A | 0.0501992 | -0.0038 | 0.009 | 0.0022 | 2.34E-08 | 0.0797297 | 31.11013708 |
| rs16851048 | C | T | C | T | 0.0744973 | 3.00E-04 | 0.0107 | 0.0027 | 4.15E-12 | 0.9107 | 48.47377715 |
| rs16867571 | G | A | G | A | -0.0657035 | -5.00E-04 | 0.0104 | 0.0025 | 2.68E-10 | 0.8386 | 39.91201761 |
| rs17194490 | T | G | T | G | 0.0781994 | 0.0011 | 0.0116 | 0.0028 | 1.80E-11 | 0.6917 | 45.44480191 |
| rs17731 | A | G | A | G | 0.0523992 | -0.002 | 0.0089 | 0.0022 | 4.37E-09 | 0.3487 | 34.66272097 |
| rs1860002 | T | C | T | C | -0.0837987 | 0.0012 | 0.0087 | 0.0021 | 1.04E-21 | 0.5616 | 92.77466799 |
| rs187557 | T | C | T | C | -0.0666956 | -0.0014 | 0.0119 | 0.003 | 2.03E-08 | 0.6297 | 31.41187036 |
| rs1881046 | T | G | T | G | -0.0507026 | -1.00E-04 | 0.0092 | 0.0022 | 3.39E-08 | 0.9518 | 30.37233331 |
| rs1901512 | C | T | C | T | -0.058401 | 0.0015 | 0.0094 | 0.0023 | 5.72E-10 | 0.52 | 38.5991918 |
| rs1915019 | G | A | G | A | -0.0570984 | -0.0029 | 0.0098 | 0.0025 | 6.57E-09 | 0.2391 | 33.94603678 |
| rs2053079 | G | A | G | A | 0.0598986 | 0.0041 | 0.0101 | 0.0025 | 3.01E-09 | 0.095971 | 35.17093772 |
| rs2078266 | G | A | G | A | -0.0696007 | -0.0049 | 0.0126 | 0.0031 | 2.94E-08 | 0.1101 | 30.5126183 |
| rs215412 | A | G | A | G | 0.0577033 | 9.00E-04 | 0.0091 | 0.0023 | 2.69E-10 | 0.7003 | 40.20794418 |
| rs2167378 | T | C | T | C | -0.0648978 | -0.0049 | 0.0087 | 0.0021 | 7.30E-14 | 0.0213398 | 55.64354563 |
| rs217336 | A | C | A | C | -0.0503033 | 0.0036 | 0.0087 | 0.0022 | 8.05E-09 | 0.0969996 | 33.43087929 |
| rs2252074 | G | T | G | T | 0.0685037 | -0.0022 | 0.0088 | 0.0022 | 6.19E-15 | 0.3125 | 60.59768948 |
| rs2333321 | G | A | G | A | -0.0712038 | -0.0024 | 0.0105 | 0.0026 | 1.25E-11 | 0.3509 | 45.98551945 |
| rs2381411 | C | T | C | T | 0.050399 | 0.0012 | 0.0088 | 0.0022 | 1.25E-08 | 0.5784 | 32.79984912 |
| rs2455415 | T | C | T | C | 0.0494949 | 0.0025 | 0.0088 | 0.0021 | 1.69E-08 | 0.241 | 31.63362117 |
| rs2456020 | T | C | T | C | -0.0815984 | 7.00E-04 | 0.0102 | 0.0025 | 1.13E-15 | 0.774599 | 63.9965105 |
| rs2514218 | T | C | T | C | -0.0704957 | -0.003 | 0.0092 | 0.0022 | 1.35E-14 | 0.1809 | 58.71417343 |
| rs2696466 | G | A | G | A | -0.0611986 | 0.0056 | 0.0092 | 0.0022 | 2.64E-11 | 0.00949905 | 44.24871581 |
| rs2710323 | C | T | C | T | -0.0784044 | -4.00E-04 | 0.0086 | 0.0021 | 1.23E-19 | 0.8465 | 83.11460022 |
| rs2815731 | A | C | A | C | -0.0600033 | 0.001 | 0.0091 | 0.0022 | 4.39E-11 | 0.6507 | 43.47712708 |
| rs2909457 | A | G | A | G | -0.0489997 | 4.00E-04 | 0.0087 | 0.0021 | 1.48E-08 | 0.852 | 31.72062154 |
| rs2999392 | T | C | T | C | 0.0517987 | 0.0029 | 0.0094 | 0.0023 | 3.05E-08 | 0.2059 | 30.36514539 |
| rs308697 | A | C | A | C | -0.0501036 | -6.00E-04 | 0.0087 | 0.0021 | 8.83E-09 | 0.791899 | 33.16597044 |
| rs34555420 | T | G | T | G | -0.168696 | -0.013 | 0.0173 | 0.0035 | 1.54E-22 | 0.000232199 | 95.08471634 |
| rs35351411 | C | A | C | A | 0.0635044 | -0.0032 | 0.0087 | 0.0021 | 2.21E-13 | 0.1366 | 53.27978705 |
| rs35426637 | T | G | T | G | -0.0622985 | 0.0028 | 0.0093 | 0.0023 | 2.15E-11 | 0.2147 | 44.87274468 |
| rs35734242 | C | T | C | T | 0.050704 | -0.0042 | 0.0089 | 0.0021 | 1.37E-08 | 0.0472096 | 32.45620829 |
| rs3739118 | A | G | A | G | -0.057004 | -0.0019 | 0.0095 | 0.0023 | 2.36E-09 | 0.4268 | 36.00450162 |
| rs3791710 | C | T | C | T | -0.0600033 | 0.0014 | 0.0108 | 0.0026 | 3.02E-08 | 0.604301 | 30.86712014 |
| rs3795310 | T | C | T | C | -0.0509972 | 8.00E-04 | 0.0087 | 0.0021 | 5.75E-09 | 0.723401 | 34.35955336 |
| rs3802924 | C | A | C | A | -0.0736036 | 0.0048 | 0.0108 | 0.0027 | 9.58E-12 | 0.0714496 | 46.44553325 |
| rs3814883 | T | C | T | C | -0.0670977 | -0.0039 | 0.0087 | 0.0021 | 1.58E-14 | 0.0629999 | 59.47988405 |
| rs3824451 | C | T | C | T | 0.0655951 | 0.0021 | 0.0118 | 0.0029 | 2.54E-08 | 0.4553 | 30.90097152 |
| rs4129585 | C | A | C | A | -0.0749962 | 2.00E-04 | 0.0087 | 0.0021 | 5.11E-18 | 0.9085 | 74.30762203 |
| rs4575535 | G | A | G | A | 0.0557982 | -0.0015 | 0.0096 | 0.0023 | 5.77E-09 | 0.508401 | 33.78245942 |
| rs4632195 | T | C | T | C | 0.0471964 | 0.0017 | 0.0086 | 0.0021 | 4.59E-08 | 0.4168 | 30.11717243 |
| rs4636654 | A | G | A | G | -0.0483043 | 0.0013 | 0.0089 | 0.0022 | 4.89E-08 | 0.5579 | 29.45675645 |
| rs4653164 | T | C | T | C | 0.0511038 | -0.0024 | 0.0092 | 0.0023 | 3.08E-08 | 0.3009 | 30.85489596 |
| rs4702 | A | G | A | G | -0.0843044 | -1.00E-04 | 0.0089 | 0.0021 | 2.79E-21 | 0.9677 | 89.7250733 |
| rs4766428 | T | C | T | C | 0.0750038 | 0.001 | 0.0089 | 0.0021 | 3.93E-17 | 0.631401 | 71.01986989 |
| rs4779050 | G | T | G | T | -0.0579953 | 9.00E-04 | 0.0089 | 0.0022 | 7.27E-11 | 0.6951 | 42.46185244 |
| rs4812325 | A | G | A | G | 0.0719042 | 0.002 | 0.0089 | 0.0022 | 8.96E-16 | 0.3658 | 65.27123883 |
| rs4921741 | G | A | G | A | 0.0559991 | -0.0052 | 0.0098 | 0.0025 | 1.21E-08 | 0.0388097 | 32.65151181 |
| rs500102 | C | T | C | T | -0.0517002 | 9.00E-04 | 0.0088 | 0.0022 | 4.87E-09 | 0.6854 | 34.51536365 |
| rs505061 | A | C | A | C | 0.0534957 | -0.0016 | 0.0086 | 0.0021 | 5.80E-10 | 0.4398 | 38.69315992 |
| rs56205728 | A | G | A | G | 0.0630037 | -7.00E-04 | 0.0097 | 0.0024 | 1.01E-10 | 0.784401 | 42.18732539 |
| rs56335113 | G | A | G | A | -0.064701 | 0.0032 | 0.0094 | 0.0023 | 6.02E-12 | 0.1677 | 47.3761353 |
| rs5751191 | C | T | C | T | 0.0655951 | 0.0019 | 0.0086 | 0.0021 | 3.00E-14 | 0.3647 | 58.1753823 |
| rs58120505 | C | T | C | T | -0.089603 | 1.00E-04 | 0.0088 | 0.0021 | 2.24E-24 | 0.9729 | 103.6747766 |
| rs6001259 | T | C | T | C | 0.1915 | -0.0058 | 0.0348 | 0.0092 | 3.70E-08 | 0.532299 | 30.28113884 |
| rs6010045 | C | T | C | T | 0.0548998 | -0.0029 | 0.0095 | 0.0023 | 7.44E-09 | 0.197 | 33.39547662 |
| rs60135207 | T | G | T | G | -0.0495994 | 0.0038 | 0.0088 | 0.0021 | 1.53E-08 | 0.0759591 | 31.76734012 |
| rs61937595 | T | C | T | C | -0.130098 | -0.0034 | 0.0162 | 0.0037 | 1.15E-15 | 0.3596 | 64.49180954 |
| rs62018952 | C | T | C | T | 0.0584027 | 0.0012 | 0.0097 | 0.0024 | 1.94E-09 | 0.6236 | 36.25064461 |
| rs62183855 | C | A | C | A | -0.0660967 | -0.0024 | 0.0111 | 0.0026 | 2.66E-09 | 0.3562 | 35.457405 |
| rs634940 | T | G | T | G | 0.0663962 | -0.0025 | 0.0099 | 0.0024 | 1.78E-11 | 0.2901 | 44.97896017 |
| rs6482437 | C | A | C | A | 0.0989036 | -0.0015 | 0.0142 | 0.0034 | 3.33E-12 | 0.653 | 48.51107094 |
| rs6538539 | T | G | T | G | -0.0567961 | -0.0012 | 0.0086 | 0.0021 | 4.43E-11 | 0.574 | 43.61475922 |
| rs6546857 | G | A | G | A | 0.0603978 | 0.0029 | 0.0102 | 0.0025 | 2.74E-09 | 0.2418 | 35.06188389 |
| rs6549963 | C | T | C | T | -0.0483043 | -0.0029 | 0.0088 | 0.0021 | 4.31E-08 | 0.18 | 30.130032 |
| rs6673880 | G | A | G | A | 0.062301 | -3.00E-04 | 0.0091 | 0.0021 | 7.19E-12 | 0.8742 | 46.8706096 |
| rs6715366 | A | G | A | G | 0.0540972 | -0.0019 | 0.0097 | 0.0024 | 2.49E-08 | 0.4289 | 31.10279782 |
| rs6798742 | G | A | G | A | 0.0610991 | -0.0027 | 0.0093 | 0.0023 | 4.57E-11 | 0.2425 | 43.16155476 |
| rs6943762 | C | T | C | T | -0.105098 | -0.0016 | 0.0132 | 0.0032 | 1.57E-15 | 0.610401 | 63.39199099 |
| rs6974218 | C | A | C | A | -0.0548953 | 0.0021 | 0.0089 | 0.0022 | 6.80E-10 | 0.3395 | 38.04378019 |
| rs6984242 | A | G | A | G | -0.0546965 | -0.0032 | 0.0087 | 0.0022 | 3.85E-10 | 0.1412 | 39.5251858 |
| rs708228 | T | C | T | C | 0.0527997 | -0.0013 | 0.0091 | 0.0022 | 6.56E-09 | 0.5581 | 33.6646014 |
| rs7112616 | C | T | C | T | -0.0522034 | -0.0032 | 0.0086 | 0.0021 | 1.52E-09 | 0.1357 | 36.84631222 |
| rs7113199 | C | A | C | A | -0.0522983 | -1.00E-04 | 0.0094 | 0.0023 | 2.80E-08 | 0.9514 | 30.95371561 |
| rs72802868 | T | G | T | G | -0.0691995 | 7.00E-04 | 0.0096 | 0.0023 | 4.55E-13 | 0.7716 | 51.95852315 |
| rs72986630 | T | C | T | C | 0.112296 | 1.00E-04 | 0.0179 | 0.0045 | 3.59E-10 | 0.9832 | 39.35644507 |
| rs73229090 | A | C | A | C | -0.102602 | 0.0092 | 0.0142 | 0.0033 | 4.34E-13 | 0.00529395 | 52.20694925 |
| rs7515363 | T | C | T | C | -0.0535029 | 0.0027 | 0.0089 | 0.0022 | 1.84E-09 | 0.2227 | 36.13832201 |
| rs7575796 | G | A | G | A | -0.0963006 | 1.00E-04 | 0.0172 | 0.004 | 2.07E-08 | 0.9848 | 31.34688882 |
| rs7634476 | G | A | G | A | 0.0577033 | -0.0038 | 0.0088 | 0.0022 | 5.46E-11 | 0.0785398 | 42.9961242 |
| rs7647398 | T | C | T | C | -0.0774979 | 0.0016 | 0.0109 | 0.0026 | 1.07E-12 | 0.5405 | 50.5498911 |
| rs778371 | G | A | G | A | 0.0806029 | -0.0044 | 0.0095 | 0.0024 | 1.49E-17 | 0.0599805 | 71.98590615 |
| rs7798283 | G | T | G | T | -0.074003 | -0.0022 | 0.0134 | 0.0032 | 3.49E-08 | 0.487 | 30.49877574 |
| rs79210963 | C | T | C | T | 0.0856015 | 3.00E-04 | 0.0137 | 0.0033 | 4.14E-10 | 0.938 | 39.04046367 |
| rs79445414 | C | T | C | T | 0.1234 | -0.0038 | 0.0222 | 0.0051 | 2.80E-08 | 0.454099 | 30.89710024 |
| rs8138941 | A | G | A | G | 0.0580953 | -7.00E-04 | 0.0106 | 0.0026 | 4.46E-08 | 0.792899 | 30.03748855 |
| rs9304548 | A | C | A | C | -0.0567016 | 3.00E-04 | 0.01 | 0.0024 | 1.59E-08 | 0.9043 | 32.15022224 |
| rs9318627 | C | A | C | A | -0.0611986 | -8.00E-04 | 0.0088 | 0.0021 | 4.35E-12 | 0.7135 | 48.36274931 |
| rs9454727 | G | A | G | A | -0.054403 | 0.0029 | 0.0098 | 0.0024 | 3.35E-08 | 0.2232 | 30.81675448 |
| rs9461916 | C | T | C | T | 0.0532953 | 0.001 | 0.0088 | 0.0022 | 1.64E-09 | 0.6367 | 36.67801549 |
| rs9636107 | G | A | G | A | 0.0698969 | -3.00E-04 | 0.0086 | 0.0021 | 5.11E-16 | 0.8753 | 66.05600105 |
| rs9687282 | G | T | G | T | 0.0525994 | -0.0019 | 0.0091 | 0.0022 | 7.33E-09 | 0.4096 | 33.40966702 |
| rs9876421 | T | C | T | C | 0.0625033 | 0.0024 | 0.0092 | 0.0022 | 9.19E-12 | 0.2909 | 46.155514 |

Abbreviations: 1, Schizophrenia; 2, mitochondrial DNA copy number; SE, Standard Error; EA, Effect allele;OA, Other allele; SNPs, single nucleotide polymorphisms.

Table S20. Genetic variants used as instrumental variables for the relationship between anxiety disorders and mitochondrial DNA copy number.

| **SNPs** | **EA 1** | **OA 1** | **EA 2** | **OA 2** | **Beta 1** | **Beta 2** | **SE 1** | **SE 2** | ***P* value 1** | ***P* value 2** | ***F*** |
| --- | --- | --- | --- | --- | --- | --- | --- | --- | --- | --- | --- |
| rs10065355 | C | T | C | T | -0.0393526 | 0.0011 | 0.0076965 | 0.0022 | 3.17E-07 | 0.609901 | 26.14329449 |
| rs10092618 | A | G | A | G | 0.0580949 | 0.0034 | 0.00849686 | 0.0022 | 8.07E-12 | 0.1175 | 46.74757551 |
| rs10093007 | A | G | A | G | -0.0430119 | -0.0037 | 0.00851311 | 0.0025 | 4.36E-07 | 0.1268 | 25.52705798 |
| rs10752913 | G | A | G | A | 0.0469873 | 0.0012 | 0.00925525 | 0.0027 | 3.84E-07 | 0.670199 | 25.774169 |
| rs12310367 | G | A | G | A | 0.0424523 | 0.002 | 0.00831509 | 0.0022 | 3.30E-07 | 0.3713 | 26.06564888 |
| rs12548983 | C | T | C | T | -0.0398593 | -0.0036 | 0.0077282 | 0.0022 | 2.50E-07 | 0.0945802 | 26.60128581 |
| rs13185476 | G | A | G | A | 0.0488288 | -1.00E-04 | 0.00942843 | 0.0031 | 2.23E-07 | 0.9675 | 26.82089961 |
| rs13244325 | G | T | G | T | -0.055893 | -0.0011 | 0.0109043 | 0.0033 | 2.96E-07 | 0.7426 | 26.27358032 |
| rs1336189 | A | C | A | C | -0.0375299 | -0.0024 | 0.00765983 | 0.0022 | 9.60E-07 | 0.2815 | 24.00582038 |
| rs145274568 | A | G | A | G | 0.0710007 | -0.0097 | 0.012768 | 0.0072 | 2.69E-08 | 0.1819 | 30.92284994 |
| rs145281382 | T | C | T | C | 0.106083 | 0.0309 | 0.0169065 | 0.0768 | 3.50E-10 | 0.6876 | 39.37170009 |
| rs1480567 | C | T | C | T | 0.047968 | 0.002 | 0.00860221 | 0.0027 | 2.46E-08 | 0.4458 | 31.09446876 |
| rs153806 | A | G | A | G | 0.0377499 | 0.0016 | 0.00768158 | 0.0021 | 8.91E-07 | 0.4423 | 24.15074294 |
| rs16923640 | A | G | A | G | 0.0407118 | 6.00E-04 | 0.00828443 | 0.0024 | 8.91E-07 | 0.793601 | 24.14990037 |
| rs17201899 | A | G | A | G | 0.154863 | -0.001 | 0.0314909 | 0.0064 | 8.76E-07 | 0.8777 | 24.18383609 |
| rs17405662 | T | C | T | C | -0.0522498 | 8.00E-04 | 0.00960507 | 0.0026 | 5.33E-08 | 0.757399 | 29.59158286 |
| rs197261 | G | A | G | A | -0.0475201 | -9.00E-04 | 0.00910243 | 0.0024 | 1.78E-07 | 0.7022 | 27.25461153 |
| rs2071044 | T | C | T | C | -0.0444189 | -2.00E-04 | 0.00766037 | 0.0021 | 6.69E-09 | 0.9167 | 33.62297248 |
| rs2127346 | G | A | G | A | -0.0385229 | -0.0016 | 0.00763508 | 0.0021 | 4.52E-07 | 0.4514 | 25.45720752 |
| rs2397672 | A | G | A | G | 0.0474339 | -6.00E-04 | 0.00826477 | 0.0023 | 9.51E-09 | 0.782499 | 32.9394333 |
| rs2430979 | T | C | T | C | 0.0512182 | 0.0076 | 0.0104368 | 0.0029 | 9.23E-07 | 0.00982698 | 24.08318384 |
| rs283038 | A | C | A | C | -0.0378599 | 0.0019 | 0.00769105 | 0.0021 | 8.54E-07 | 0.3695 | 24.23191046 |
| rs2904662 | A | G | A | G | 0.0408906 | -0.0039 | 0.00777262 | 0.0021 | 1.43E-07 | 0.0620297 | 27.67655907 |
| rs4619804 | C | A | C | A | 0.0507766 | -0.0022 | 0.00808434 | 0.0024 | 3.37E-10 | 0.3571 | 39.44919032 |
| rs4702 | A | G | A | G | -0.0386799 | -1.00E-04 | 0.00767126 | 0.0021 | 4.60E-07 | 0.9677 | 25.42361366 |
| rs4870060 | A | G | A | G | -0.0412159 | -0.0032 | 0.00788481 | 0.0022 | 1.72E-07 | 0.1485 | 27.32417827 |
| rs4955420 | C | T | C | T | -0.0464927 | -0.0085 | 0.00850427 | 0.0022 | 4.58E-08 | 0.000137101 | 29.88790374 |
| rs6123193 | T | C | T | C | -0.0477711 | 0.0076 | 0.00953863 | 0.0025 | 5.50E-07 | 0.002334 | 25.08178711 |
| rs62081501 | A | G | A | G | 0.0720813 | 0.0069 | 0.0134173 | 0.0039 | 7.78E-08 | 0.07358 | 28.86123829 |
| rs62099231 | A | G | A | G | 0.0432544 | 0.002 | 0.00764966 | 0.0021 | 1.56E-08 | 0.349 | 31.97247958 |
| rs6751342 | C | A | C | A | -0.0405649 | -0.0015 | 0.00800109 | 0.0024 | 3.98E-07 | 0.514301 | 25.70410628 |
| rs67774492 | G | A | G | A | -0.0562117 | -7.00E-04 | 0.0111027 | 0.0031 | 4.13E-07 | 0.8261 | 25.63281064 |
| rs7305960 | G | A | G | A | 0.0457877 | 0.0036 | 0.00899894 | 0.0026 | 3.62E-07 | 0.1604 | 25.8889803 |
| rs75429814 | C | T | C | T | -0.0480344 | -0.0056 | 0.00911308 | 0.0027 | 1.36E-07 | 0.0403404 | 27.78269524 |
| rs76418138 | A | G | A | G | -0.0816928 | -0.004 | 0.016637 | 0.0041 | 9.09E-07 | 0.3287 | 24.11112808 |
| rs77333198 | A | G | A | G | -0.0605951 | 0.0042 | 0.012017 | 0.0029 | 4.60E-07 | 0.1442 | 25.42628383 |
| rs77548727 | T | C | T | C | 0.110643 | 0.0171 | 0.0213724 | 0.0055 | 2.26E-07 | 0.002053 | 26.80040137 |
| rs77578399 | C | T | C | T | -0.0773514 | -0.0165 | 0.014436 | 0.0149 | 8.40E-08 | 0.2679 | 28.71062259 |
| rs77683334 | T | C | T | C | -0.121261 | 0.0048 | 0.0207452 | 0.0068 | 5.06E-09 | 0.4864 | 34.16701502 |
| rs79004665 | T | C | T | C | -0.053628 | 0.0011 | 0.0104987 | 0.0035 | 3.25E-07 | 0.758099 | 26.09228716 |
| rs9468225 | G | T | G | T | -0.0506514 | -0.0051 | 0.00963437 | 0.0024 | 1.46E-07 | 0.0361701 | 27.63988714 |

Abbreviations: 1, anxiety disorders; 2, mitochondrial DNA copy number; SE, Standard Error; EA, Effect allele;OA, Other allele; SNPs, single nucleotide polymorphisms.

Table S21. Genetic variants used as instrumental variables for the relationship between post-traumatic stress disorder and mitochondrial DNA copy number.

| **SNPs** | **EA 1** | **OA 1** | **EA 2** | **OA 2** | **Beta 1** | **Beta 2** | **SE 1** | **SE 2** | ***P* value 1** | ***P* value 2** | ***F*** |
| --- | --- | --- | --- | --- | --- | --- | --- | --- | --- | --- | --- |
| rs11864961 | A | G | A | G | -0.199385 | -3.00E-04 | 0.0379102 | 0.0031 | 1.45E-07 | 0.912 | 27.66131448 |
| rs6021953 | A | G | A | G | -0.169886 | 0.006 | 0.0324023 | 0.0023 | 1.58E-07 | 0.00834699 | 27.48928883 |

Abbreviations: 1, post-traumatic stress disorder; 2, mitochondrial DNA copy number; SE, Standard Error; EA, Effect allele;OA, Other allele; SNPs, single nucleotide polymorphisms.

**B**

**A**

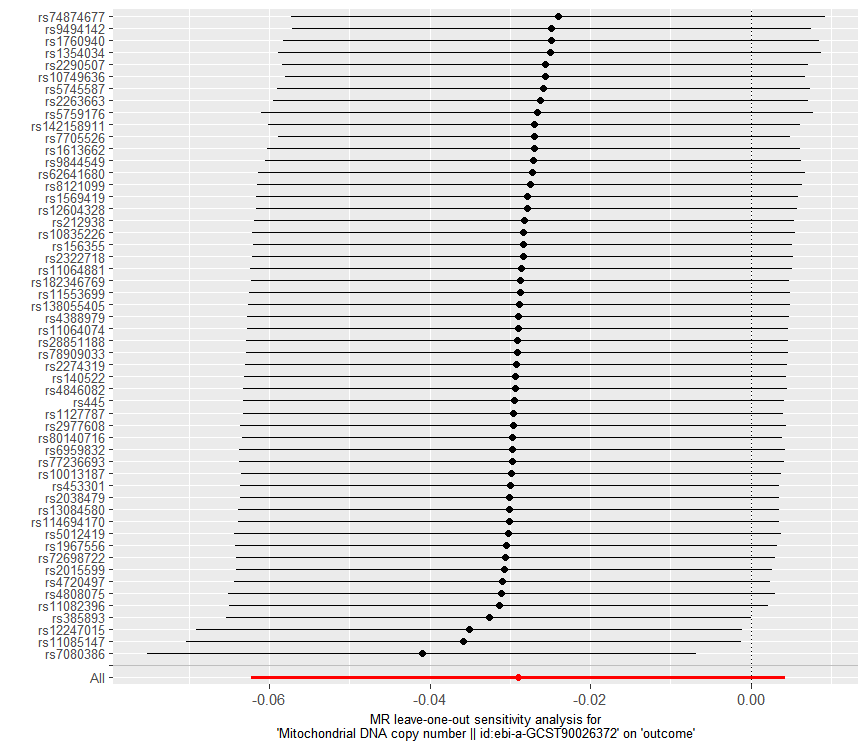

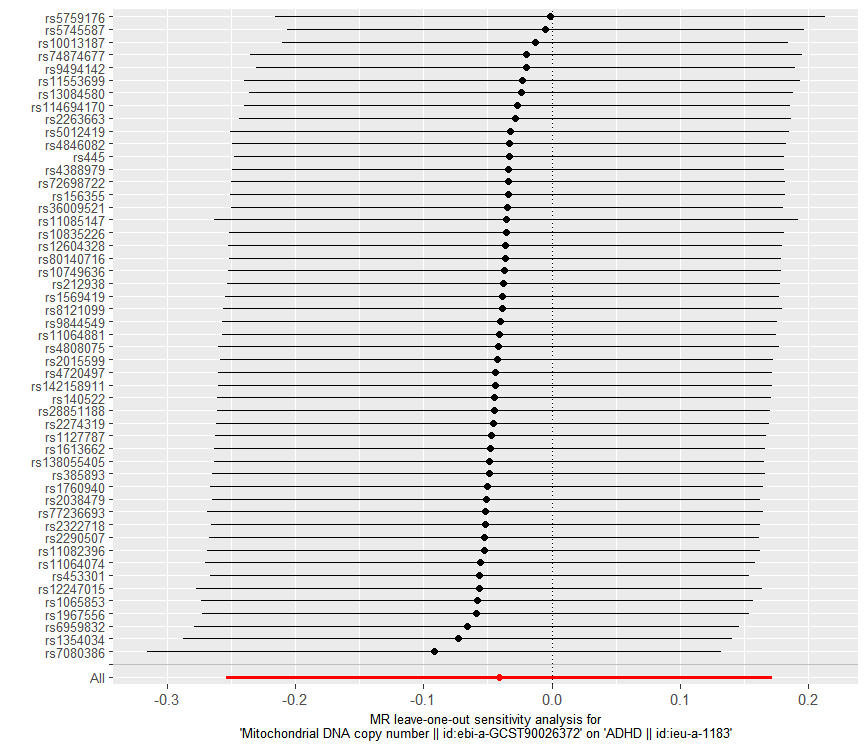

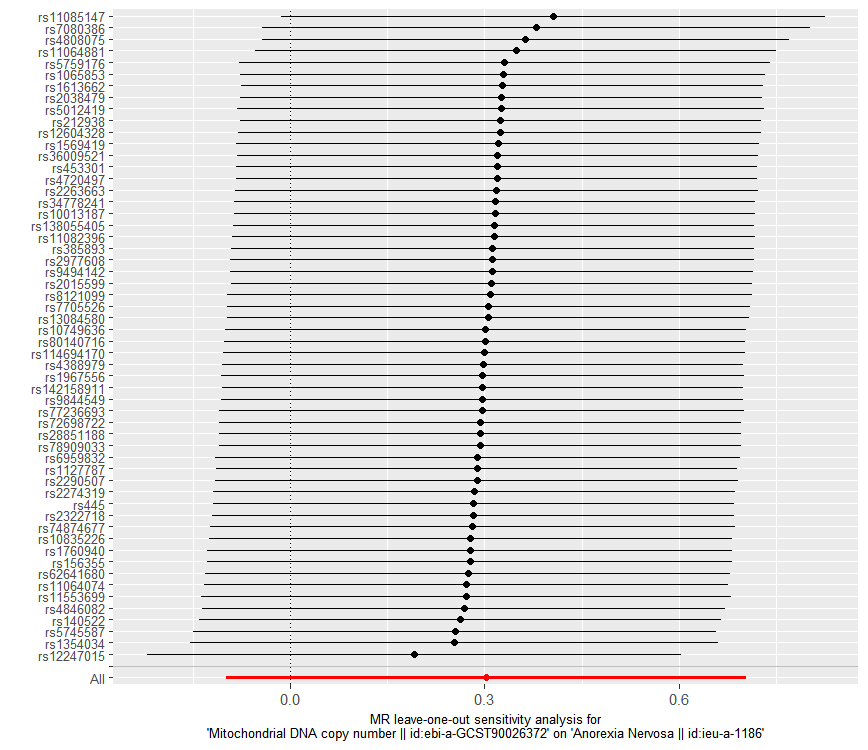

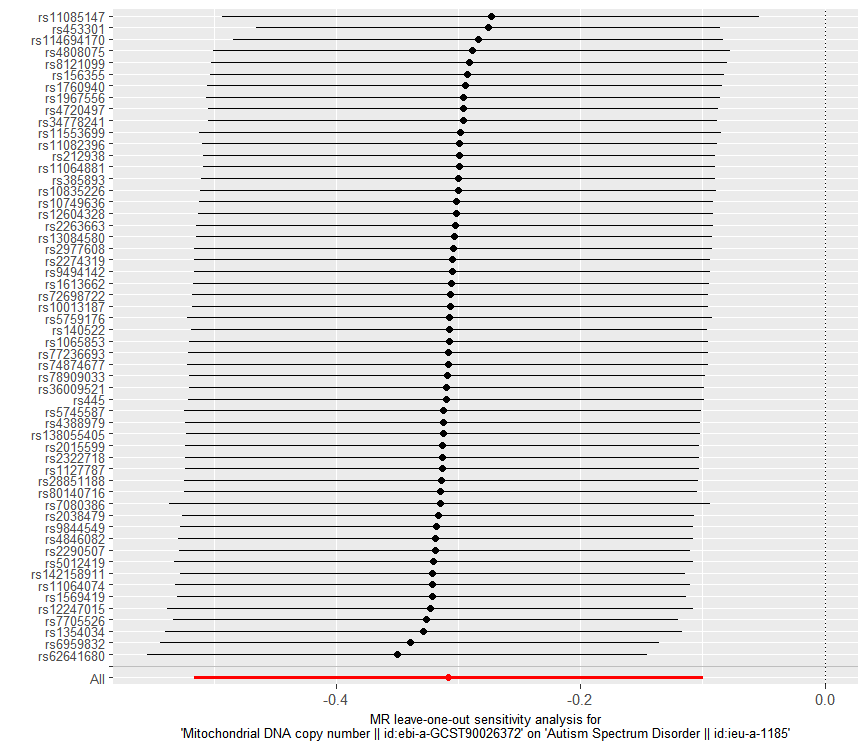

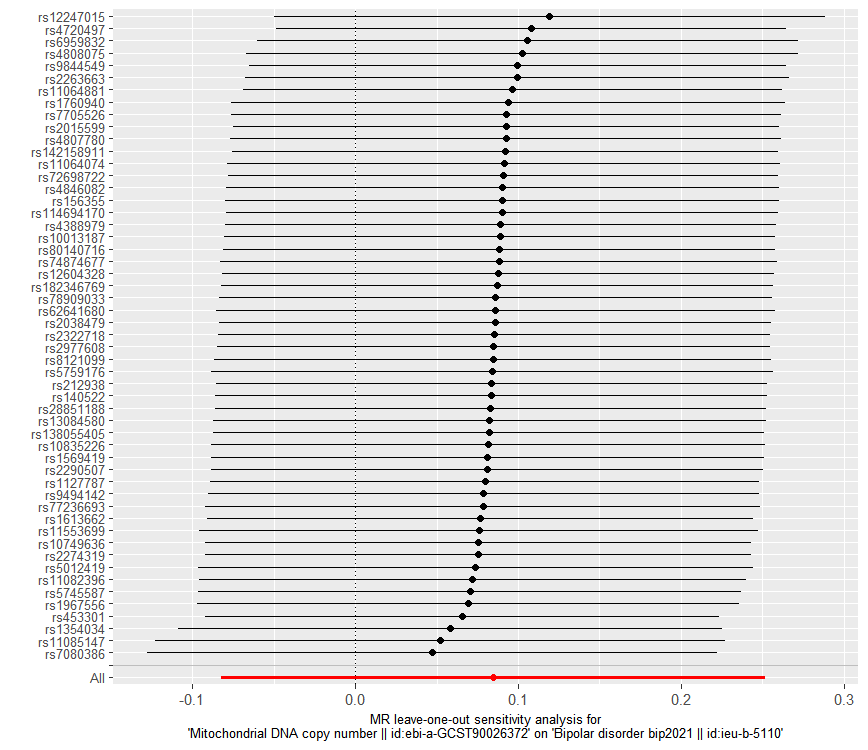

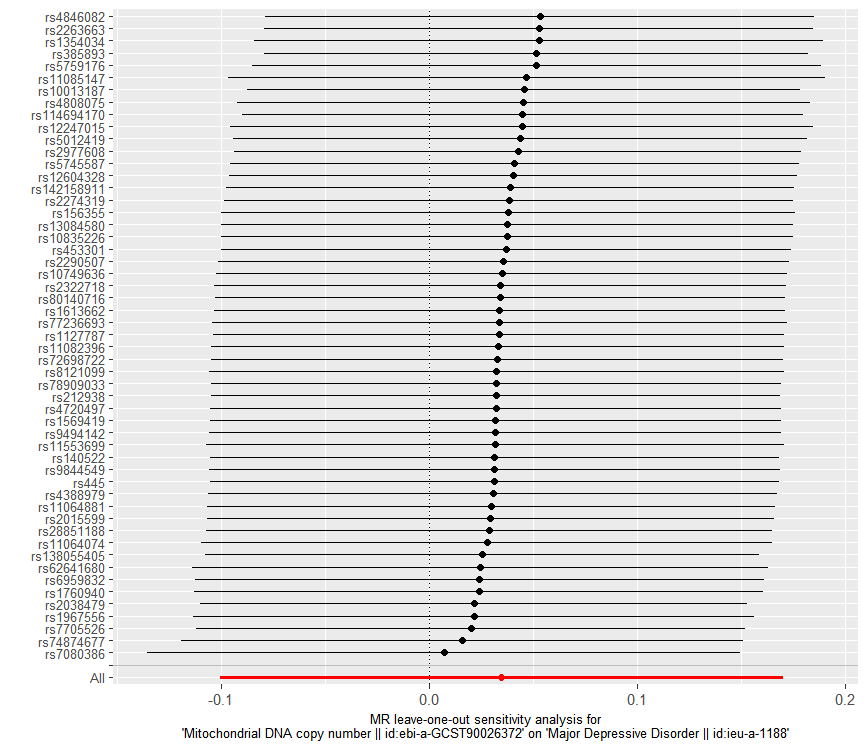

**D**

**C**

**E**

**F**

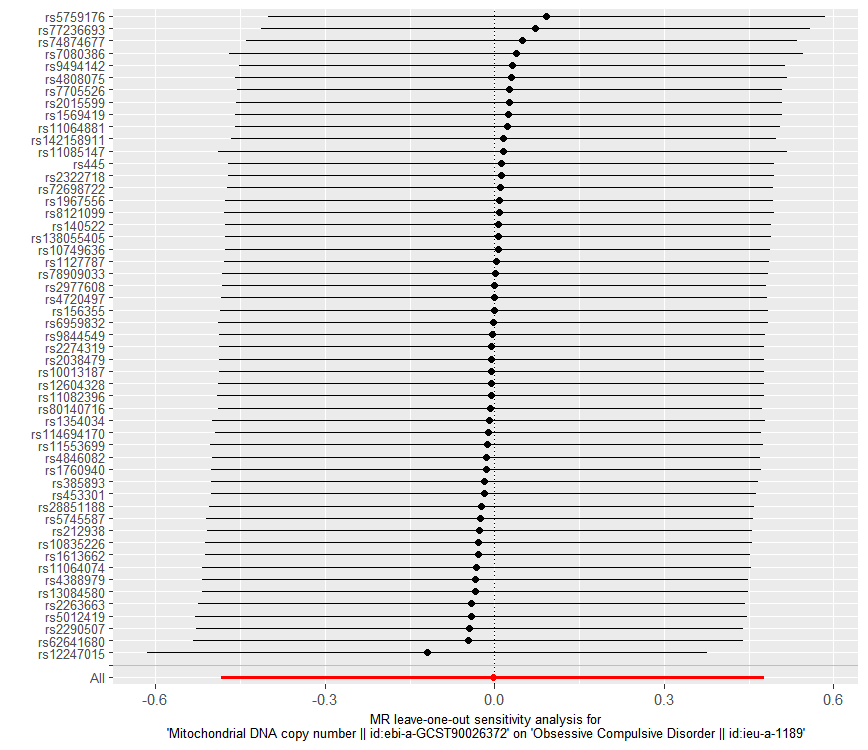

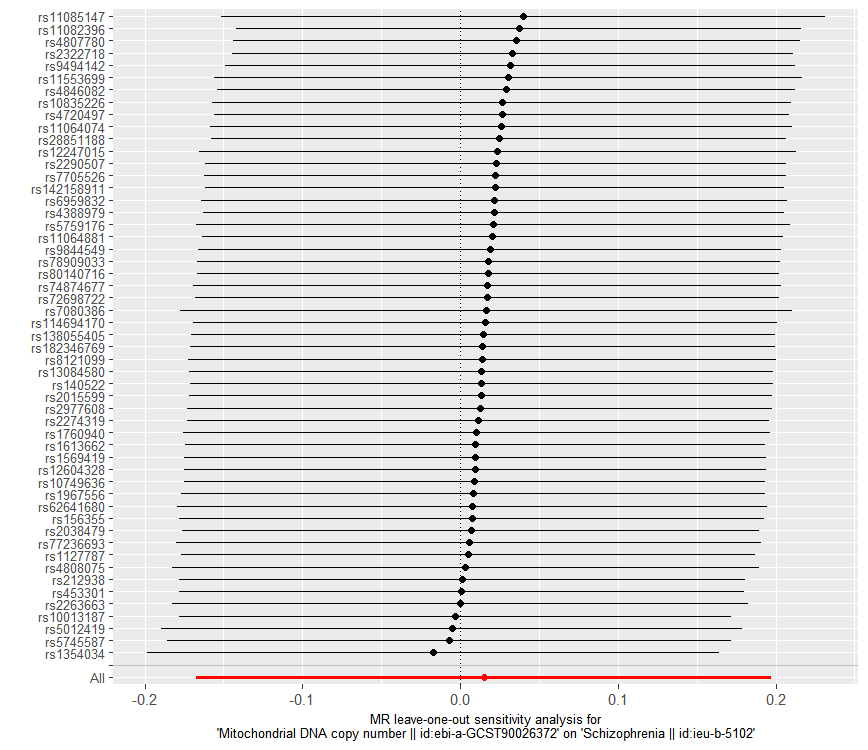

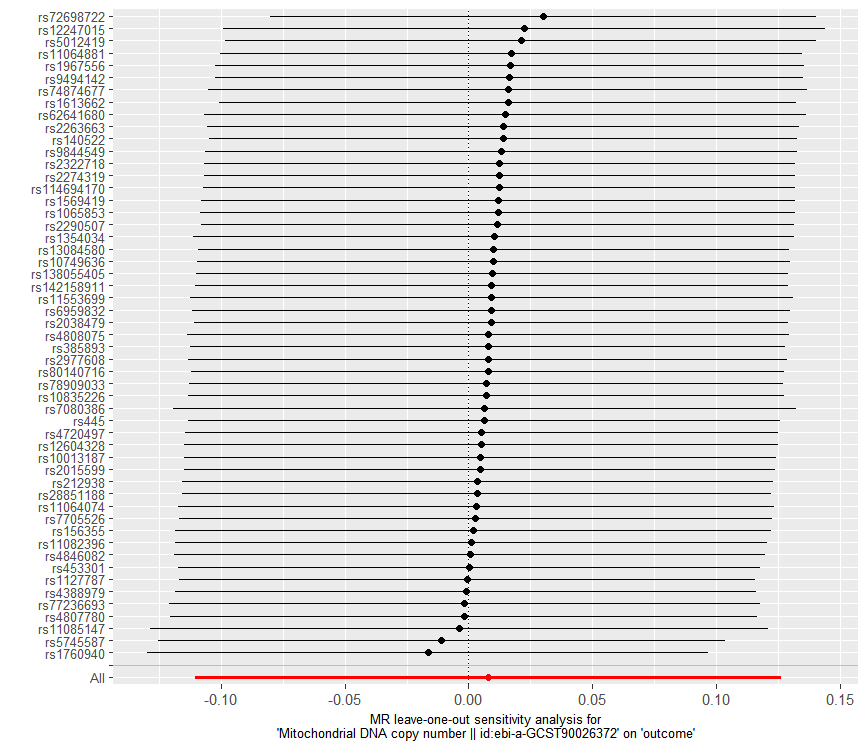

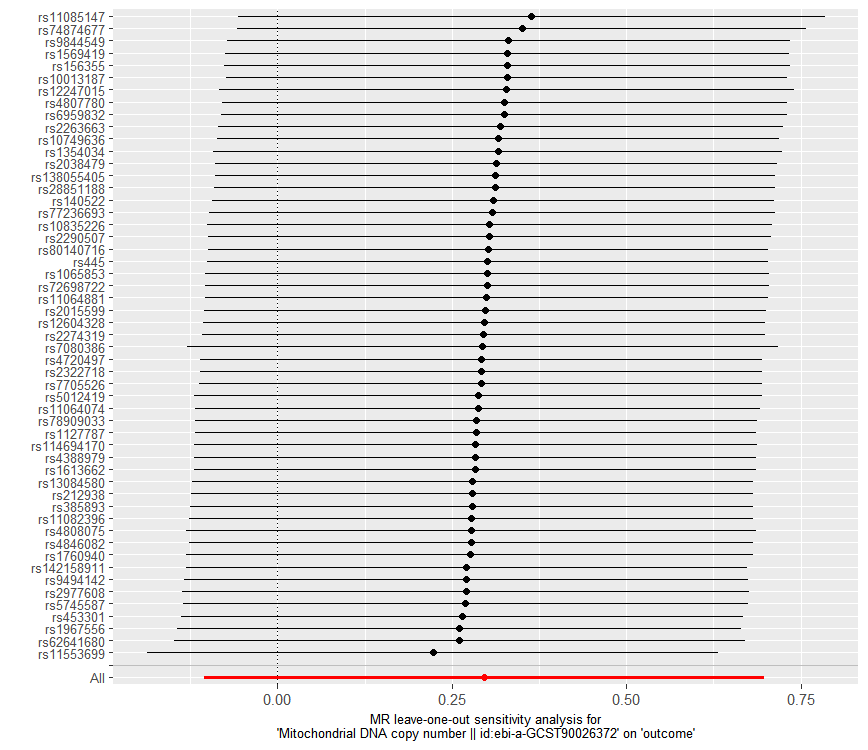

**G**

**H**

**I**

**J**

**[Figure S1](https://europepmc.org/articles/PMC9349767/figure/jmv28008-fig-0003/" \t "figure)** **The forward MR analyses:** **Plots of “leave-one-out” analyses for MR analyses of the causal effect of mtDNA copy number with the risk of neuropsychiatric disorders.** A Alzheimer’s disease, B attention-deficit/hyperactivity disorder, C anorexia nervosa, D autism spectrum disorder, E bipolar disorder, F major depressive disorder, G obsessive compulsive disorder, H Schizophrenia, I anxiety disorders, J post-traumatic stress disorder. The horizontal lines in the figure represents beta value and its 95% confidence interval [CI] of causal inference, which indicates the genetic effect of the SNP on neuropsychiatric disorders.

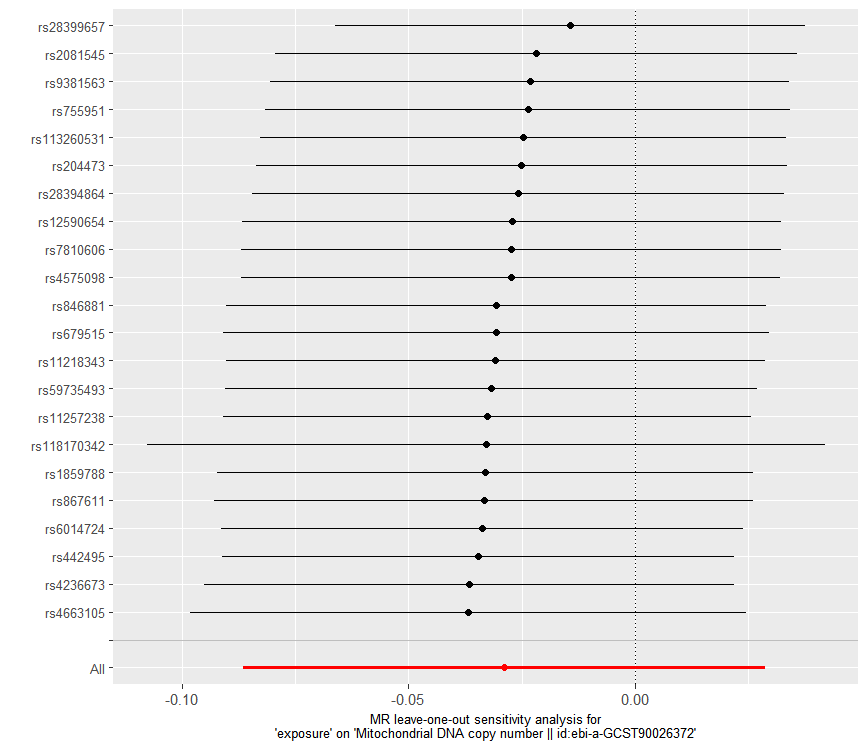

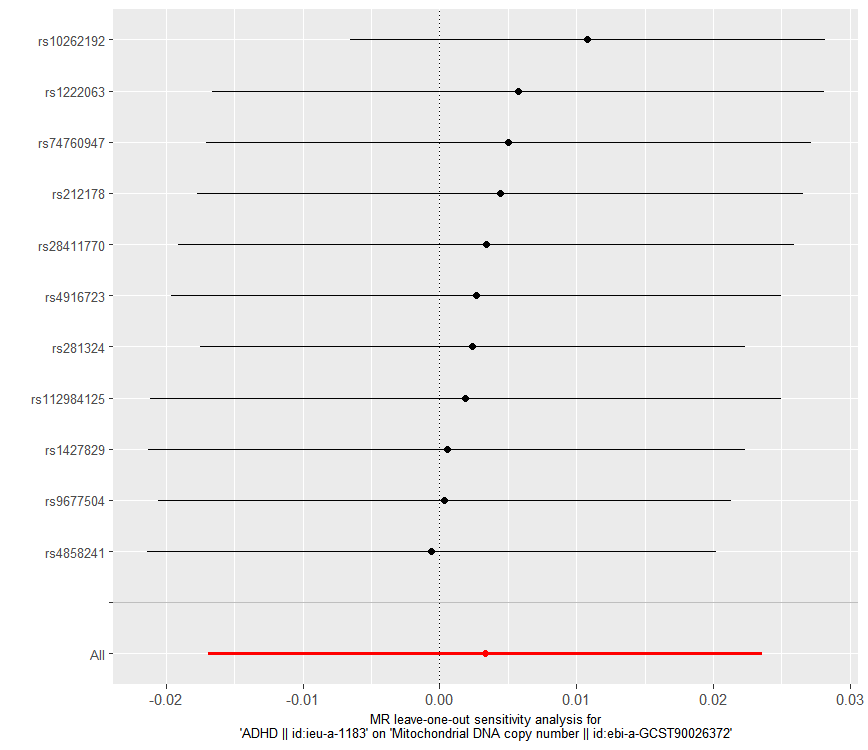

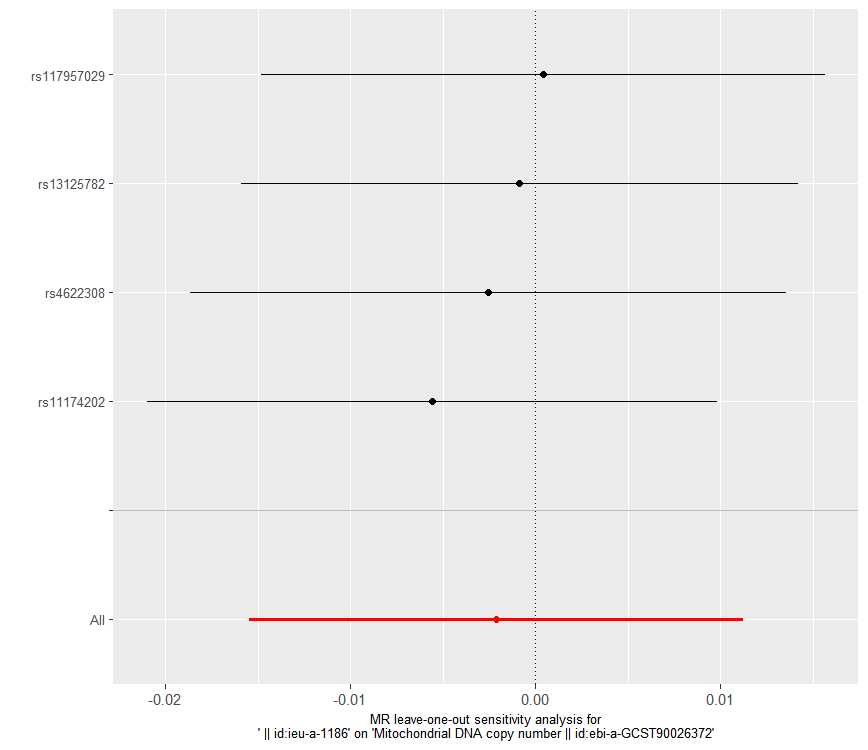

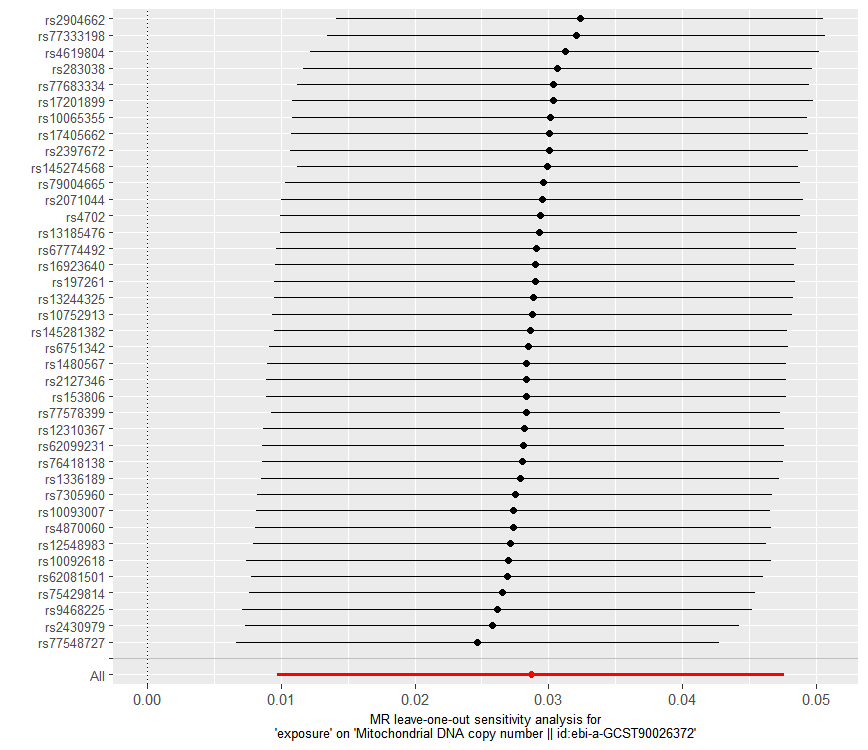

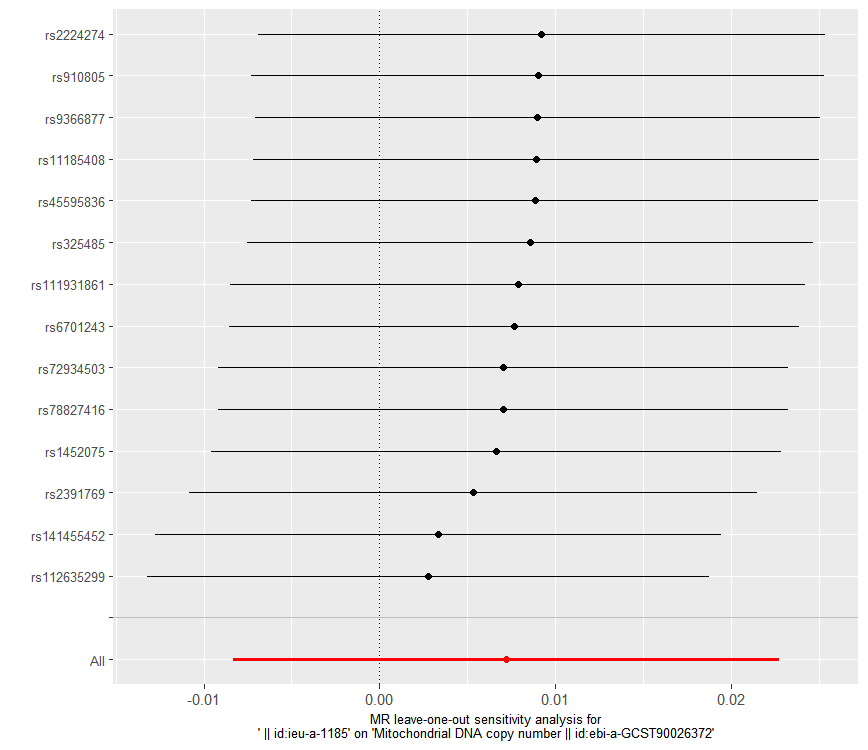

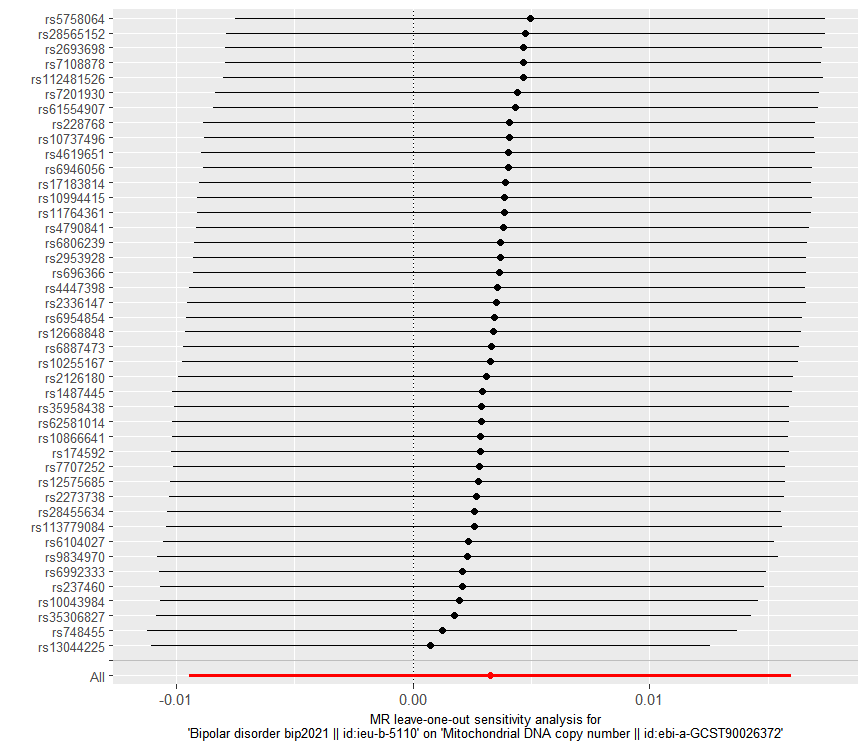

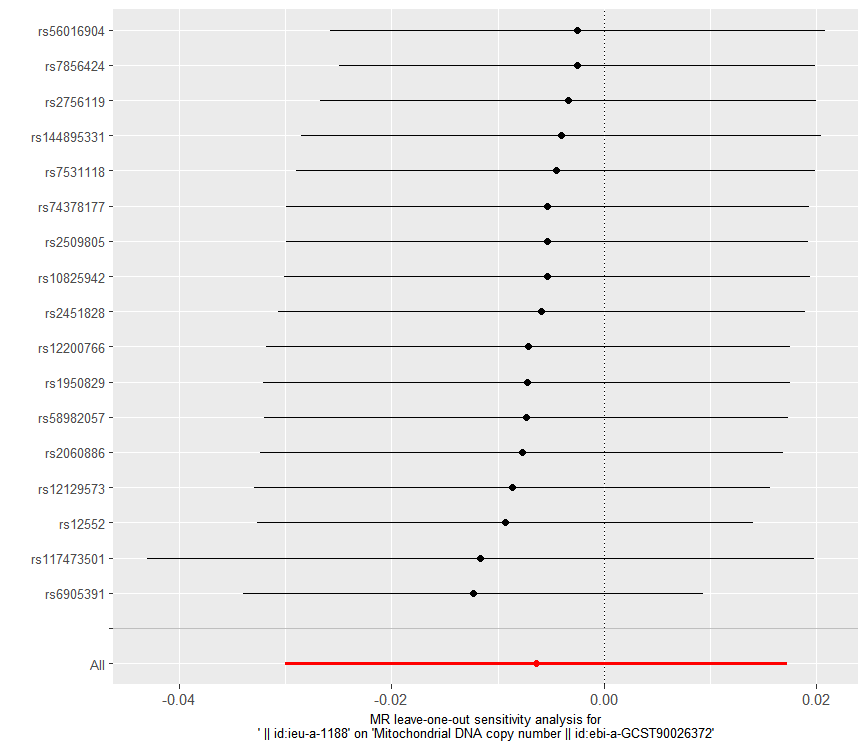

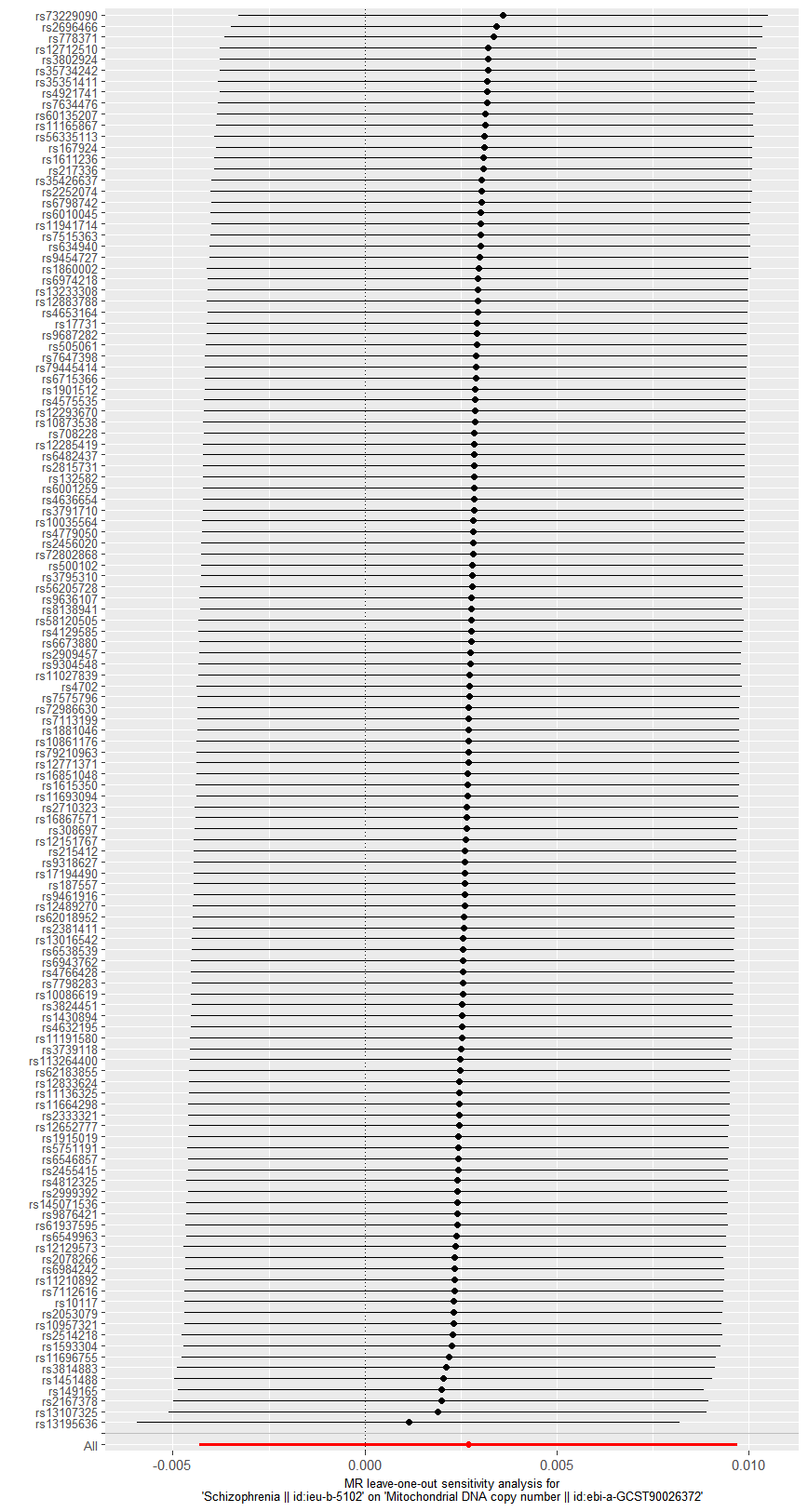

**B**

**A**

**D**

**C**

**F**

**E**

**H**

**G**

**[Figure S2](https://europepmc.org/articles/PMC9349767/figure/jmv28008-fig-0003/" \t "figure)** **The reverse MR analyses: Plots of “leave-one-out” analyses for MR analyses of the causal effect of neuropsychiatric disorders on mtDNA copy number .** A Alzheimer’s disease, B attention-deficit/hyperactivity disorder, C anorexia nervosa, D anxiety disorders, E autism spectrum disorder, F bipolar disorder, G major depressive disorder, H Schizophrenia. The horizontal lines in the figure represents beta value and its 95% confidence interval [CI] of causal inference, which indicates the genetic effect of the SNP on mtDNA copy number.
